# An accurate and rapidly calibrating speech neuroprosthesis

**DOI:** 10.1101/2023.12.26.23300110

**Authors:** Nicholas S. Card, Maitreyee Wairagkar, Carrina Iacobacci, Xianda Hou, Tyler Singer-Clark, Francis R. Willett, Erin M. Kunz, Chaofei Fan, Maryam Vahdati Nia, Darrel R. Deo, Aparna Srinivasan, Eun Young Choi, Matthew F. Glasser, Leigh R. Hochberg, Jaimie M. Henderson, Kiarash Shahlaie, David M. Brandman, Sergey D. Stavisky

**Affiliations:** Departments of Neurological Surgery; Computer Science; Biomedical Engineering University of California Davis, Davis, CA, USA; Departments of Neurosurgery; Electrical Engineering; Mechanical Engineering; Wu Tsai Neurosciences Institute; Howard Hughes Medical Institute, Stanford University, Stanford, CA, USA; Departments of Radiology and Neuroscience, Washington University School of Medicine, Saint Louis, MO, USA; School of Engineering and Carney Institute for Brain Sciences, Brown University, Providence, RI, USA; VA RR&D Center for Neurorestoration and Neurotechnology, VA Providence Healthcare, Providence, RI; Center for Neurotechnology and Neurorecovery, Department of Neurology, Massachusetts General Hospital, Harvard Medical School, Boston, MA

## Abstract

Brain-computer interfaces can enable rapid, intuitive communication for people with paralysis by transforming the cortical activity associated with attempted speech into text on a computer screen. Despite recent advances, communication with brain-computer interfaces has been restricted by extensive training data requirements and inaccurate word output. A man in his 40’s with ALS with tetraparesis and severe dysarthria (ALSFRS-R = 23) was enrolled into the BrainGate2 clinical trial. He underwent surgical implantation of four microelectrode arrays into his left precentral gyrus, which recorded neural activity from 256 intracortical electrodes. We report a speech neuroprosthesis that decoded his neural activity as he attempted to speak in both prompted and unstructured conversational settings. Decoded words were displayed on a screen, then vocalized using text-to-speech software designed to sound like his pre-ALS voice. On the first day of system use, following 30 minutes of attempted speech training data, the neuroprosthesis achieved 99.6% accuracy with a 50-word vocabulary. On the second day, the size of the possible output vocabulary increased to 125,000 words, and, after 1.4 additional hours of training data, the neuroprosthesis achieved 90.2% accuracy. With further training data, the neuroprosthesis sustained 97.5% accuracy beyond eight months after surgical implantation. The participant has used the neuroprosthesis to communicate in self-paced conversations for over 248 hours. In an individual with ALS and severe dysarthria, an intracortical speech neuroprosthesis reached a level of performance suitable to restore naturalistic communication after a brief training period.

## Introduction

Communication is a priority for the millions of people living with dysarthria from neurological disorders such as stroke and amyotrophic lateral sclerosis (ALS)^1^. People living with diseases that impair communication report increased rates of isolation, depression, and decreased quality of life^2,3^; losing communication often determines if a person will pursue or withdraw life-sustaining care in advanced ALS^4^. While existing augmentative and assistive communication technologies such as head or eye trackers are available, they suffer from low information transfer rates and become increasingly onerous to use as patients lose voluntary muscle control^5^. Brain-computer interfaces are a promising communication technology that can directly decode the user’s intended speech from cortical neural signals^6^. Efforts to develop a speech neuroprosthesis are built on largely offline (post hoc) studies using data from able-bodied speakers undergoing electrophysiological monitoring for clinical purposes (e.g. ^7–15^, but see ^16^). Several groups have performed online (real-time) brain-computer interface studies specifically to restore lost speech using chronically implanted electrocorticography (ECoG)^17–20^ and intracortical multielectrode arrays^21^. Two recent reports have established ‘brain-to-text’ speech performance^19,21^ by decoding the neural signals generated by attempted speech into phonemes (the building blocks of words), and assembling these phonemes into words and/or sentences displayed on a computer screen. These studies achieved communication performance, quantified using the word error rate metric, of 25.5% with a 1,024-word vocabulary^19^ and 23.8% with a 125,000-word vocabulary^21^, which is insufficient for reliable general-purpose communication. These prior studies also required at least 13 days of recording to collect sufficient training data to obtain that level of performance (17.7 training data hours in ^19^, 16.8 hours in^21^).

Here, we report an intracortical speech neuroprosthesis capable of providing access to a comprehensive 125,000-word vocabulary, with low training data requirements. We report that our clinical trial participant, living with advanced ALS and severe dysarthria, achieved very high accuracy brain-to-text communication (word error rates consistently below 5%), with useful function beginning on the very first day of use.

## Methods

### Study participant

A left-handed man in his 40’s (referred to as ‘SP2’ in this preprint rather than the actual trial participant designation, which the participant is familiar with, as per medRxiv policy) with amyotrophic lateral sclerosis (ALS) was enrolled into the BrainGate2 pilot clinical trial. At the time of enrollment, he had no functional use of his upper and lower extremities and had severe dysarthria (ALSFRS-R = 23). For the duration of this study (8 months following implantation), he has maintained a modified mini-mental status exam score of 27 (the highest score attainable). At the time of this report, he still retains eye and neck movements, but has limited orofacial movement with a mixed upper- and lower-motor neuron dysarthria resulting in a monotone, low-volume, hypernasal speech with off-target articulation. He requires non-invasive respiratory support at night, and does not have a tracheostomy. When speaking with non-expert listeners, he is unintelligible (Audio 1): his oral motor tasks on the Frenchay Dysarthria Assessment-2 were an “E” rating, representing non-functioning or profound impairment. When speaking to expert listeners, he communicates at 6.8 ± 5.6 (mean ± standard deviation) correct words per minute. His typing speed using his gyroscopic headmouse (Zono 2, Quha, Nokia, Finland) is 6.3 ± 1.3 correct words per minute (Fig. S1). The severity of dysarthria has remained stable during the time-period of this report, including the immediate postoperative period. Additional participant details are in Section S1.01.

### Surgical implantation

The goal of the surgery was to implant four microelectrode arrays (NeuroPort Array, Blackrock Neurotech, Salt Lake City, Utah, USA) into the precentral gyrus, an important cortical target for coordinating the motor activities related to speech^17,19,21^. Each single electrode has one recording site ∼50 μm in size, and is designed to record from a single or a small number of cortical neurons. A microelectrode array is 3.2 × 3.2 mm in size, has 64 recording sites in an 8 × 8 grid arrangement, and is inserted 1.5 mm into the cortex using a specialized high-speed pneumatic inserter.

We performed a left-sided 5 × 5 cm craniotomy under general anesthesia. Care was taken to avoid placing the microelectrode arrays through large vessels on the cortical surface, identified by visual inspection. Two arrays are connected to one percutaneous pedestal designed to transmit the neural recordings to external computers. We placed two percutaneous pedestals, for a total of 256 recording sites. Reference wires were placed in both the subdural and epidural spaces.

Our participant underwent surgery in July 2023, had no study-related serious adverse events, and was discharged on postoperative day 3. From initial incision to closure, the operation took 5 hours to perform. We began collecting data in August 2023, 25 days after surgery.

### Recording array implant locations and decoding contributions

Prior to implanting arrays in the precentral gyrus, we identified the central sulcus on an anatomical MRI scan, and confirmed that our study participant was left-hemisphere language dominant on functional MRI, using standard clinical fMRI tasks (sentence completion, silent word generation, silent verb generation, and object naming). In addition to pre-operative morphological MRI and fMRI studies, we further refined our implantation targets using the Human Connectome Project’s multi-modal MRI-derived cortical parcellation precisely mapped to the participant’s brain^22^ (Fig. 1a, Fig. S2, Section S1.02). We targeted language-related area 55b^23^ and three areas in the precentral gyrus associated with motor control of the speech articulators: premotor areas dorsal 6v (d6v) and ventral 6v (v6v), and primary motor cortex (area 4; Fig. S2). Our choice of targeting speech motor cortex was motivated by our previous study, which found that two 6v arrays provided informative signals for speech decoding^21^. Compared to that previous report, the implantation of four micro-electrode arrays doubles the number of electrodes implanted into the ventral precentral gyrus and doubles the corresponding cortical area covered.

**Figure 1.**
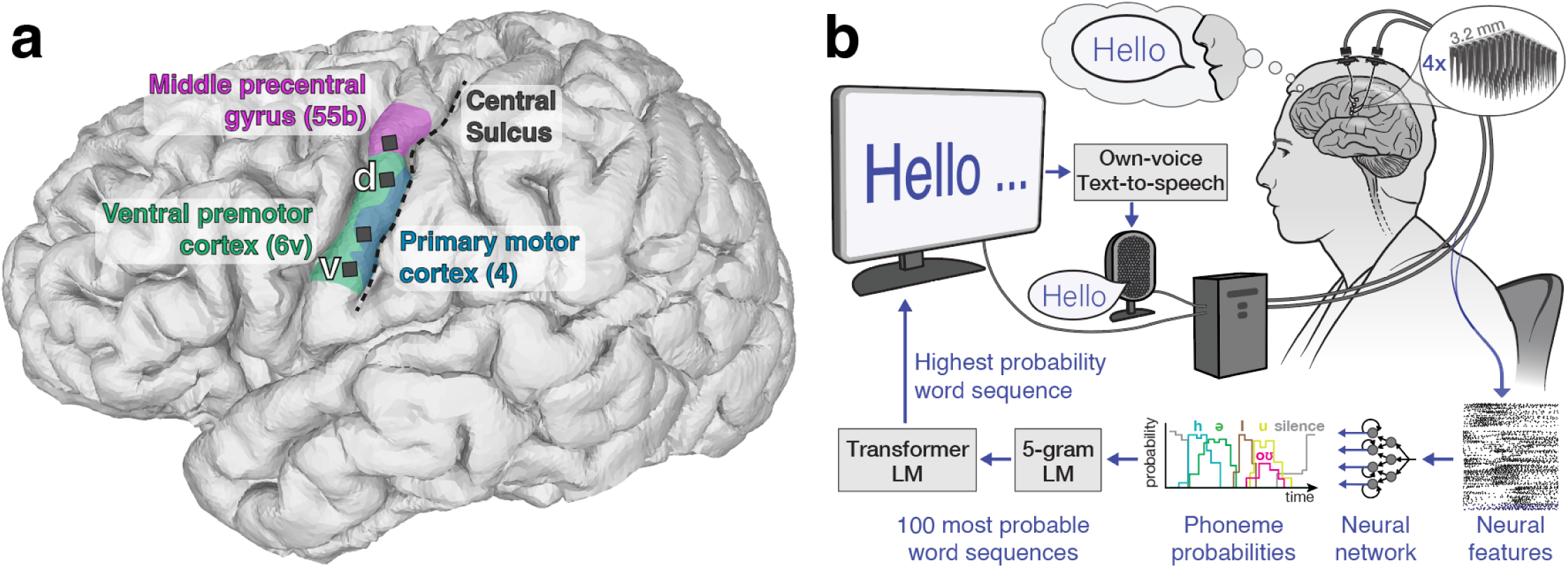
Electrode locations and speech decoding setup. **a**, Approximate microelectrode array locations, represented by black squares, superimposed on a 3d reconstruction of the participant’s brain. Colored regions correspond to the Human Connectome Project’s multi-modal atlas of cortical areas^22^ aligned to the participant’s brain using the Human Connectome Project’s MRI protocol scans before implantation, concordant with the precentral gyrus on a MNI template brain (Figure S11). **b**, Diagram of the brain-to-text speech neuroprosthesis. Cortical neural activity is measured from the left ventral precentral gyrus using four 64-electrode Utah arrays. Machine learning techniques decode the cortical neural activity into an English phoneme every 80 ms. Using a series of language models (LM), the predicted phoneme sequence is translated into a series of words that appear on a screen as the participant tries to speak. At the end of a sentence, an own-voice text-to-speech algorithm vocalizes the decoded sentence designed to emulate the participant’s voice prior to developing ALS (Section S5).

### Real-time acquisition and processing of neural data

Neural activity was transmitted from the pedestals to a computer system designed to decode the activity in real-time (Fig. 1b). A signal processing system (NeuroPort System, Blackrock Neurotech) was used to acquire signals from the two pedestals (Fig. S3) and send them to a series of commercially available computers running custom software^24^ (Section S1.5) for real-time signal processing (Section S1.4) and decoding (Sections S2, S3). The system sits on a wheeled cart and requires the computers to be connected to standard wall outlets. Blackrock Neurotech was not involved in the data collection or reporting in this study, and had no oversight regarding the decision to publish.

### Speech task designs

The study consisted of 84 data collection sessions over the course of 32 weeks (Section S1.06; Table S2) and took place in the participant’s home. Each session consisted of a series of task blocks, lasting approximately 5-30 minutes, wherein the participant used the neuroprosthesis. Between blocks, he would take breaks, eat meals, etc. One session was performed on any day. During each block, the participant used the system in two different ways: 1) an instructed-delay Copy Task (Videos 1, 2 and Section S1.07); and 2) a self-paced Conversation Mode (Videos 4, 5 and Section S1.08). The instructed-delay task consisted of words being presented on a computer screen, and the participant attempted to say the words after a visual/audio cue^21^. The self-paced Conversation Mode involved the participant trying to say whatever he wanted, from the 125,000 word dictionary, in an unstructured conversational setting. In both tasks, speech decoding occurred in real-time: as he spoke, the cortical activity at the four micro-electrode arrays were recorded and interpreted, and the predicted words were presented on the screen. Completed sentences were read aloud by a computer program and, in later sessions, automatically punctuated (Sections S4 and S3.03). The neuroprosthesis could also send the sentence to the participant’s personal computer by acting as a bluetooth keyboard, which allowed him to use it for activities such as writing emails (Section S1.08). Sampled phoneme and words used for decoder training accumulated over the course of the study (Figure S4).

### Decoding speech

Our neuroprosthesis was designed to translate the participant’s cortical neural activity into words when he attempted to speak (Section S2, Figs. S5, S6). No microphone input was used for decoding, and we found no evidence of acoustic or vibration-related contamination in the recorded neural signals (Section S4.03, Fig. S7). Every 80 ms, the activity from the population of cortical neurons was used to predict the most likely English phoneme (the building block of sounds of a language) being attempted. Phoneme sequences were then combined into words using an openly-available language model^21^. Next, we applied two further open-source language models to translate the sequence of words into the most likely English sentence. Hence, the neural activity was the only input for all subsequent language-model refinements (Section S3, Fig. S8), as described in ^21^. Machine learning techniques enabled high accuracy decoding; data from different days were combined to continuously calibrate the decoder, enabling maintenance of high performance during neuroprosthesis use (Section S2 and Figs. S9, S10).

### Evaluation

We used two measures to analyze the speech decoding performance: phoneme error rate and word error rate, consistent with previous speech decoding studies^17,19,21^. These measures are the ratio of phonemic or word errors to the total number of phonemes or words expected to be decoded, respectively. An error is defined as when an insertion, deletion, or substitution is needed to have the decoded sentence match the intended sentence. The phoneme error rate can be understood as the system’s ability to translate cortical neural activity into phonemes without language models (to make this explicit, we label it as ‘raw’ phoneme error rate in figures), and word error rate as an estimate of overall communication accuracy. Data was collected in continuous blocks (lasting 5-30 minutes in length) which were separated by short breaks. Blocks were either “training blocks” in which data were collected for decoder training or optimization, or predetermined “evaluation blocks” used to measure and report neuroprosthesis performance. Reported error rates are aggregated over all evaluation sentences for each session (Section S1.09). The first-ever closed-loop block (session 1) was excluded from evaluation because the participant cried with joy as the words he was trying to say correctly appeared on-screen; the research team paused the evaluation until after the participant and his family had a chance to celebrate the moment.

### Statistical analyses

Results for each analysis are presented with 95% confidence intervals or as mean ± standard deviation. Confidence intervals were estimated by randomly resampling each dataset 10,000 times with replacement, and have not been adjusted for multiplicity. The evaluation metrics for decoding performance (phoneme error rate and word error rate) were chosen before the start of data collection (Section S1.09).

## Results

### Online decoding performance

In the first session, the participant attempted to speak prompted sentences constructed from a 50-word vocabulary^17^. We recorded 213 sentences (30 minutes) of Copy Task data, which were used to calibrate the speech neuroprosthesis. Next, we decoded his neural cortical activity in real-time as he tried to speak. The neuroprosthesis decoded his attempted speech with a word error rate of 0.44% (95% confidence interval [CI], 0.0% to 1.4%). We replicated this result for 50-word vocabulary decoding in the second research session, in which all of the participant’s attempted sentences were decoded correctly (0% word error rate; Fig. 2).

**Figure 2.**
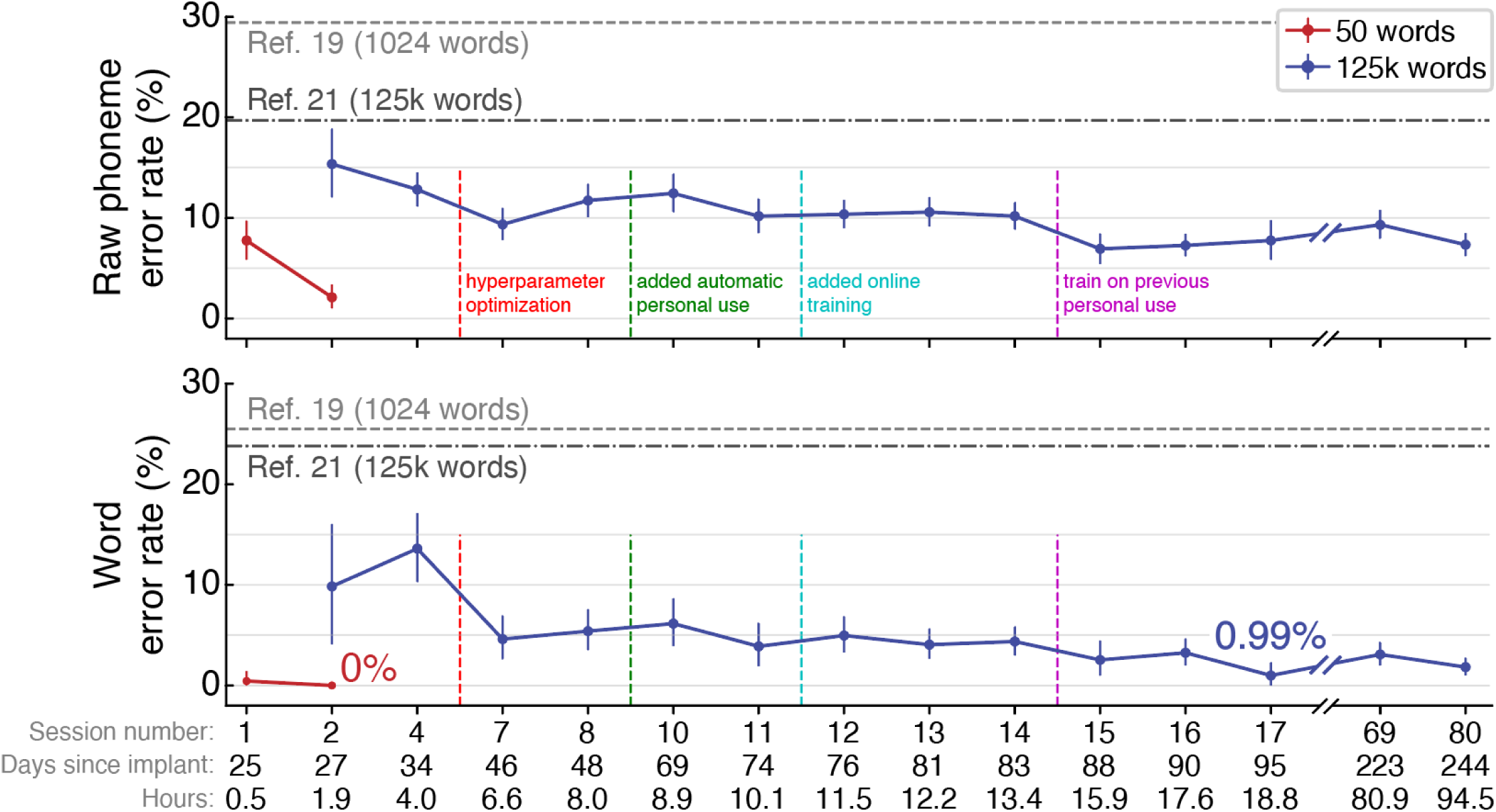
Online speech decoding performance. Phoneme error rates (top) and word error rates (bottom) are shown for each session for two vocabulary sizes (50 versus 125,000 words). Reference error rates are plotted (horizontal dashed lines) for two previous speech neuroprosthesis studies^19,21^. The horizontal axis displays the research session number, the number of days since arrays implant, and the cumulative hours of neural data used to train the speech decoder for that session. Aggregate error rates across all evaluation sentences are shown for each session (mean ± 95% confidence interval). Vertical dashed lines represent when decoder improvements were introduced. Fig. S20 shows phoneme and word error rates for individual blocks.

In this second research session, we also expanded the vocabulary of the neuroprosthesis from 50 words to over 125,000 words, which encompasses the majority of the English language^25^. We collected an additional 260 sentences of training data (1.4 hours). After being trained on these additional data, the neuroprosthesis decoded the participant’s attempted speech with a word error rate of 9.8% (95% CI, 4.1% to 16.0%; Fig. 2); offline analyses suggest that even better first-session performance may have been possible given the information content of the recorded neural signals (Figs. S12, S13). Performance continued to improve in subsequent research sessions as we collected more training data and adapted innovations for incorporating new data more effectively^26^. The neuroprosthesis achieved a word error rate of 2.5% (95% CI, 1.0% to 4.5%) by session 15, and high accuracy decoding performance was maintained through session 84, more than eight months after implant. Average Copy Task decoding performance in the final 5 evaluation sessions had a word error rate of 2.5% (95% CI, 2.0% to 3.1%) at the participant’s self-paced speaking rate of 31.6 words per minute (95% CI, 31.2% to 32.0%; Fig. S1), with individual day’s average word error rates ranging from 1.0% to 3.3% (Table S3). The neuroprosthesis’ communication rate far exceeded the participant’s standard means of communication using a head mouse or skilled interpreter (Fig. S1a).

The system achieved high accuracy at the start of sessions, maintaining an average word error rate of 4.9% (95% CI, 3.7% to 6.2%) over the first 50 sentences of seven sessions (Fig. S14). Offline analyses indicated that this rapidly-available communication capability was enabled by the neuroprosthesis being able to maintain accurate decoding for at least twenty days without any new training data, and for multiple months when continuously fine-tuning the neuroprosthesis^26^ (Fig. S15). In decreasing order, the most informative electrode arrays for decoding speech were in areas ventral 6v (best), 55b, 4, and dorsal 6v (worst; Fig. S16).

Decoding errors tended to occur between phonemes that are articulated similarly (Fig. S17). The speech decoder generalized to new words, and accuracy improved for a given word the more often it appeared in the training data (Fig. S18). It could also generalize to different attempted amplitudes of speech, including non-vocalized speech (Fig. S19).

### Conversational speech using the brain-computer interface

We developed a system for the participant to have conversations via self-initiated speech. Relying exclusively on cortical neural activity, the speech neuroprosthesis detected when he started or stopped speaking and decoded his attempted speech accordingly (Section S1.08, Fig. S21). The system rarely falsely detected that he wanted to speak (Fig. S22). Additionally, the participant had the option to use an eye tracker for selecting actions (Fig. 3a) to i.) finalize and read aloud the sentence, ii.) indicate whether the sentence was decoded correctly or not, or iii.) spell out words letter-by-letter that were not correctly predicted by the decoder (e.g., because they were not in the vocabulary, such as certain proper nouns).

**Figure 3.**
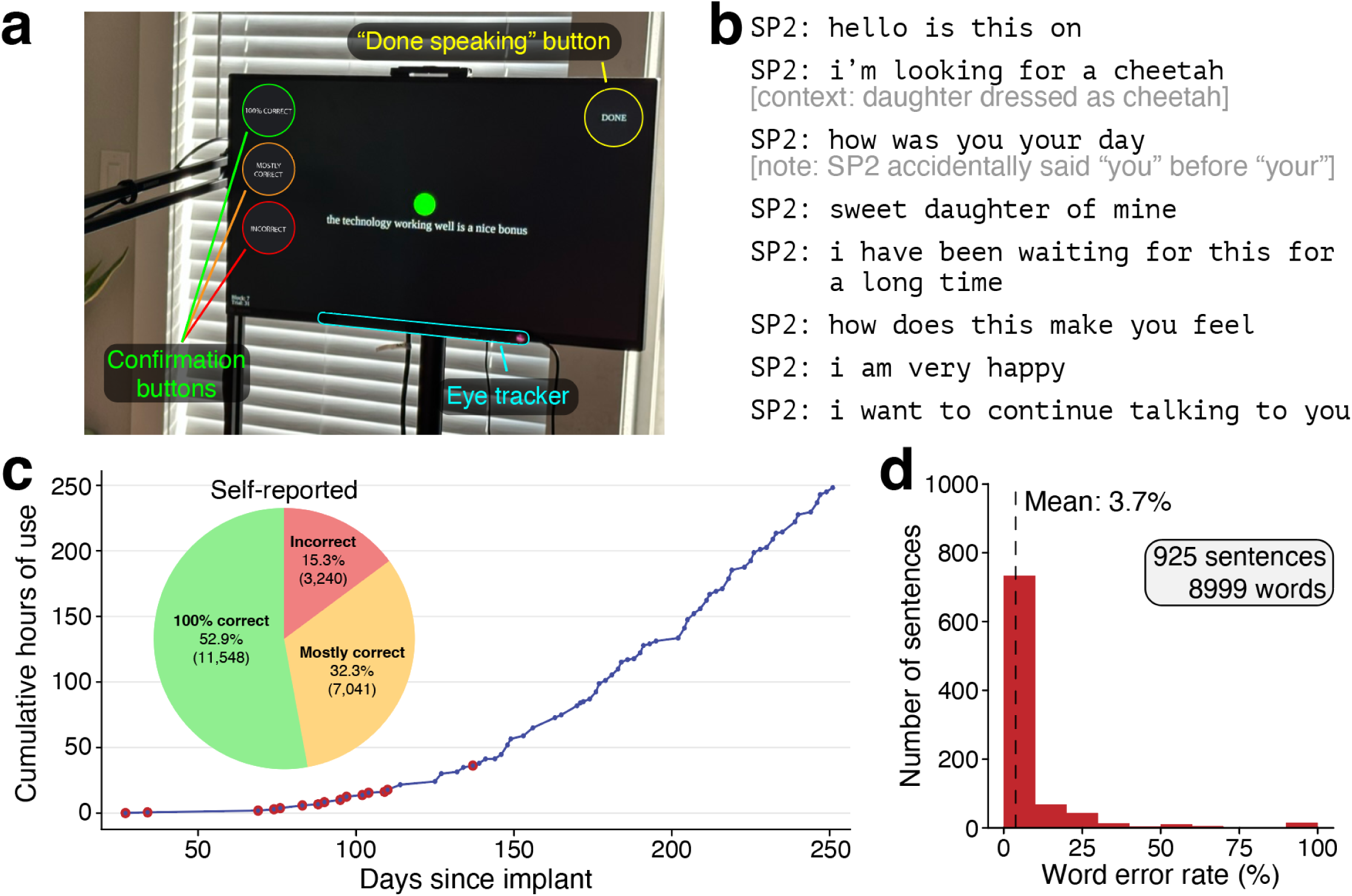
Extensive use of the neuroprosthesis for accurate self-initiated speech. **a**, Photograph of the participant and speech neuroprosthesis in Conversation Mode. The neuroprosthesis detected when he was trying to speak solely based on neural activity, and concluded either after 6 seconds of speech inactivity, or upon his optional activation of an on-screen button via eye tracking. After the decoded sentence was finalized, the participant used the on-screen confirmation buttons to indicate if the decoded sentence was correct. **b**, Sample transcript of our participant using the speech neuroprosthesis to speak to his daughter on the second day of use (Video 3). Additional transcripts are available in Table S4. **c,** Cumulative hours that the participant used the speech neuroprosthesis to communicate with those around him in structured research sessions and during personal use. For sessions represented by points outlined in red, decoding accuracy is quantified in (**d**). The distribution of self-reported decoding accuracy for each sentence across all Conversation Mode data (n = 21,829) is shown in the inset pie chart. Sentences where the participant did not self-report decoding accuracy within 30 seconds of sentence completion are excluded (n = 868). **d**, Evaluating speech decoding accuracy in conversations (n = 925 sentences with known true labels, sourced from red-labeled sessions in (**c**)). The average word error rate was 3.7% (95% CI, 3.3% to 4.3%).

The participant’s first use of the neuroprosthesis for naturalistic communication with his family is exemplified in Fig. 3b (Video 3; Table S4 provides additional transcripts). In subsequent sessions, he utilized the neuroprosthesis for personal use (e.g., Videos 4-5), communicating a total of 1189 sentences from sessions 1-31. For the majority of these Conversation Mode sentences (925 sentences; 77.8%) we were able to confirm the participant’s intended speech through directly asking him, contextual analysis, and examining the decoded phoneme probability patterns. Self-initiated sentences for which we knew the ground-truth were decoded with a word error rate of 3.7% (95% CI, 3.3% to 4.3%; Fig. 3d), although we note that the word error rate reported only for sentences with known ground truth labels may be inflated (Section S1.09). For one session where we validated the ground truth of every sentence (43 sentences, 873 words) with the participant, the word error rate was 2.5% (95% CI, 1.3% to 4.0%). Using the neuroprosthesis, the participant told the research team, “I hope that we are very close to the time when everyone who is in a position like me has the same option to have this device as I do” (Table S4). The participant used the neuroprosthesis in this Conversation Mode during 72 (out of 84 total) sessions over 8 months (248.3 cumulative hours; 22,697 sentences). The longest continuous use of the speech neuroprosthesis in Conversation Mode was 7.7 hours. He used it to perform activities ranging from talking to the research team and his family and friends, to performing his occupation by participating in videoconferencing meetings and writing documents and emails.

## Discussion

Beginning on the first day of device use, a brain-to-text speech neuroprosthesis with 256 recording sites in the left precentral gyrus accurately decoded intended speech in a man with severe dysarthria due to ALS. He communicated using a comprehensive 125,000 word vocabulary on the second day of use. Within 16 hours of use, the neuroprosthesis correctly identified 97.3% of attempted words. To contextualize this 2.7% word error rate, the state-of-the-art for English automated speech recognition (e.g., smartphone dictation) has an approximate 5% word error rate^27^ and able speakers have a 1-2% word error rate^28^ when reading a paragraph aloud. We believe that the high decoding accuracy demonstrated in this study indicates that speech neuroprostheses have reached a level of performance suitable for rapidly and accurately restoring communication to people living with paralysis.

This study’s participant used the brain-to-text speech neuroprosthesis to converse with family, friends, healthcare professionals, and colleagues. His regular means of communication without a neuroprosthesis involved either (1) having expert caregivers interpret his severely dysarthric speech, or (2) using a head-mouse with point-and-click selections on a computer screen. The investigational BrainGate Neural Interface System became his preferred way to communicate with our research team, and he used it on his own time (with a researcher’s assistance to connect and launch the system) to be able to more rapidly write and communicate as part of his occupation and family life. The participant and his family and friends also reported being pleased with the own-voice text-to-speech at the end of each sentence; they indicated that the system’s voice did resemble his own.

This study demonstrated a large reduction in the quantity of training data required to achieve high accuracy decoding. In our previous study^21^, the participant attempted to speak 260-480 sentences at the start of each day, after which up to ∼30 minutes of computation time was required until the speech neuroprosthesis was ready for use. That previous study’s reported closed-loop results were measured starting 113 days post-implant, and used 16.8 hours of training data, collected over 15 days, to achieve a word error rate of 23.8%. A previous ECoG speech neuroprosthesis required 17.7 hours of training data, collected over 13 days, to reach a word error rate of 25.5%^19^. The new neuroprosthesis demonstrated here provided over 99% accuracy on a limited set of 50 words^17^ after just 30 minutes of training data on the very first day of use. It also achieved over 95% accuracy on a large vocabulary after collecting 6.6 cumulative hours of training data (over 7 sessions), and offline analyses indicate that optimized methods could provide >91% accurate large-vocabulary communication on the first day of use.

Previous studies have reported that intracortical devices require frequent recalibration^29–31^. Motivated by recent studies showing that multiple days of neural data can be used to calibrate an effective decoder for a new day^32,33^, here we demonstrated that a speech decoder trained with multiple previous days of data could similarly be used to provide >95% accuracy at the start of a new session. Future work is needed to establish whether the strategy we employed^26^ can maintain performance indefinitely in the absence of ground-truth labels of intended speech, but it is encouraging that across the 29 session days where the participant used the neuroprosthesis solely for personal use, he only requested to recalibrate the decoder on three occasions. Each calibration took approximately 7.5 minutes, during which twenty Copy Task sentences were displayed to provide ground-truth training labels and thereby rapidly update the decoder.

It is possible that an important factor enabling the higher performance of this study relative to our prior intracortical speech neuroprosthesis^21^ was doubling the number of microelectrodes in speech motor cortex. Our finding that ∼200 electrodes in these regions is sufficient for very high accuracy brain-to-text communication provides an important design parameter to guide ongoing efforts to build neural interface hardware that can reach patients at scale. Using an improved phoneme-to-sentences language model (relative to ^21^) also improved performance (Fig. S8), and the participant’s slow speaking rate (Fig. S1) may also have contributed.

In addition to recording from two arrays in the putative ventral portion of area 6v (speech motor cortex) as in ^21^, we also targeted one array each into two areas which, to our knowledge, have not previously been recorded from with multielectrode arrays: area 4 (primary motor cortex, which in humans is often in the sulcus^22^ and thus largely not accessible with Utah arrays) and area 55b. We found that the strongest phoneme encoding was from the array in ventral 6v, which is consistent with our previous participant^21^. The array in area 4 also showed high phoneme encoding, as did the array in area 55b, which has recently been proposed as an important node in the wider speech production network^23^. We note that these brain area descriptions are estimations based on precisely aligning the participant’s brain to a Human Connectome Project derived atlas using multi-modal MRI.

## Limitations

As with other recent clinical trial reports in the nascent field of implanted speech brain-computer interfaces^17–19,21^, this study involved a single participant. Future work with additional participants is needed to establish the across-individual distribution of performances for speech decoding. Whether similar results can be expected in future users may depend on whether the signal-to-noise ratio of this participant’s speech-related neural signal modulation is typical. Nevertheless, these data, when combined with our previous speech decoding results with two 64-electrode arrays in area 6v^21^, demonstrate both successful initial replication and subsequent methodological improvements of the intracortical speech neuroprosthesis approach. Furthermore, while the demonstrated brain-to-text capabilities can provide widely useful communication, they do not capture the full expressive richness of voice; the more difficult challenge of closed-loop brain-to-voice synthesis remains an active area of speech neuroprosthesis research^19,34^. Notable steps that remain before brain-to-text neuroprostheses are likely to be adopted at scale include removing the percutaneous connector (i.e., making the electrodes fully implanted with wireless power and telemetry), reducing the size and number of computers used for processing, and automating the software so that users and their care partners can operate it independently.

### Decoding speech in patients with other neurological impairments

The participants in both this study and our previous report^21^ had dysarthria due to ALS. Further work is needed to assess whether similar methods will work for other etiologies of dysarthria. Given that we recorded from ventral precentral gyrus, which is upstream of the neuronal injury incurred in many conditions, and that recent ECoG speech neuroprostheses were demonstrated in two individuals with brainstem stroke^17–19^, we predict that this approach will also work in other conditions^35^.

We note that the participant retains voluntary (albeit impaired) bulbar muscle control, normal sensation, and normal hearing. We do not know whether our approach will continue to restore communication should he develop anarthria. We also do not know if our approach will work in patients who already have anarthria or have hearing impairments.

### Long-term stability of neural decoding

Prior reports have described that neural recording quality can decrease over years with the type of microelectrode array used in this study (e.g., ^36^). While intracortical neural decoding of attempted hand movements has been shown to be sustained beyond five years^37–40^, our study reports data only up to 8 months after implantation. It is not known how well decoding performance will be sustained over time.

## Conclusion

Overall, the rapid and highly accurate restoration of full vocabulary, speech-based communication, enabled by an intracortical neuroprosthesis and used by a person with advanced ALS, suggests that this approach may be useful in improving the communication ability and autonomy of people with severe speech and monitor impairments.

### Audio 1 - Demonstration of the participant’s unintelligible dysarthric speech

The participant is attempting to say prompted sentences aloud in an instructed delay Copy Task displayed on the screen in front of him (session 10; see Video 2). He retains intact eye movement and limited orofacial movement with the capacity for vocalization, but is unable to produce intelligible speech. At the end of each sentence, the decoded sentence is read aloud by a text-to-speech algorithm that sounds like his pre-ALS voice. Link to listen online: https://ucdavis.box.com/s/f1qt5p4r9bdqflqgm519p49i10isefp5

### Video 1 - Copy Task speech decoding (session 10)

This video shows the same speech decoding trials as in Audio 1 (session 10). Prompted sentences appear on the screen in front of the participant. When the red square turns green, he attempts to say the prompted sentence aloud while the speech decoder predicts what he is saying in real time. In this video, he is signaling the end of a sentence by using an eye tracker to hit an on-screen “done” button. The participant could also end trials by attempting to squeeze his right hand into a fist, the neural correlates of which were decoded (see Video 2; Section S6). At the end of each trial, the decoded sentence is read aloud by a text-to-speech algorithm that sounds like his pre-ALS voice. Link to view online: https://ucdavis.box.com/s/b7g9jmobangbjvdp28gb8d0d5esy7fuy

### Video 2 - Copy Task speech decoding (session 17)

This video shows another example of Copy Task speech decoding from session 17. Prompted sentences appear on the screen in front of the participant. When the red square turns green, he attempts to say the prompted sentence aloud while the speech decoder predicts what he is saying in real time. In this video, he is signaling the end of a sentence by attempting to squeeze his right hand into a fist, the neural correlates of which are decoded (Section S6). At the end of each trial, the decoded sentence is read aloud by a text-to-speech algorithm that sounds like his pre-ALS voice. Link to view online: https://ucdavis.box.com/s/bj155qbekzc93l5b2bbjd9h7xb4fm68r

### Video 3 - First self-directed use of the speech neuroprosthesis (session 2)

The participant uses the speech neuroprosthesis to say whatever he wants for the first time (125,000-word vocabulary; session 2). He chose to speak to his daughter; the transcript of what he said is also available in Fig. 3b. Because the dedicated Conversation Mode (and speech detection) had not yet been developed during session 2, the participant waits for the onscreen square to turn from red to green before attempting to speak. Link to view online: https://ucdavis.box.com/s/7svzpzq2ldxp0wbqd4ewdry4cc5jdalm

### Video 4 - Conversation Mode speech decoding (session 31)

The participant is using the speech decoder in Conversation Mode to engage in freeform conversation with those around him. The video is muted while conversation partners are speaking for privacy reasons. The speech neuroprosthesis reliably detects when the participant begins attempting to speak, and shows the decoded words on-screen in real time. He can signal the end of a sentence using an on-screen eye tracker button (“DONE” button in the top-right of the screen), or by not speaking for 6 seconds (as he does in this video), after which the neuroprosthesis finalizes the sentence. At the end of each sentence, the decoded sentence is read aloud by a text-to-speech algorithm that sounds like his pre-ALS voice. Finally, the participant uses the eye tracker to confirm whether the decoded sentence was correct or not. Correctly decoded sentences are used to fine-tune the neural decoder online. Link to view online: https://ucdavis.box.com/s/ezpz1fbizsma8cqmavjhhlarce7a2v40

### Video 5 - Conversation Mode speech decoding (session 80)

Another example of the participant using the speech decoder in Conversation Mode to engage in freeform conversation with those around him. The participant is speaking to a researcher about movies. He is able to select on-screen buttons using an eye tracker. The participant interface for Conversation Mode was updated in session 72 to add new features (see Section S1.09), including the ability for him to control when text-to-speech audio is played. The own-voice text-to-speech model was updated in session 55 to sound more lifelike and closer to the participant’s pre-ALS voice (see Section S5.02).

Link to view online: https://ucdavis.box.com/s/f7glntzwvj9h4p79yic5zfrq5844o2z2

## Data Availability

Derivatives of the neural data, including RNN probabilities and language model outputs, which can reproduce the reported performance quantification measurements and figures will be made publicly available on Dryad at publication. Code that implements an offline reproduction of the central findings in this study (high-performance neural decoding of real-time attempted speech) will be made publicly available at publication.

## List of investigators and contributions

### List of investigators (authors)

1. Nicholas S. Card
2. Maitreyee Wairagkar
3. Carrina Iaccobacci
4. Xianda Hou
5. Tyler Singer-Clark
6. Francis R. Willett
7. Erin M. Kunz
8. Chaofei Fan
9. Maryam Vahdati Nia
10. Darrel R. Deo
11. Aparna Srinivasan
12. Eun Young Choi
13. Matthew F. Glasser
14. Leigh R. Hochberg
15. Jaimie M. Henderson
16. Kiarash Shahlaie
17. David M. Brandman*
18. Sergey D. Stavisky*

* These authors contributed equally

### Author contributions

N.S.C. led the experiments, implemented the real-time neural decoder and language models, designed and implemented the Copy Task and the self-initiated Conversation Mode, analyzed the data, and created the figures. N.S.C. and M.W. developed and implemented the real-time neural signal processing, noise removal, and feature extraction pipelines. N.S.C., M.W., X.H., and T.S-C. coded the real-time data collection system and built the neuroprosthetic cart system. N.S.C., M.W., and C.I. collected the primary data for this study. N.S.C. and C.I. interfaced with the participant and scheduled research sessions. E.M.K. ran hyperparameter sweeps to identify optimal hyperparameters for the neural decoder. C.F. designed and trained the language models. N.S.C. and M.V.N. created the own-voice text-to-speech models. A.S. checked for acoustic contamination and contributed to the speech intensity analyses. E.Y.C. acquired the MRI data for the HCP cortical parcellation for surgical targeting. F.R.W., D.R.D., E.Y.C, and M.F.G. processed and interpreted the HCP cortical parcellation data and provided presurgical target locations for array implantation. D.M.B and K.S. led planning and performing the surgical implant placement procedure. L.R.H. is the sponsor-investigator of the multisite clinical trial. D.M.B. was responsible for all clinical trial-related activities at U.C. Davis. N.S.C., M.W., S.D.S., and D.M.B. were involved in conceptualization of the study and experimental design. S.D.S. and D.M.B. supervised all aspects of the project. J.M.H. supervised the team members at Stanford. N.S.C., S.D.S., and D.M.B wrote the manuscript. All authors reviewed and helped edit the manuscript.

## Supplemental Text

### S1 - Experimental procedures

#### S1.01 - Study participant

This study includes data from one participant (referred to as ‘SP2’ in this preprint rather than the actual trial participant designation, which the participant is familiar with, as per medRxiv policy) who gave informed consent and was enrolled in the BrainGate2 clinical trial (identifier: NCT00912041). This pilot clinical trial was approved under an investigational device exemption by the US Food and Drug Administration (Investigational Device Exemption #G090003). Permission was also granted by the Institutional Review Boards at Mass General Brigham (#2009P000505) and the University of California, Davis (protocol #1843264). SP2 gave consent to publish photographs and videos containing his likeness. All research was performed in accordance with the relevant guidelines and regulations.

BrainGate2 is an ongoing open-label, non-blinded, multi-center, sponsor-investigator-led, feasibility study investigating the safety of chronically implanted electrodes (Utah array) in people living with paralysis. The primary endpoint of the trial quantifies the safety of the implanted device at 13 months after implantation. After meeting the primary endpoint, participants may decide to remain enrolled in the clinical trial, or to have their device explanted. Inclusion criteria include patients living with spinal cord injury, brainstem stroke, and motor neuron diseases such as ALS. Further details of the interim safety analysis have recently been reported^1^.

SP2 is a left-handed man in his 40’s with Amyotrophic Lateral Sclerosis (ALS). Four years before enrollment in the BrainGate2 clinical trial, he developed fasciculations and cramping in the legs, and was originally diagnosed with benign fasciculation syndrome. His weakness progressed resulting in recurrent falls and dropping objects. One year after initial symptoms he met El Escorial EMG criteria and was diagnosed with ALS by a neuromuscular-disease board-certified neurologist. At initial diagnosis he had mild weakness in the upper and lower extremities, and was still ambulatory with a cane. SP2 was enrolled into the clinical trial after providing informed consent. At the time of enrollment, he had no functional use of his upper and lower extremities (with the exception of 4/5 MRC-grade strength in knee extension and flexion) and had severe dysarthria, with an ALSFRS-R = 23. Since enrollment, he has maintained a modified mini-mental status exam score of 27 (the highest scores attainable). At the time of this report, he still retains eye and neck movements, but has limited orofacial movement with a mixed upper- and lower-motor neuron dysarthria resulting in a monotone, low-volume, hypernasal speech with off-target articulation. He requires non-invasive respiratory support only at night, and does not have a tracheostomy.

When speaking with non-expert listeners, he is unintelligible (Audio 1): his oral motor tasks on the Frenchay Dysarthria Assessment-2 were an “E” rating, representing non-functioning or profound impairment, and his reading of the “Grandfather Passage” was at 49 words in one minute, corresponding to severe intelligibility impairment^2,3^. When speaking to expert listeners, he communicates at 6.8 ± 5.6 (mean ± standard deviation) correct words per minute. His typing speed using his gyroscopic headmouse (Quha Zono 2), which enables him to move and click a mouse on a computer screen, is 6.3 ± 1.3 correct words per minute (Fig. S.7). The severity of dysarthria has remained stable during the time-period of this report, including the immediate postoperative period.

During the summer of 2023, four 64-electrode, 1.5 mm-length silicon microelectrode arrays coated with sputtered iridium oxide (Blackrock Microsystems, Salt lake City, UT) were placed in his left precentral gyrus. For more information on array targeting, see Sections S1.02 and S1.03, below.

#### S1.02 - Multi-modal MRI-based speech localization

Prior to microelectrode array placement, SP2 underwent a multi-modal MRI session for array targeting based on the Human Connectome Project (HCP’s) prior protocols^4,5^ and as described for BrainGate2 clinical trial participant ‘T12’^6^. SP2 was scanned in a 3T Ultra High Performance scanner (GEHealthcare) with a Nova 32-channel coil. Scan parameters were based on HCP Lifespan protocols and modified for the GE system (Table S1). Briefly, 0.8mm isotropic T1w and T2w images were acquired together with 2 mm isotropic resting state fMRI with TR = 800 ms in 4 runs each lasting 5 minutes and 45 seconds. In addition, phase reversed single band reference fMRI and spin echo MRI images geometrically and distortion matched to the MRI were acquired for distortion correction, unaliasing, and motion correction. The HCP’s minimal preprocessing pipelines^4^ were used to align the data within and across modalities, correct for image distortions, reconstruct white and pial cortical surfaces, and compute T1w/T2w myelin maps and cortical thickness maps. Subsequently, multi-run spatial Independent Components Analysis (sICA) was applied to remove spatially specific fMRI artifacts related to head motion, physiology, and the MRI scanner^7^.

These independent components were hand checked after initial automated classification using the “FIX” tool^8^ before non-aggressive regression of the artifactual components out of the fMRI data. Hand component classification was used because the application involved surgical planning. At this point the T1w/T2w myelin maps and fMRI data were used to align SP2’s brain to the HCP’s atlas space using MSMAll areal-feature-based surface registration^9^. This multi-modal cortical surface registration compensates for individual variability in areal size, shape, and position and enabled the HCP’s multi-modal cortical parcellation^4^ to be overlaid directly on SP2’s pial surface. The multi-modal surface registration was hand checked by comparing the multi-modal features computed from SP2’s brain to the same features in the atlas, with special attention paid to the features that defined the borders of areas 4, 6v, and 55b, including the T1w/T2w myelin maps (Fig. S2e) and multiple spatial ICA-based functional networks, including the language network (Fig. S2c-d), head sensori-motor network, and upper extremity sensori-motor network. Again these maps were hand checked given the neurosurgical targeting application. Once precise alignment of the HCP’s atlas of multi-modal cortical areas was confirmed, targets for the arrays were proposed, including targets in ventral area 6v, area 4, dorsal area 6v, and area 55b. These visual analyses were carried out within the HCP’s Connectome Workbench software (Fig. S2b-h).

#### S1.03 - Array placement targeting

The surgical targets for array placement within the precentral gyrus were chosen based on gross anatomical structure, vasculature, previous speech decoding studies^6,10–12^, and from estimates of cortical boundaries obtained using a cortical parcellation method derived from multi-modal Human Connectome Project (HCP) data (see Section S1.02, above). For two arrays, we targeted the dorsal and ventral aspects of area 6v due to their contributions to speech decoding in ^6^. A third array was targeted to speech primary motor cortex (area 4). We targeted area 55b for the fourth array due to emerging evidence of its importance in speech^13^.

#### S1.04 - Neural signal processing and feature extraction

Neural signals were recorded using Neuroplex-E headstages (Blackrock Microsystems) attached to two percutaneous connectors (each connected to two of the four implanted microelectrode arrays). The headstages analog filtered raw signals between 0.3 Hz to 7.5 kHz (4^th^ order Butterworth filter) and performed analog-to-digital conversion with sampling rate of 30 kHz (250 nV resolution). 1 ms windows of the digitized 30 kHz signal from 256 channels were sent to our custom BRAND node (see Section S1.05) written in Python for real-time digital filtering and feature extraction.

Each incoming 1 ms neural signal window was first band pass filtered between 250 to 5000 Hz using a 4^th^ order zero-phase non-causal Butterworth filter. 1 ms neural signal windows were padded on both sides (using the previous 1 ms window on the left side and 1.2 ms of mean padding on the right side) to minimize discontinuities at the edges. Linear Regression Referencing (LRR) was applied to each array-group of 64 electrodes to reduce noise artifacts^6,14^.

We extracted threshold crossings (putative action potentials) and spike-band power features from every 1 ms window of filtered and denoised neural signals. Threshold crossings were identified for each electrode if the voltage of the signal in this window crossed the threshold of −4.5 times the root mean squared (RMS) value of the neural signal for that electrode. Spike-band power was obtained by squaring the samples in the window and temporally averaging it for each electrode. Spike-band power was clipped at 12,500 µV^2^ to avoid outliers. This real-time signal processing, de-noising and feature extraction was performed in less than 1 ms, minimizing signal latency. These neural features were then binned into 20 ms non-overlapping bins. Binned threshold crossing counts were obtained by summing threshold crossings in 20 consecutive 1 ms neural feature windows. Binned spike-band power was computed by averaging spike-band power in 20 consecutive neural feature windows. Threshold crossings and spike-band power are commonly used measurements of local spiking (neuronal action potential) activity that have been shown to be comparable to sorted single unit activity in terms of decoding performance and neural population structure^15–17^. For brain-to-text decoding, binned threshold crossings and spike-band power from all 256 electrodes were assembled into a single 1 x 512 feature vector at every time step. Sequences of the feature vectors were smoothed and normalized before passing them into the RNN decoder (see Section S2.01).

At the start of each session, a short “diagnostic” block with attempted speech of repeated single words was recorded (see Section S1.07), used to estimate electrode-specific RMS thresholds for obtaining threshold crossing features, and LRR filter coefficients for de-noising signals as described above. Subsequently during the session, we recomputed these RMS thresholds and LLR coefficients after every block of neural data recording. Recomputing these parameters after every block helped with minimizing nonstationarities in the neural activity throughout the day.

#### S1.05 - Data collection rig

In this research, we used multiple computers to enable both rigorous scientific study of high bandwidth neural data and rapidly calibrated, real-time speech decoding with a 125,000 word vocabulary. The current form factor is larger (and has more compute capability) than is strictly necessary for the algorithms described here, in order to support rapid iteration and flexible exploration of different signal processing and decoding methods. A future, final-form medical device would reduce the current setup to an embedded system with a small, portable external component. We direct readers to a previous Utah arrays-based reach-and-grasp brain-computer interface study as an example of how a similar research data collection rig was made substantially more compact and portable^18^.

All real-time data collection, processing, analysis, and decoding was performed by a group of four computers connected in a local area network. A computer running Windows 10 interfaced with the Neuroplex-E system to start and stop neural data recording. A second computer (running Ubuntu 22.04 LTS) was used to process and extract neural features from raw 30 kHz neural data. A third computer (running Ubuntu 22.04 LTS) was responsible for real time neural decoding, fine-tuning the RNN model online, displaying the task to the participant, and displaying the task control GUI on the research team-facing monitor. Finally, a fourth computer (running Ubuntu 22.04 LTS) was used to run the language model that converted phoneme sequences to words. We used the Backend for Realtime Asynchronous Neural Decoding (BRAND^19^) to run our data collection computer setup. All code was written in Python, C, or MATLAB.

#### S1.06 - Overview of data collection sessions

Neural data were recorded in 5-7 hour long research sessions, which took place at the participant’s home 2-4 days per week. SP2 chose the dates and times for sessions to occur. Sessions typically included 1-2 breaks for food or beverages. During the sessions, SP2 sat in his power lift chair in an upright position. A computer monitor placed in front of SP2 displayed the task. An eye tracker mounted to the bottom of the computer monitor allowed SP2 to select on-screen “buttons” by looking at them. Data was collected in 15-25 minute “blocks” consisting of an uninterrupted series of trials. Trials could be paused as necessary and continued, or terminated, as appropriate. Between blocks, SP2 was encouraged to rest as needed. Table S2 lists all data collection sessions reported in this study. Across all sessions, we collected an average of 236 minutes of neural data. Microphone data was recorded at 30 kHz during all sessions and synchronized with neural data via the Neural Signal Processor’s analog in port. Video was recorded for all sessions from two cameras set up behind and in front of the participant.

In keeping with historical precedent in our clinical trial, we began data collection 25 days after implantation^1^. While a delay between surgery and device initialization is standard practice in clinically approved neuromodulation procedures such as deep brain stimulation and vagal nerve stimulation, in principle data collection could have begun within hours or days after implantation. Building on our previous demonstration of rapid point-and-click communication with first-time BCI users^20^, we began closed-loop speech neuroprosthesis evaluation on the first day when the participant was connected to the recording system (session 1).

#### S1.07 - Instructed delay Copy Task

In an instructed-delay Copy Task (Videos 1-2), a prompted sentence was displayed as text on a computer monitor facing SP2. A colored square changed from red to green to indicate when he should begin speaking. SP2 triggered the end of each sentence using an on-screen eye-tracker “button”, at which time the final decoded sentence was read aloud with a text-to-speech algorithm that was customized to sound like the participant’s pre-ALS voice^21^ (Section S5). To support future neuroprosthesis users incapable of eye gaze control, we also demonstrated sentence finalization triggered by neural decoding of SP2’s attempted hand squeezes (Section S6). The majority of Copy Task blocks were 50 trials long, which took 15-25 minutes depending on how long the prompted sentences were. SP2 controlled the pace of each Copy Task trial via either eye tracking (Section S1.11) or gesture decoding (Section S6). In early sessions, prior to eye tracking implementation, SP2 signaled to the researcher that he was done speaking at the end of each trial by making eye contact with the researcher, who would press a button to end the trial.

At the start of each session, we did a “diagnostic block”, which was an instructed delay task with 8 single-word cues each repeated 6-8 times. The word set consisted of the words ‘bah’, ‘choice’, ‘day’, ‘kite’, ‘though’, ‘veto’, ‘were’, and a ‘DO NOTHING’ condition where SP2 was instructed not to say or do anything, consistent with ^6^. Data from this block was used to calculate initial thresholds and weights for linear regression referencing, which were then updated after each subsequent block.

#### S1.08 - Conversation Mode

In Conversation Mode (Videos 3, 4, and 5), no prompted sentences were shown on screen. Instead, SP2 could say whatever he wanted. Video 2 shows the first ever self-directed use of the speech neuroprosthesis in Session 2, where he spoke to his daughter (Fig. 3b). In this first self-directed usage, speech detection had not been implemented yet, so SP2 waited until the square turned from red to green (like in the Copy Task) and then began to say whatever he wanted to.

In subsequent sessions, we developed a dedicated Conversation Mode that detected when SP2 spoke and decoded accordingly. In Conversation Mode, SP2 would initiate a new sentence by simply attempting to speak, which the speech neuroprosthesis would reliably detect using only neural data (Fig. S22). To accomplish this detection, the RNN decoder was always running in the background to predict phoneme probabilities every 80 ms. These phoneme probabilities were analyzed in real time to detect when speech had started (as indicated by the probability of any phoneme being higher than the probability of silence) or ended (if the probability of silence was higher than the probability of any phoneme for 6 consecutive seconds; this duration was determined over the first few sessions of this task to balance accidentally timing out a sentence early, with not making SP2 wait too long for it to end when he wanted it to). SP2 could end a sentence using the eye tracker (by dwelling his gaze over an on-screen button that said “DONE”) or by waiting six seconds for the trial to time out, after which time the final sentence was read aloud by the TTS algorithm. SP2 used the eye tracker to confirm whether the final decoded sentence was correct, or if not, he could specify whether it was “mostly correct” or “incorrect”. We also included a “spelling mode”, toggled by an on-screen eye tracker button, which allowed SP2 to spell out individual words by attempting to say each individual letter (e.g., “h”, “e”, “l”, “l”, “o”). Spelling mode leveraged a separate language model that had an output space limited to the 26 English letters, and was biased towards (but not limited) predicting letter sequences that formed valid English words. Sentences and letter sequences that were confirmed to be correct were used to fine-tune the RNN in the background (Section S2.03), which helped decoding performance remain stable and accurate throughout usage of the speech neuroprosthesis.

A major upgrade to Conversation Mode was implemented in session 72 (see Video 5). This upgrade included two major additional features: (1) SP2 could now control if and when the text-to-speech audio was played after each sentence by looking at an on-screen eye tracker button. Previously, every sentence was automatically read aloud upon finalization. This functionality helped SP2 to better fit his sentences into the flow of conversation, and also enabled him to play audio for sentences multiple times, either to repeat himself to get someone’s attention, or for comedic effect. (2) Upon SP2’s marking a final decoded sentence as “mostly correct”, the speech neuroprosthesis would now offer him up to 5 additional candidate sentences to choose from, which were the next five most likely decoded sentences. In each candidate sentence, words that differed from the original final decoded sentence were highlighted in yellow to make the differences easier to see. This functionality enabled SP2 to communicate more quickly by arriving at the correct decoded sentence without having to repeat himself.

The duration of personal use blocks ranged from approximately five minutes to over 7 hours. The user interface design of the self-paced Conversation Mode was iteratively improved in response to feedback from SP2 over the course of the study (e.g., see Table S4). SP2 used the speech neuroprosthesis in Conversation Mode for over 248 cumulative hours to speak with the research team, with family and friends, or with work colleagues. Conversations took place in-person, over phone and video calls, and over text messages and emails. In session 36 we added the (optional) ability for the speech neuroprosthesis to type final decoded sentences onto SP2’s personal computer by acting as a bluetooth keyboard. This functionality enabled SP2 to independently send text messages, emails, and perform other applications on his personal computer that involved text entry.

#### S1.09 - Decoder evaluation

To evaluate speech decoding performance, we computed phoneme error rate and word error rate using Levenshtein distance, which counts the number insertions, deletions, or substitutions necessary to match the decoded phonemes or words to the ground truth labels. For assessing the RNN output (without language models), we calculate the “raw phoneme error rate” by comparing the most probable phoneme decoded in each time step (duplicates removed) with the ground truth phoneme sequence. Consistent with ^6^, reported error rates were aggregated across all evaluation sentences from each session by summing the number of errors (insertions, deletions, or substitutions) for all sentences and then dividing it by the total number of words in those sentences. This helps prevent very short sentences from overly influencing the result. Confidence intervals for error rates were computed via bootstrap resampling over individual trials and then re-calculating the aggregate error rates over the resampled distribution (10,000 resamples).

Blocks where the participant was excessively tired, per his own report, were excluded from evaluation (2 of 41 total blocks); the word error rates on these blocks were 8.3% (session 14) and 5.3% (session 15). In each session, we collected 1-5 evaluation blocks (50-250 sentences). The number of evaluation blocks collected on any given day was dependent on the variable amount of time available in each session based on the participant’s schedule.

Before every session, an RNN was trained (Section S2.02) on all previous data. In early sessions (1-11), an additional new model was also trained halfway through data collection to calibrate it to the current day. From session 12 onward, after online fine-tuning was introduced, we stopped training a new model halfway through the day and instead relied on the online fine-tuning (Section S2.03).

We note that evaluating performance of Conversation Mode blocks has some inherent limitations. First, since some of the sentences may have been aborted early or be so incorrect as to be unintelligible, an estimate of the ground truth of these sentences would be impossible to determine. Second, aside from the session where we verified the content of all attempted sentences by asking the participant to identify ground truth labels for incorrectly decoded sentences, we used a combination of directly asking the participant, and examining the predicted phoneme patterns and the top 100 candidate sentences for each utterance in light of the context of the conversation; the latter approach involves some subjectivity.

#### S1.10 - Sentence selection

For 50-word vocabulary decoding (sessions 1 and 2), custom-written prompted sentences contained words from a 50-word vocabulary^10^. For 125,000-word vocabulary decoding (sessions 2 onward), sentences were sourced from the Switchboard corpus^22^, as in ^6^. We chose to use the same vocabularies as these two prior studies to facilitate comparison of speech decoding performance. Additional training sentences were sourced from the OpenWebText2 corpus^23^ and the Harvard Sentences^24^ in an effort to expand the sampled vocabulary and thus the decoder’s ability to generalize (Fig. S18, S17). Sentences were manually screened for grammatical errors or offensive language. We collected data for 10,481 prompted sentences over 50 sessions, totaling 67.4 hours of neural recording.

#### S1.11 - Eye tracking

SP2’s gaze data were tracked using a Tobii Pro Spark eye tracker (Tobii AB, Stockholm, Sweden). Eye tracker calibration was performed at the beginning of each session, and repeated as necessary between data collection blocks. During data collection blocks, SP2’s on-screen gaze location was recorded at 60 Hz and used by the speech neuroprosthesis to allow him to select on-screen “buttons” by looking at them for 0.5 seconds. Gaze data was recorded independently from each eye before being averaged and smoothed over time. All eye tracker calibration, gaze data recording, and logic for on-screen button selection was done with custom written Python code that was integrated into our BRAND-based^19^ data collection rig.

#### S1.12 - Estimating electrode array placement in MNI space

We estimated the location of the four Utah arrays in the precentral gyrus using the asymmetric MNI2009b atlas. This was performed as follows. First, by corroborating intra-operative photographs with a post-operative CT scan co-registered to the pre-operative anatomical T1 MRI, we identified the center-point of each array on SP2’s precentral gyrus. Second, we aligned SP2’s brain to ACPC coordinate space, and computed the ACPC coordinate of each array. Third, we used the Advanced Normalization Tools package^25^, to “skull strip” the MRI (using the asymmetric MNI2009b atlas as a template), and then applied antsRegistration to perform non-linear deformation from SP2’s anatomical T1 to the MNI2009b template^26,27^. Finally, we applied the warping composite deformation of the ACPC coordinates of the arrays.

This approach yielded the following coordinates:

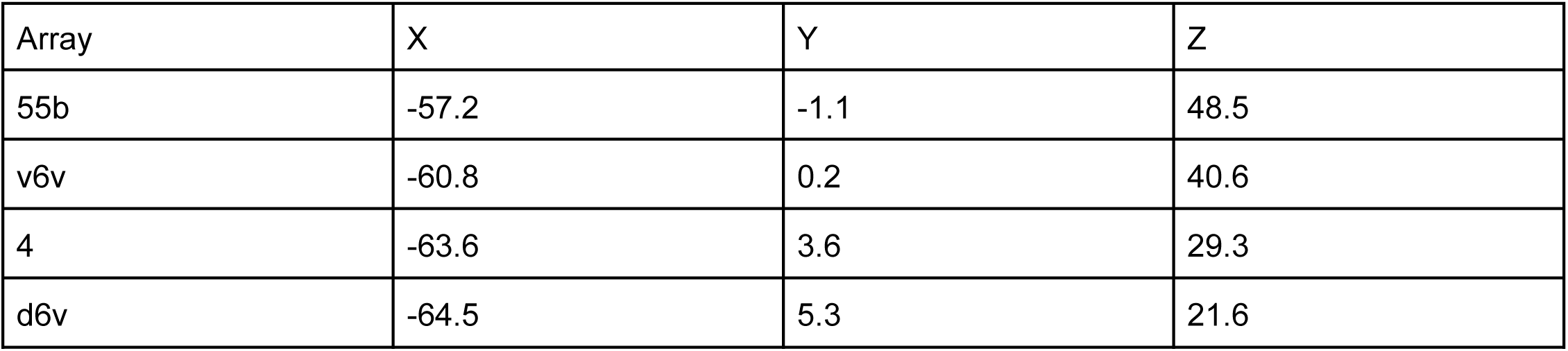

| Array | X | Y | Z |
| --- | --- | --- | --- |
| 55b | -57.2 | -1.1 | 48.5 |
| v6v | -60.8 | 0.2 | 40.6 |
| 4 | -63.6 | 3.6 | 29.3 |
| d6v | -64.5 | 5.3 | 21.6 |

We note that this approach to estimating corresponding array location in MNI space is highly dependent on parameters chosen at each step of the ANTs registration. The “skull stripping” procedure used the default parameters provided in the ANTs registration package of version 2.4.0. Motivated by author D.M.B.’s experience in performing nonlinear deformations of template atlases for post-operative analysis of stereotactic procedures, the following parameters were chosen, implemented using the open-source *nipype* package (version 1.8.3):

Transforms = [’Rigid’, ‘Affine’,’SyN’]
Transform_parameters = [(0.1,), (0.15,), (0.3,3,0)]
Convergence_window_size = [10,10,10]
Shrink_factors = [[12,8,4,1], [8,4,2,1], [8,6,4,2,1]]
Convergence_threshold = [1e-6, 1e-6, 1e-6]
Sigma_units = [’vox’, ‘vox’,’vox’]
Metric = [’MI’,’MI’,’MI’]
Smoothing_sigmas = [[4,3,2,1],[4,3,2,1],[4,3,2,1,1]]
Number_of_iterations = [[1000,500,250,0],[100,500,250,0],[200]*5]
Radius_or_number_of_bins = [32, 32, 32]
Sampling_strategy = [’Regular’, ‘Regular’,’Regular’]
Sampling_percentage = [0.25, 0.25, 0.25]

### S2 - RNN decoder

#### S2.01 - RNN architecture and feature preprocessing

A recurrent neural network was used in this study to predict sequences of phoneme probabilities from neural data. In brief, the RNN consisted of (1) linear day-specific input layers to correct for nonstationarity in neural data between days, (2) 5 layers of gated recurrent unit (GRU) architecture with 512 units per layer, and (3) a dense output layer that outputted the probabilities of 41 classes (39 phonemes, silence, and a CTC blank token). The RNN ran every 4 bins (20 ms per bin) to predict phoneme probabilities from the most recent 14 bins of neural data (280 ms). Thus, a set of probabilities of each phoneme being spoken were generated by the RNN every 80 ms.

Before neural features were input into the RNN, they were z-scored using their means and standard deviations from the speech epochs of the previous 20 trials, and then smoothed with a Gaussian kernel (s.d. = 40 ms) that was delayed by 160 ms. The smoothing step reduced noise in the data, but also added 160 ms of latency to the neuroprosthesis, which was not consequential for the brain-to-text decoding described here, but may negatively affect performance in other decoder paradigms where minimizing feedback latency is important. Connectionist temporal classification (CTC) loss was used to output a sequence of predicted labels (phonemes) from an unlabeled time sequence of neural data. For additional details about the RNN architecture, refer to the supplemental methods section of ^6^. All RNN parameters are listed in Table S5. We note that the hyperparameters chosen here were optimized for decoding SP2’s attempted speech (Fig. S9) and may need to be individually tuned to future users.

#### S2.02 - Offline RNN training

The offline RNN training protocol used here is the same as in ^6^. A new RNN was trained before each session, and also mid-session for the first 11 sessions (before online fine-tuning was introduced). Whenever an offline RNN was trained, 90% of all previous data were used for training, and 10% of data were randomly (uniformly from each session) held-out for validation. Prompted sentences from each trial were converted to a sequence of phonemes using the Python *g2p-en* package ^28^. The RNN was trained with neural feature sequences and target phoneme sequences for 2,000-50,000 batches (the number of batches was increased as the training data pool grew throughout data collection), and the learning rate was linearly decayed from 0.02 to 0.0 across all batches. In each batch, up to 64 trials of data from a randomly selected session were input into the corresponding day-specific input layer followed by the GRU and dense layers. Neural data was dynamically augmented on a batch-by-batch basis during training to improve decoder generalizability and stability by adding (1) white noise and (2) artificial constant offsets to the neural features^29^. At the end of each batch, weights for the relevant day-specific input layer, GRU layers, and dense output layer were updated using stochastic gradient descent (ADAM; *β*_1_ = 0.9, *β*_2_ = 0.999, *ε* = 0.1). We applied dropout and L2 weight regularization during training to improve generalization. Parameters used for online speech decoding are listed in Table S5.

#### S2.03 - Online RNN fine-tuning

Online RNN fine-tuning was introduced in session 12 in an effort to frequently adjust the RNN to shifts in neural signals to ensure that speech decoding performance remained consistently high throughout a session. The online fine-tuning method employed here is similar to the one introduced in ^30^ for a handwriting decoder, but was adapted here to work with speech decoding tasks. At the beginning of a new session, an RNN trained on all previous data was loaded, and the day-specific input layer corresponding to the most recent session was duplicated. After each trial in the new session (starting after 10 trials of data had accumulated in sessions 12-25, and 5 thereafter]), the weights of this day-specific input layer and the weights of the base GRU model were updated using the neural data and ground-truth sentences from each trial. Ground-truth sentences were defined as either the prompted sentence (in the Copy Task) or as decoded sentences that SP2 confirmed to be correct (using the eye tracker) in the self-initiated Conversation Mode. During each fine-tuning epoch, data from previous sessions was randomly sampled in a proportion of 60% new data (from the current session) to 40% old data (from previous sessions) when training the model in an effort to ensure that the model did not overfit to the current day’s data. A static learning rate of 0.04 was used to fine-tune the RNN throughout the session. For additional details about online RNN fine-tuning, refer to ^30^. Parameters used for online speech decoding are listed in Table S5.

#### S2.04 - Hyperparameter optimization

Optimal hyperparameters for RNN architecture and training were determined with hyperparameter sweeps twice throughout data collection (Fig. S21), and optimal parameters were used in subsequent online decoding sessions. RNNs were trained offline (using all previous data) with one hyperparameter varied at a time. Each RNN was validated on randomly-selected held-out validation trials to evaluate performance (raw phoneme error rate). For each parameter condition, 10 RNNs were trained and their validation performances were averaged.

We note that all data used to train the hyperparameters for the RNN were exclusively from participant SP2. An open question remains whether the hyperparameters from one participant could be used to seed a generic RNN, to be used with a future participant.

### S3 - Language model

#### S3.01 - Architecture

The *n*-gram language models in this study take sequences of phoneme probabilities as an input and output the most likely sequence of words. The 50-word language model used here was a 5-gram model trained on 2,413 custom-written sentences that contained only words from the 50 word vocabulary^10^. This model outputs only the singular most likely sequence of words. The 125,000-word language model, trained on the OpenWebText2 corpus^23^, was the same 5-gram model described for post-hoc offline analyses in ^6^. The 125,000-word vocabulary included in this language model stems from the CMU dictionary (http://www.speech.cs.cmu.edu/cgi-bin/cmudict) and encompasses the majority of the English language; native English speakers typically know ∼20,000 to 40,000 words^31,32^. We added a list of names provided by the participant (e.g., those of his family members and the research team) to the output vocabulary space of the language model, enabling him to say those names using the speech neuroprosthesis. This language model initially predicts up to 100 of the most likely sequences of words, before rescoring them in multiple stages to identify the singular most likely sequence of words. To use this language model online and in real time, we implemented custom-written Python code to integrate it into our real-time decoding system. For additional details about the language models utilized in this study, refer to the supplemental methods section of ^6^. Parameters used for online speech decoding are listed in Table S5.

#### S3.02 - Hyperparameter optimization

As we collected data, we ran offline language model hyperparameter sweeps to identify optimal speech decoding parameters (Fig. S21). In particular, the *acoustic scale* and *alpha* parameters had the biggest impact on decoding performance. The *acoustic scale* parameter is the weighting ratio between the RNN-derived phoneme probabilities and the n-gram derived sentence probabilities, and the *alpha* parameter is the weight ratio between the *n*-gram model rescoring and the OPT LLM rescoring (see supplemental methods in ^6^ for more details). After hyperparameter tuning, we used these identified optimal hyperparameters to enhance speech decoding accuracy online in subsequent speech decoding sessions.

#### S3.03 - Automatic sentence punctuation

Starting in session 38, the final decoded sentence was automatically punctuated by passing the predicted word sequence text through a publicly available open-source auto punctuation model (“fullstop-punctuation-multilang-large”)^33^. This model is capable of quickly predicting the punctuation of English, Italian, French and German texts. Punctuation was not always correct, and the participant was instructed not to consider punctuation in his assessment of correctness when using the speech neuroprosthesis in Conversation Mode. The participant reported being largely pleased with this auto-punctuation implementation, and the punctuation also helped to add appropriate pauses to audio generated by the text-to-speech model.

### S4 - Offline analyses

#### S4.01 - Offline RNN analyses

Offline RNN decoding analyses were utilized in Figures S6, S10, S11, S13, S14, S15, S19, and S20 of this study. All analyses averaged the results from 5-10 RNN seeds per condition. Chance decoding values (reported in Fig. S16c) were calculated by training a decoder where the phonemes of the ground-truth sentence for each trial were shuffled. This allowed us to maintain the statistical distribution of uttered phonemes while obtaining a chance decoding value. For each electrode count condition of the electrode dropping analysis (Fig. S16b), random electrodes were sampled uniformly from all arrays, and data from unused electrodes was zeroed out. Unless specified, RNN architecture and hyperparameters remained consistent between all conditions.

#### S4.02 - Offline language model analyses

Offline language model performance analyses were performed for Figures S12, S15, and S21. In these offline analyses, RNN-predicted phoneme sequences, either from online speech decoding sessions or from offline RNN analyses (see Section S4.01), were sequentially fed into language models for each trial of data, before finalization. Offline language models were initialized with a range of parameters or vocabularies as relevant for the analysis. Language model performance was assessed as the aggregate word error rate of all predicted sentences, calculated as described in Section S1.09.

#### S4.03 - Acoustic contamination analysis

We assessed whether the neural data was contaminated by the audio vibrations associated with attempted speech due to a potential microphonic effect at one or more elements of the electrophysiology recording chain^34^. We designed a Speech Amplitude task where SP2 was instructed to say single words (“be”, “go”, “my”, “know”, “have”, and “going”) at a specified amplitude ( “SILENT”, “WHISPER”, “NORMAL”, and “LOUD”). In offline analyses, we then performed the same analysis as described in ^34^ to determine if the neural data was potentially acoustically contaminated.

The Speech Amplitude task was designed similarly to the Copy Task described in Section S1.07. Here, a prompt consisting of a word and the required amplitude at which the word needs to be spoken was displayed as text (for example, “LOUD: be”) on a screen facing SP2. A red square on the screen turned to green and a beep sound played over a speaker, prompting SP2 to attempt to speak. Each trial had a fixed duration of 3 seconds. The words used in this task were selected because they are some of the most frequently sampled words in the Switchboard dataset. For the “SILENT” amplitude, SP2 was instructed to attempt to speak absolutely silently, as if he was mouthing a word to someone across a room. For the “NORMAL” amplitude, SP2 was instructed to maintain the same speech amplitude level as used during the Copy Task (Section 1.07). We also included a “DO NOTHING” prompt, where SP2 was instructed to not say or do anything.

In each research block, the 25 prompts (6 words × 4 amplitudes + “DO NOTHING”) were repeated 5 times each in a random order. Each block took approximately 10 minutes. In total, we collected 5 blocks of data, resulting in 625 total trials. Two trials were excluded because SP2 was coughing during them.

We used the methods and code described in ^34^ to check each trial from the speech amplitude task for acoustic contamination. Neural data from 585/623 trials (93.9%) did not have evidence of acoustic contamination (p > 0.05; Fig. S7a). Neural data from the remaining 38/623 trials (6.1%) were flagged as potentially acoustically contaminated using a statistical threshold of p < 0.05. These trials with significant correlation between the microphone recording and multielectrode array voltages included 4 “DO NOTHING” trials, 6 “LOUD” trials, 9 “SILENT’’ trials, 10 “NORMAL” trials and 9 “WHISPER” trials. The flagged trials were distributed across the different speech amplitudes and, given that we have chosen alpha to be 0.05, we expect ∼5% of the trials will be flagged as potentially contaminated at random due to chance under the null hypothesis that there is no acoustic contamination. Thus, we consider the 6.1% of all trials that were flagged as potentially acoustically contaminated to be at the chance level, and conclude that there is no evidence that the neural data presented in this study was acoustically contaminated. This is supported by the finding that 9/149 trials during the “SILENT” condition showed potential acoustic contamination despite the complete absence of vocalized speech.

#### S4.04 - Decoding accuracy during attempted silent, whispered, or loud speech

In both offline and online analyses, we aimed to determine if the speech neuroprosthesis could generalize to other amplitudes of attempted speech, including attempted silent, whispered, or loud speech.

First, we used a decoder trained on speech attempted at a normal volume to predict (offline) what SP2 was attempting to say during the Speech Amplitude task described in Section S4.03. We found that the decoder generalized reasonably well to the other speech amplitudes (Fig. S19a), which could be decoded with much better accuracy than would be expected by chance (∼80%; Fig. S16a). It is not surprising that we observed the lowest phoneme error rate for the “normal” speech volume condition, considering that the decoder was trained solely on attempted speech using the same strategy.

We hypothesized that including more training data where SP2 speaks silently may help the decoder adapt to that condition and more accurately predict the intended words. To this end, we dedicated an entire session and a half to calibrating the speech neuroprosthesis with silent speech training data, and then evaluating how accurately it could predict silent speech online. We found that we could decode silent speech online with word error rates below 15%, and as low as below 5% (Fig. S19b). As predicted, word error rates started higher in early training blocks during each session and tended to be lower in subsequent blocks as the neuroprosthesis calibrated itself. These results show that silent speech could be decoded online with accuracy exceeding previous speech decoding accuracies^6,12^ but lower than SP2’s attempted normal amplitude speech performance. We predict that collecting additional silent speech data and optimizing the RNN and LM hyperparameters on that speech modality could further increase decoding accuracy, which remains an avenue for further investigation.

### S5 - Own-voice text-to-speech

#### S5.01 - Fine-tuned VITS text-to-speech model

We trained a text-to-speech algorithm^21^ to sound like SP2’s pre-ALS voice. SP2 and his family provided us with home videos and other recordings of SP2 speaking. Recordings with clear samples of SP2’s voice were segmented into individual sentences, noise-reduced (RNNoise 1.4^35^), and amplitude-normalized in preparation for training the TTS model. Signal-to-noise ratio (SNR; calculated with Waveform Amplitude Distribution Analysis (WADA) with the Coqui TTS Check-DatasetSNR notebook^21^) was used to quantify how noisy each audio clip was. Each audio clip was then manually transcribed, and the phoneme distribution across all audio clips was calculated (using the Python *g2p-en* package^28^) to ensure that each phoneme was adequately sampled. Comprehensive coverage of each phoneme is required to train robust text-to-speech models and accurately reproduce speech patterns.

For training an own-voice TTS, we chose the VITS model^36^, which we subjectively found to reproduce SP2’s pre-ALS voice most accurately and without a long delay at the end of each sentence. A pre-trained VITS model with LJSpeech corpus was fine-tuned using SP2’s processed audio samples to create a TTS that sounded like SP2’s pre-ALS voice. This TTS was used to read the final decoded sentence at the end of each trial in both the Copy Task and Conversation Mode. SP2 and his family reported that they found our recreation of his pre-ALS voice to be more representative of his pre-ALS voice than his previously purchased commercial version.

#### S5.02 - Fine-tuned StyleTTS2 text-to-speech model

In session 55 we deployed a significantly upgraded own-voice TTS model which was based on StyleTTS2, which leverages style diffusion and adversarial training with large speech language models to achieve human-level TTS synthesis^37^. Using the same training dataset described in Section S5.01, plus additional audio clips acquired from SP2 since training the original own voice TTS model, we followed the instructions available on the StyleTTS2 GitHub repo to fine-tune a pre-trained StyleTTS2 model to sound like SP2’s pre-ALS voice. The fine-tuned StyleTTS2 own-voice TTS model sounded much more naturalistic and less robotic than the previously described VITS own-voice TTS model. SP2 and several of his family members and friends independently agreed that the StyleTTS2 own-voice TTS model closely resembled SP2’s pre-ALS voice. The first time that SP2 used the new TTS with the speech neuroprosthesis, he was emotionally moved and used the speech neuroprosthesis to repeatedly tell the research team and his family: “I am so f*****g back.” (The censored-for-publication word was decoded and synthesized aloud with correct spelling and pronunciation.)

### S6 - Gesture decoding for task control

People with ALS may lose precise and reliable eye gaze control as their disease progresses. Thus, eliminating reliance on eye gaze (e.g., by making the system controllable through exclusively neural signals) is necessary to ensure that the neuroprosthesis will remain usable in the long-term for users with degenerative diseases such as ALS. We provided SP2 with a “neural click” functionality as an alternative to eye tracker control for indicating that he is done speaking, which triggers sentence finalization and text-to-speech output. This neural click functionality was tested during closed-loop speech neuroprosthesis use in sessions 17 and 18 (275 total sentences).

#### S6.01 - Motor imagery

We chose “right-hand squeeze” as the motor imagery to perform the neural click, because it had a robust neural signal to noise ratio in previously collected SP2 movement sweep data, and the hand squeeze gesture has been used for neural click in prior intracortical brain-computer interface studies^38,39^. Other discrete gestures may have worked just as well for this purpose^40^.

#### S6.02 - Decoder architecture

We implemented a linear gesture decoder (independent from the RNN speech decoder) to solve the binary classification problem: *“For each 10 ms bin, is the user attempting right-hand squeeze or not?”*. We chose to use a linear discriminant analysis (LDA) model, because linear models are simple and fast to train, and in this case were able to reliably distinguish the neural correlates of hand squeezes from those of speech or silence in preceding offline tests.

#### S6.03 - Decoder training

We interspersed 16 trials of a “*RIGHT HAND - CLOSE”* condition into the instructed delay task (our “diagnostic block”) performed at the start of each session. After the diagnostic block, we trained the LDA classifier on all the trials (“*RIGHT HAND - CLOSE”* = click, single-word speech trials = non-click, “*DO NOTHING”* = non-click). The training data from each trial came from the epoch 0.5 - 1.5 seconds after the go cue. Each trial yielded 100 training samples, as our LDA classifier operated on individual 10 ms time bins.

Each training sample was a single time bin’s feature vector (256 threshold crossings + 256 spike band power = 512 features). These feature vectors were processed identically to those used for speech decoding (filtered, z-scored, etc.). To fit the LDA model on these training data, we used the *LinearDiscriminantAnalysis* class from the Python package *sklearn*^41^.

#### S6.04 - Decoder inference

After training the LDA classifier, we used it during closed-loop speech blocks to decode neural clicks in real-time. Because this LDA neural click decoder was independent from the RNN speech decoder, it was run in parallel using the BRAND software architecture. Though the LDA model outputted a prediction for every 10 ms time bin, a click was not immediately performed every time the LDA model predicted click. Instead, a click was only performed when all time bins in a 100 ms sliding window were predicted as click, to reduce spurious clicks. Additionally, after each click we maintained a refractory period of 1 second during which no additional clicks were performed, to avoid erroneous repeated clicking.

### S7 - Trial Registration

Clinicaltrials.gov: NCT00912041

<u>Date registered:</u> June 2009

<u>Date first patient enrolled in study:</u> May 2009*

<u>Number of patients enrolled before registration:</u> 1*

<u>Reason for delay:</u> *No new patients were enrolled before registration. The “first” participant in this study was a transitional participant who was previously enrolled in another clinical trial. She was technically transitioned from that study to this study in May 2009, which was the date the Investigational Device Exemption (IDE) for this study was granted. The IDE at that time preceded the clinicaltrials.gov registration (June 2009); we’ve consistently used the May date to mark the enrollment of this transitional participant, as this is when all regulatory permissions required had been granted for her to transition to the current trial.

## Supplemental Figures

**Figure S1:**
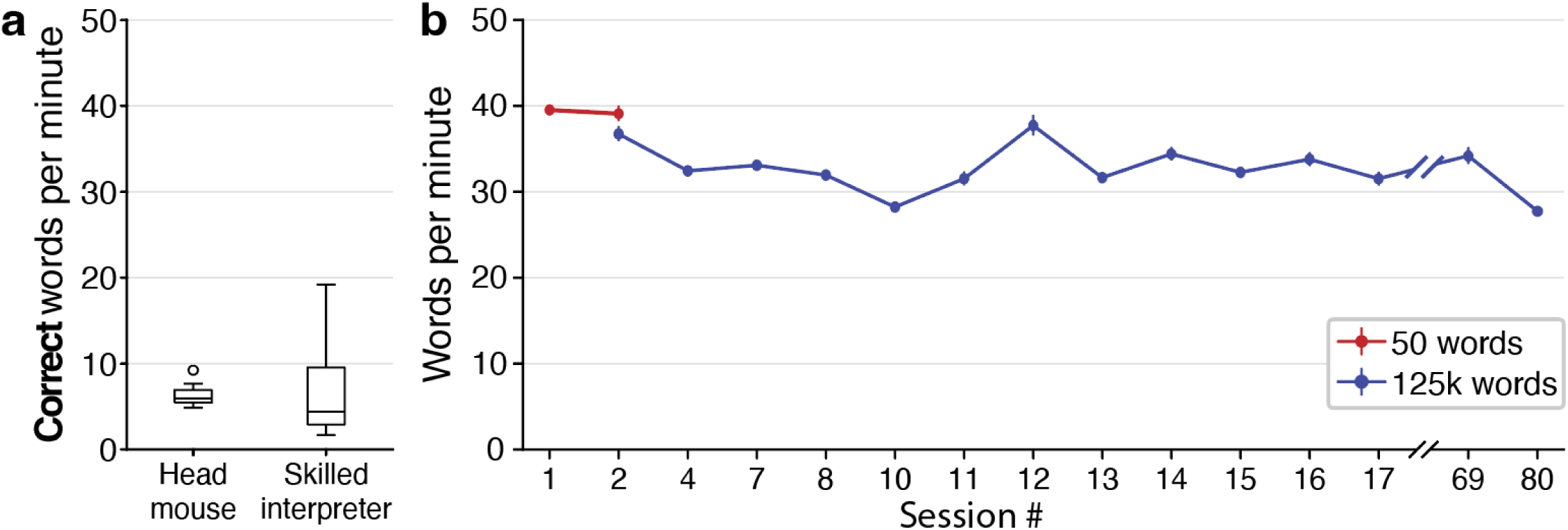
Communication rates with and without the speech neuroprosthesis. **a,** The participant’s correct words per minute communicated using either a gyroscopic head mouse (left) or when attempting to speak to a skilled interpreter (right). Communication rates were calculated for each modality based on ten switchboard sentences. For the head mouse modality, SP2 wore a gyroscopic device over his right ear that allowed him to move a computer cursor along an on-screen keyboard by tilting his head in each direction. The on-screen keyboard showed standard keyboard keys in addition to auto-complete word predictions, which SP2 took advantage of when the correct word was available. For the skilled interpreter modality, SP2 attempted to say prompted sentences, one word at a time, to a skilled interpreter. When the interpreter struggled to correctly identify a word, SP2 used other strategies such as spelling the word out, or providing additional context to the interpreter. **b,** SP2’s average rate of attempted speech during evaluation blocks for each session. Error bars denote the 95% confidence interval. For each sentence, words per minute was calculated as the number of words in the target sentence divided by the duration from the beginning of the first word until SP2 signaled the end of the sentence (using the eye tracker or gesture decoder).

**Figure S2:**
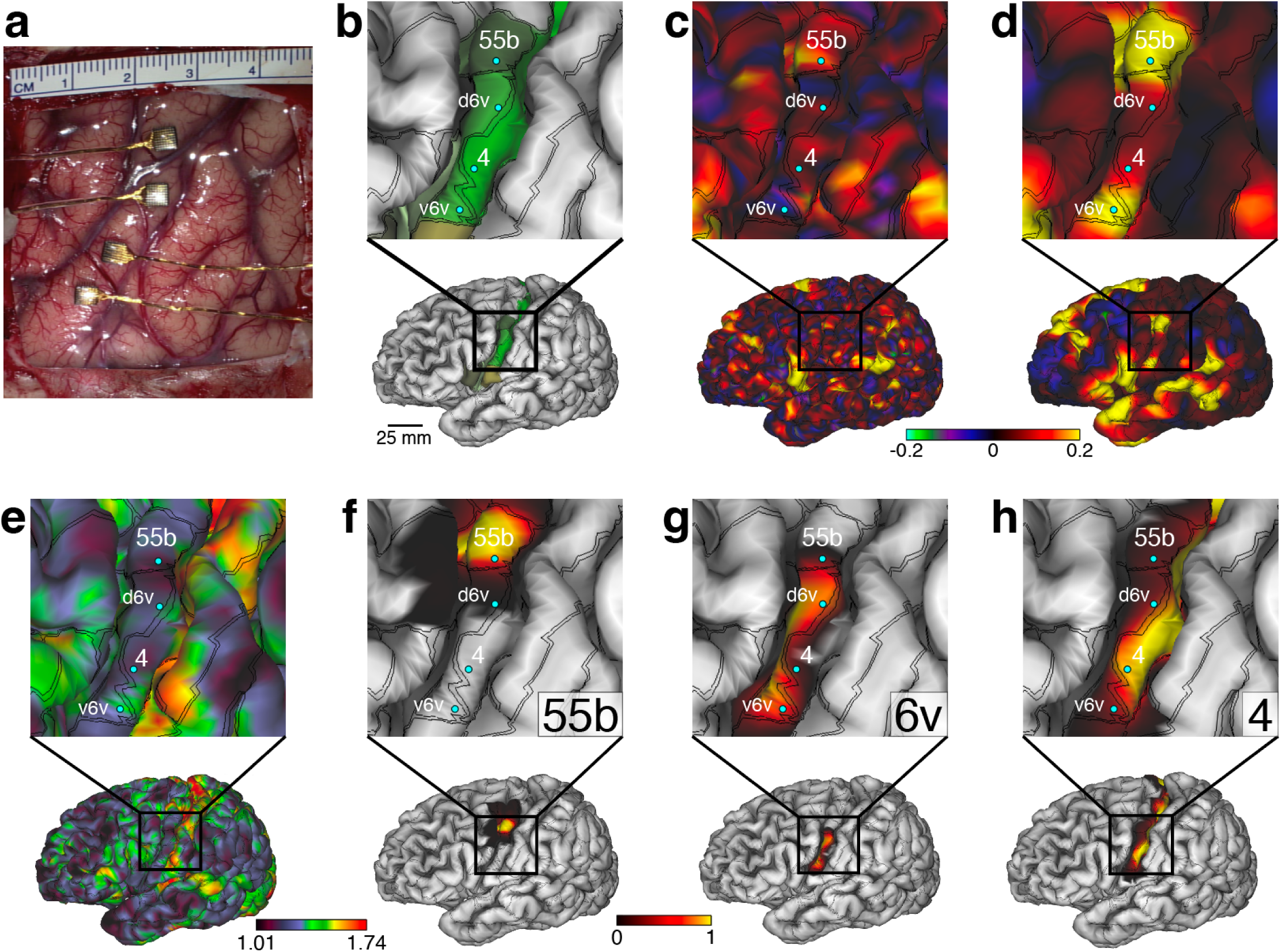
Multi-modal MRI-based speech localization and array targeting. **a,** Array implants shown on the surface of SP2’s brain during surgery. **b,** Approximate array locations on SP2’s inflated brain using Connectome Workbench software, overlaid on the cortical areal boundaries (double black lines) estimated by the Human Connectome Project (HCP) cortical parcellation. **c**, Approximate array locations overlaid on a language-related resting state network shown for SP2’s individual scan. **d**, The same resting state network identified in the Human Connectome Project data (i.e., averaged across many subjects) and aligned to SP2’s brain. **e**, Approximate array locations overlaid on a myelin density map. **f-h**, Approximate array locations overlaid on the confidence maps of the areal region labeled in the bottom right of the magnified panel.

**Figure S3:**
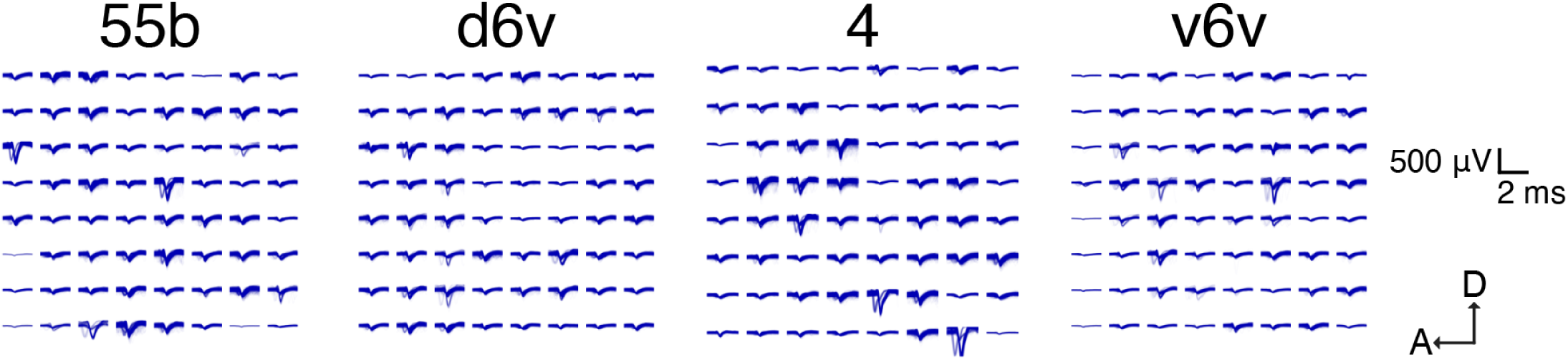
Action potential waveforms from the four microelectrode arrays. Representative neuron action potential waveforms from a 60-second segment of attempted speech during the Copy Task. The waveforms show a 1 ms period around the −4.5 RMS threshold crossings. Neural data was processed as described in section S1.04.

**Figure S4:**
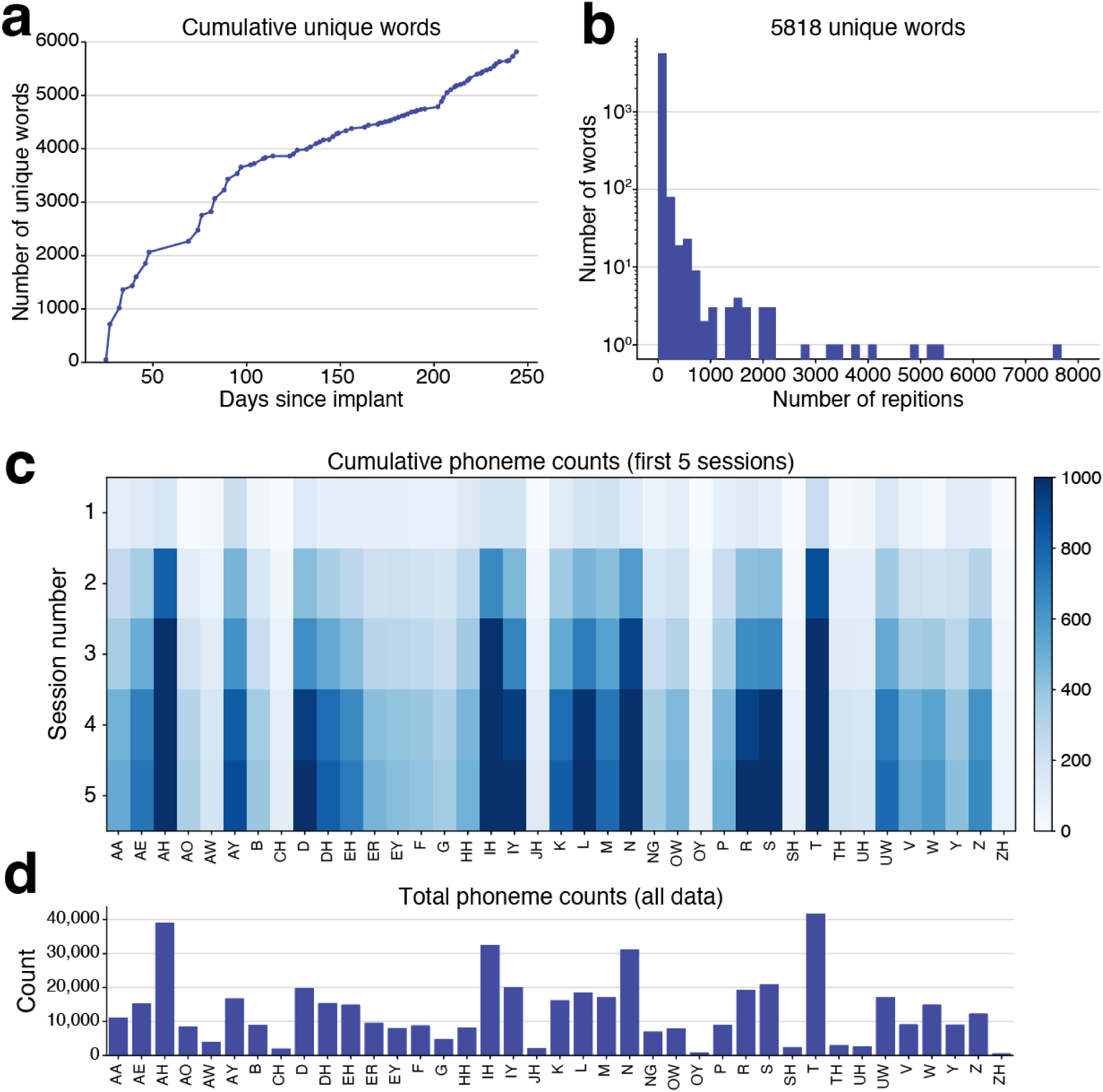
Phoneme and vocabulary coverage over time in the speech neuroprosthesis’ use. **a,** Number of cumulative unique spoken words over time, reaching 5818 unique words by session 80. Data from sessions 1-80 is included here, from both the Copy Task and from Conversation Mode. On session #2 (the first day of 125k-word vocabulary decoding), 706 unique words were spoken. Data from Conversation Mode is limited to sentences where the participant confirmed that the sentence was decoded correctly. **b,** Frequency of each word being spoken. The word “I” was the most frequent word, being spoken more than 7500 times. **c,** Cumulative phoneme counts from all data in the first five sessions. **d,** Total cumulative phoneme counts from sessions 1-80.

**Figure S5:**
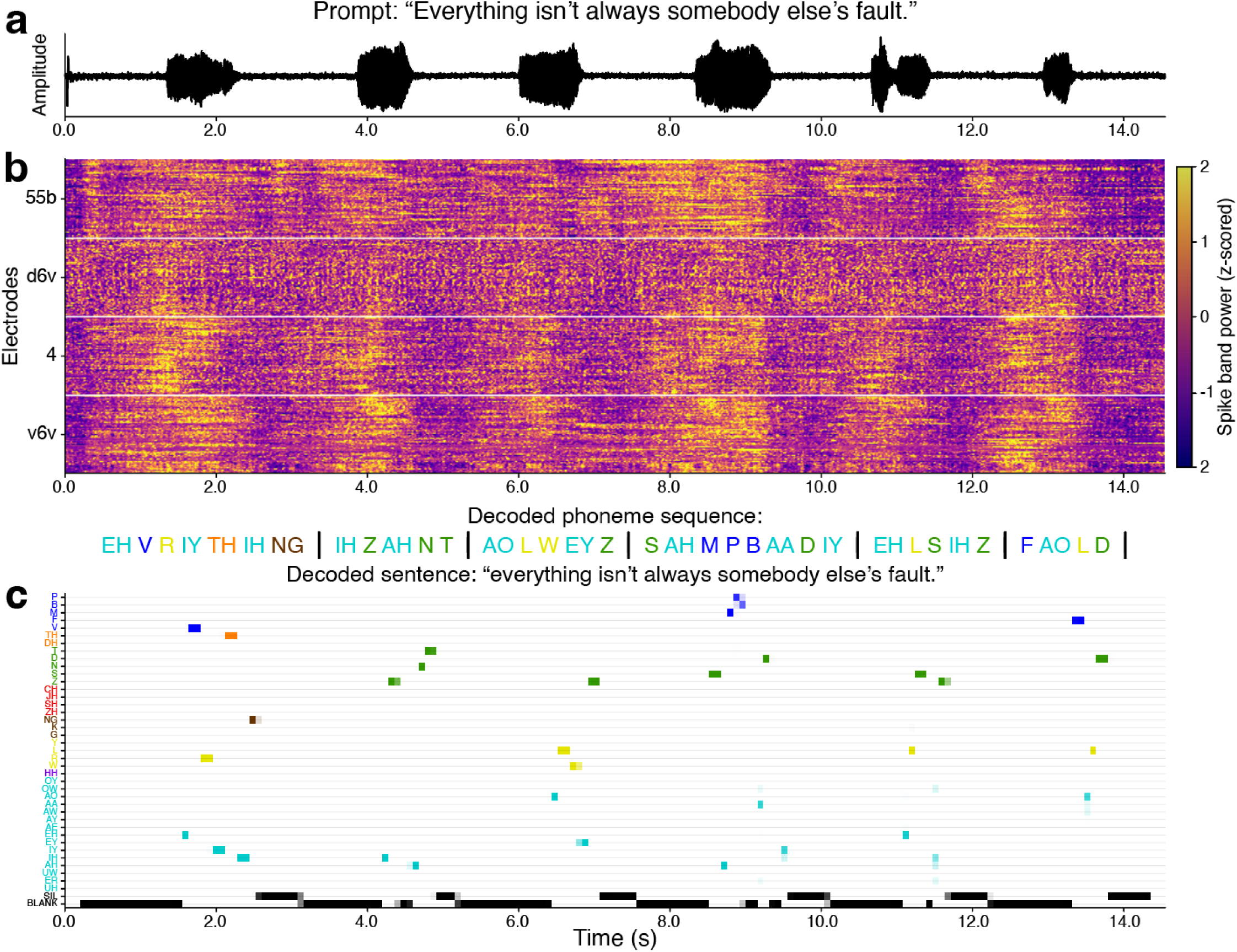
Audio, neural data, and predicted phonemes from an overt speech Copy Task trial. Data shown are from a trial of a Copy Task evaluation block during session 17. The participant was instructed to attempt to say the prompted sentence out loud at a normal volume and at his natural speaking rate. **a,** Microphone voltage trace of the participant’s attempted speech. Six individual words can be seen in the voltage trace, matching the prompted sentence. **b,** Neural data (z-scored spike band power) is plotted at 10 ms resolution. Electrodes are grouped by array, and arrays are labeled along the y axis and separated by horizontal white lines. **c,** Time course of phoneme predictions made by the speech neuroprosthesis in real-time during this trial. The highest probability phoneme sequence is written above the panel. Phonemes are grouped and colored by articulation type (see Fig. S16c).

**Figure S6:**
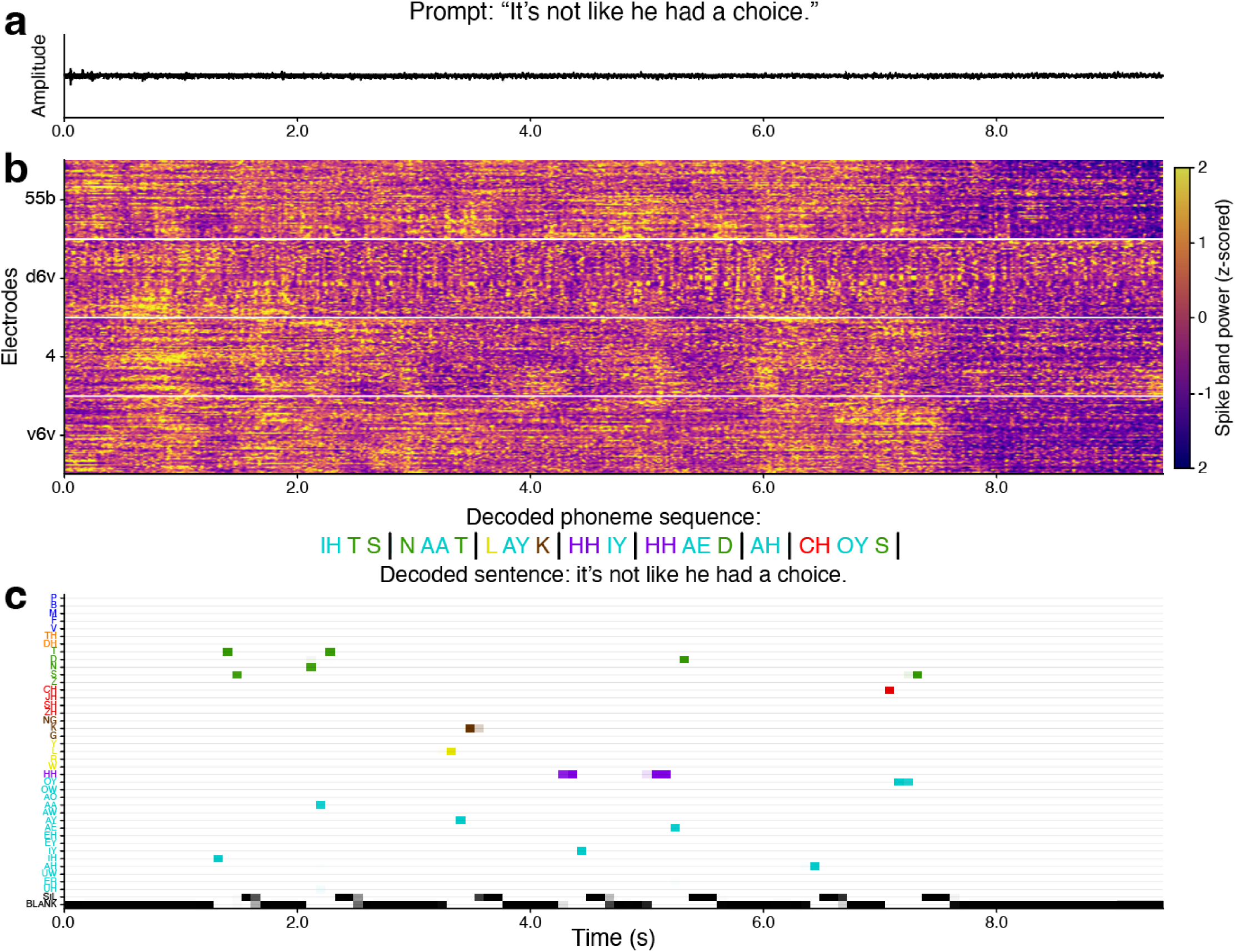
Audio, neural data, and predicted phonemes from a silent speech Copy Task trial. Data shown are from a trial of a Copy Task evaluation block during session 84. The participant was instructed to attempt to say the prompted sentence completely silently and at his naturalistic speaking rate. **a,** Microphone voltage trace of the participant’s attempted silent speech. No speech-related amplitude shifts can be seen in the microphone signal. **b,** Neural data (z-scored spike band power) is plotted at 10 ms resolution. Arrays are labeled along the y axis and separated by horizontal white lines. **c,** Time course of phoneme predictions made by the speech neuroprosthesis in real-time during this trial. The highest probability phoneme sequence is written above panel **c**. Phonemes are grouped and colored by articulation type (see Fig. S16c).

**Figure S7:**
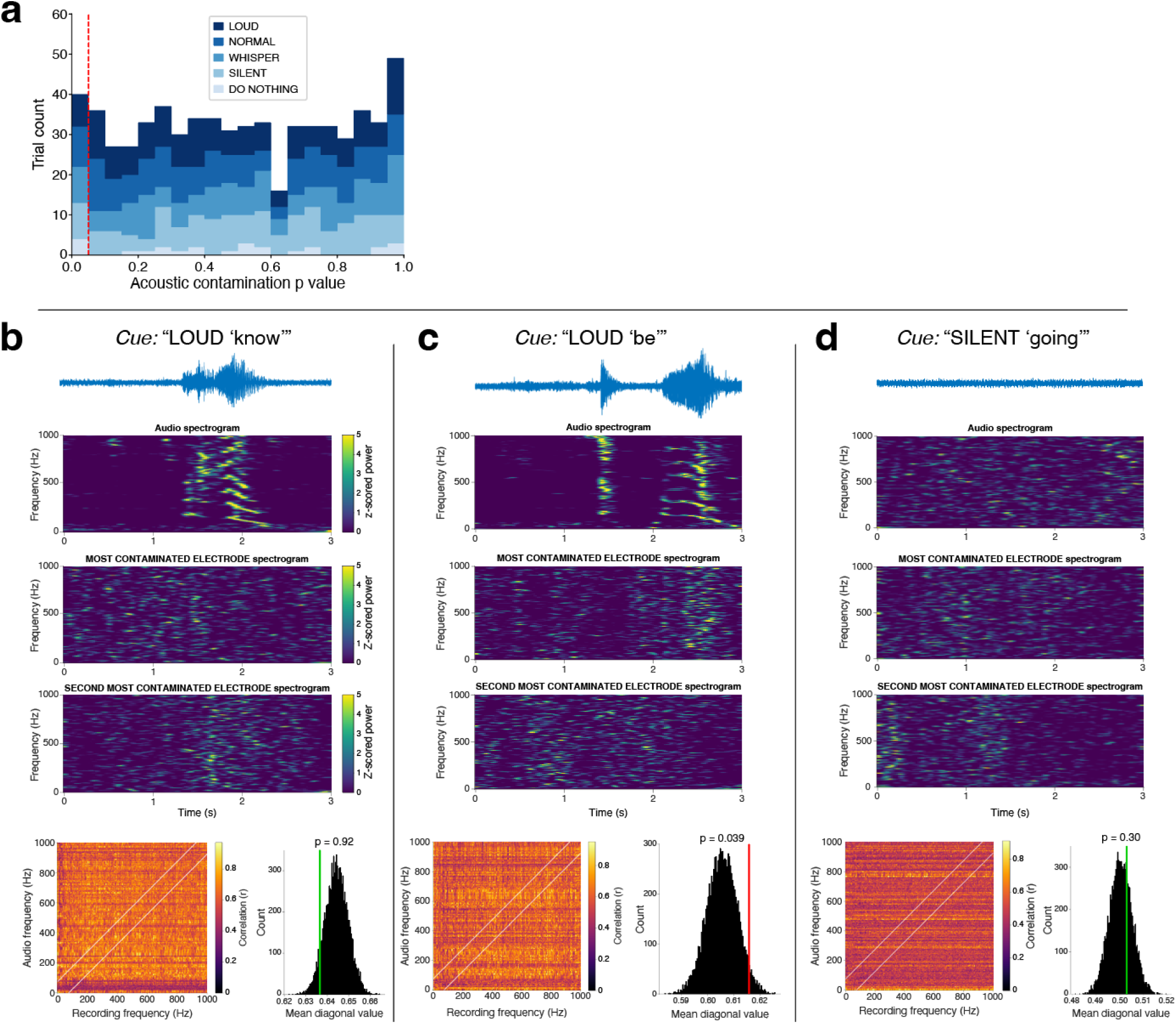
Neural data was not acoustically contaminated. **a,** Range of acoustic contamination p-values and the corresponding number of trials per attempted amplitude level with a particular p-value. The red line indicates a p-value of 0.05. Trials with p >= 0.05 (i.e., to the right of the red line) did not show evidence of acoustic contamination (93.9% of trials; 585/623). **b,** An example LOUD trial without evidence of acoustic contamination (p = 0.92). Spectrograms of even the most correlated electrodes (according to the analysis described in Section S4.03) do not have apparent speech-related power patterns (i.e., spectral patterns that line up with those of the audio spectrogram). **c,** An example LOUD trial with putative acoustic contamination (p = 0.039). The audio waveform shows two sounds, the first of which is the sound of a door closing, and the second of which is the sound of SP2’s attempted speech. Spectrograms of the most correlated electrodes have some potential speech-related power patterns matching the audio spectrogram. **d,** Example SILENT trial with no putative acoustic contamination (p = 0.3). There is no speech-related sound recorded on the microphone.

**Figure S8:**
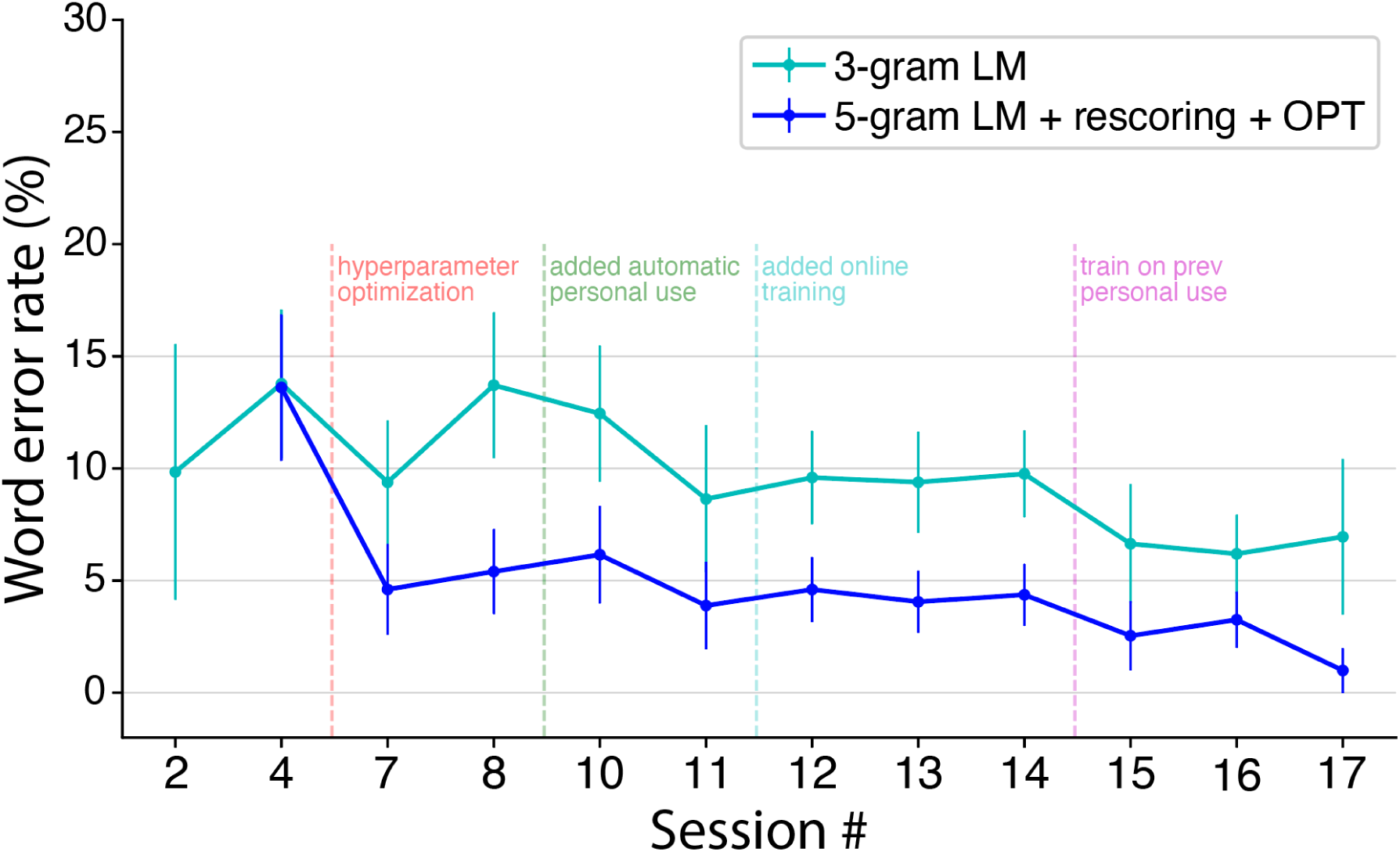
Decoding performance comparison between 3-gram and 5-gram language models. Comparison of offline evaluation performance using a 3-gram language model without rescoring (cyan line; as demonstrated online in ^6^) or a 5-gram language model with multi-stage rescoring of candidate sentences (blue line; as demonstrated offline in ^6^ and in closed loop in the main figures of this study). Both models used the same 125k-word English vocabulary. RNN-decoded phoneme probabilities from SP2’s closed-loop evaluation blocks were fed into both language models in offline analyses to compare their performance. Results were averaged over 5 RNN seeds. We used the 3-gram language model for online evaluation in session 2, and the upgraded 5-gram language model in subsequent sessions. After hyperparameter optimization of both the RNN decoder and the language model (red dashed line; between sessions 4 and 7), the 5-gram language model consistently outperformed the 3-gram model, resulting in lower word error rates.

**Figure S9:**
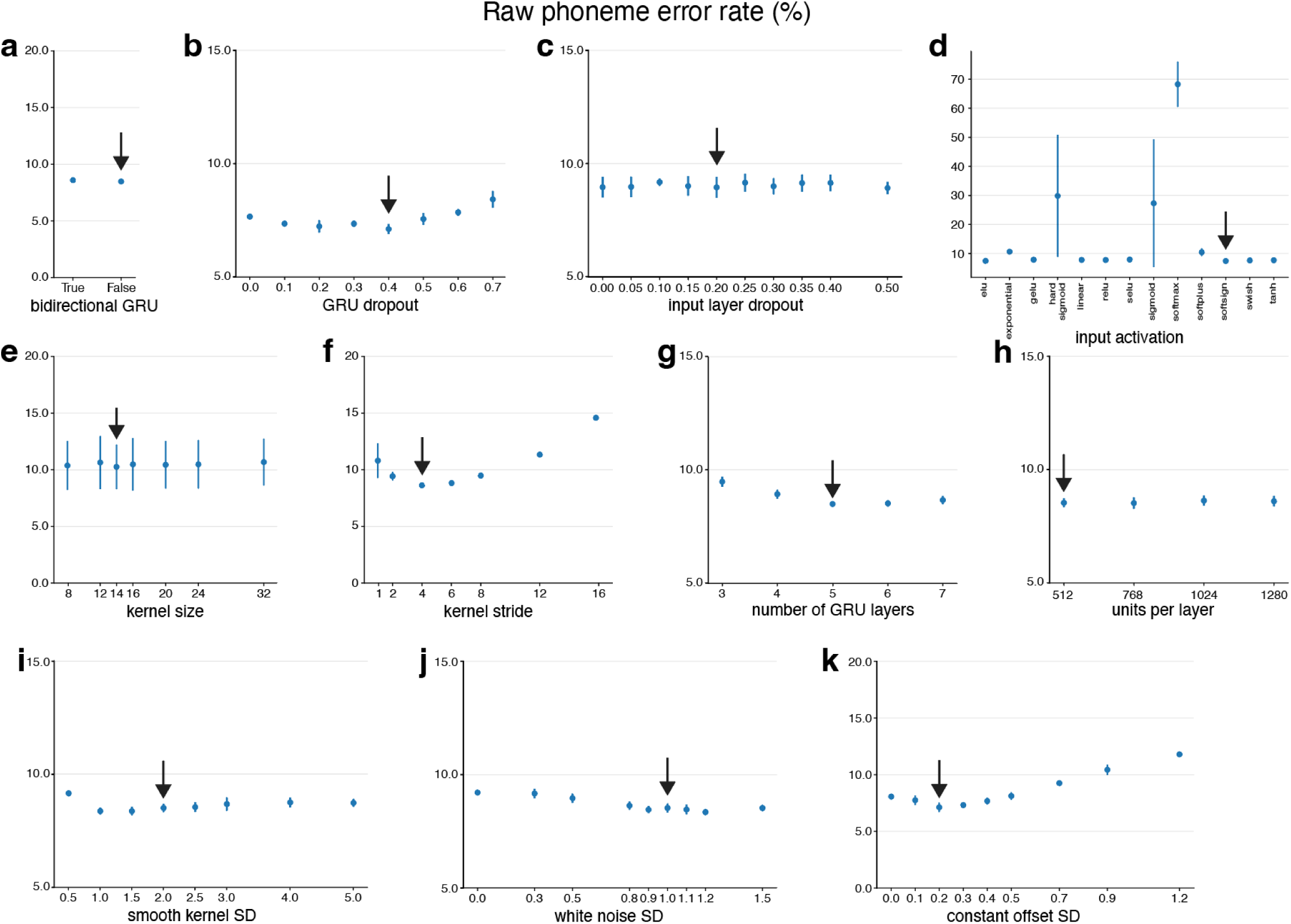
Offline parameter sweeps indicate near-optimal RNN parameter choices were used online. We tested the effect on raw (pre-language model) phoneme error rate as a function of several RNN parameters. Each point in each plot represents the average phoneme error rate (± standard deviation) of 10 RNN seeds trained with the corresponding parameter, on data from the first *n* sessions. This process was repeated twice throughout data collection to ensure that we were using optimal RNN decoding parameters in subsequent online decoding sessions. Here, results from the first 12 sessions of data are shown. Black arrows represent parameters used in closed-loop evaluation. Tested parameters include: **a**, Bidirectional vs. unidirectional GRU layers. **b,** Dropout percentage for GRU layers. **c,** Dropout percentage for input layers. **d,** Activation type for input layers. **e,** “Kernel size” (i.e., the number of 20 ms bins stacked together as input and fed into the RNN at each time step). **f,** “Kernel stride” (a stride of N means the RNN steps forward only every N time bins). **g,** Number of GRU layers. **h,** Number of units per GRU layer. **i,** Standard deviation of the Gaussian smoothing kernel (larger number means more smoothing). This parameter was not quite optimized for closed-loop decoding. **j**, Standard deviation of white noise dynamically added to training data during RNN training for data augmentation. This parameter was not quite optimized for online decoding. **k,** Standard deviation of constant offset noise added to training data during RNN training for data augmentation.

**Figure S10:**
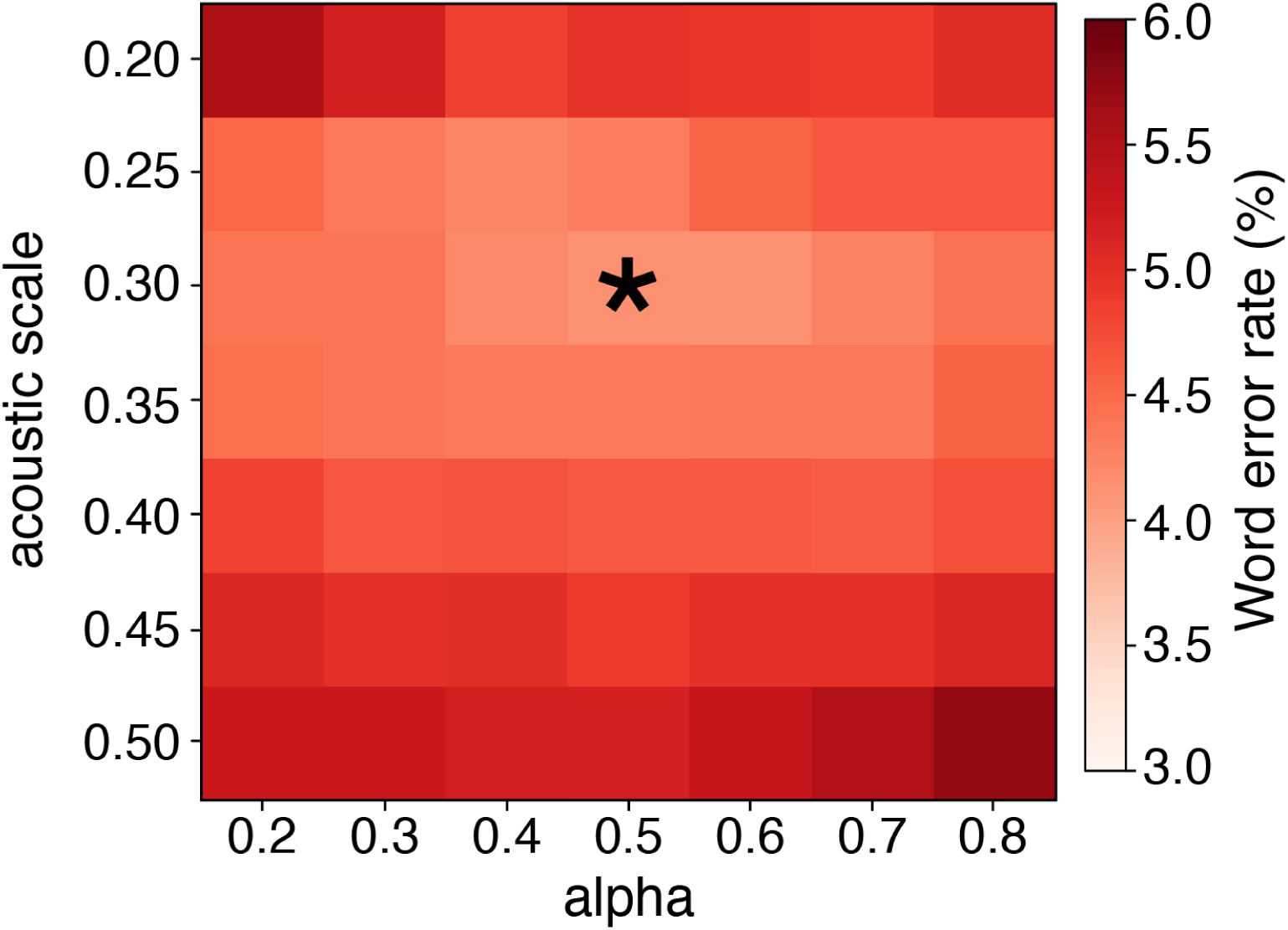
Offline language model parameter sweeps informed subsequent online parameter choices. We conducted offline analyses to identify the optimal language model parameters (i.e., the parameters yielding the lowest word error rate). An RNN was trained on all data from the first *n* sessions (in this case, *n*=12), and the RNN-decoded phoneme probabilities from held-out validation trials were fed into 5-gram language models initialized using a range of parameters. Varied parameters included the blank penalty, acoustic scale, and alpha values (Section S3; also see supplemental methods section of ^6^). Although varying the blank penalty (set to log(9) here) did not result in a large change in word error rate, the acoustic scale and alpha parameters made appreciable differences in accuracy. This language model parameter sweep was repeated thrice throughout data collection, and consistently showed that an acoustic scale of 0.3 and an alpha of 0.5 (denoted with * in the plot) resulted in the lowest word error rate. These optimal language model parameters were subsequently used for online speech decoding.

**Figure S11:**
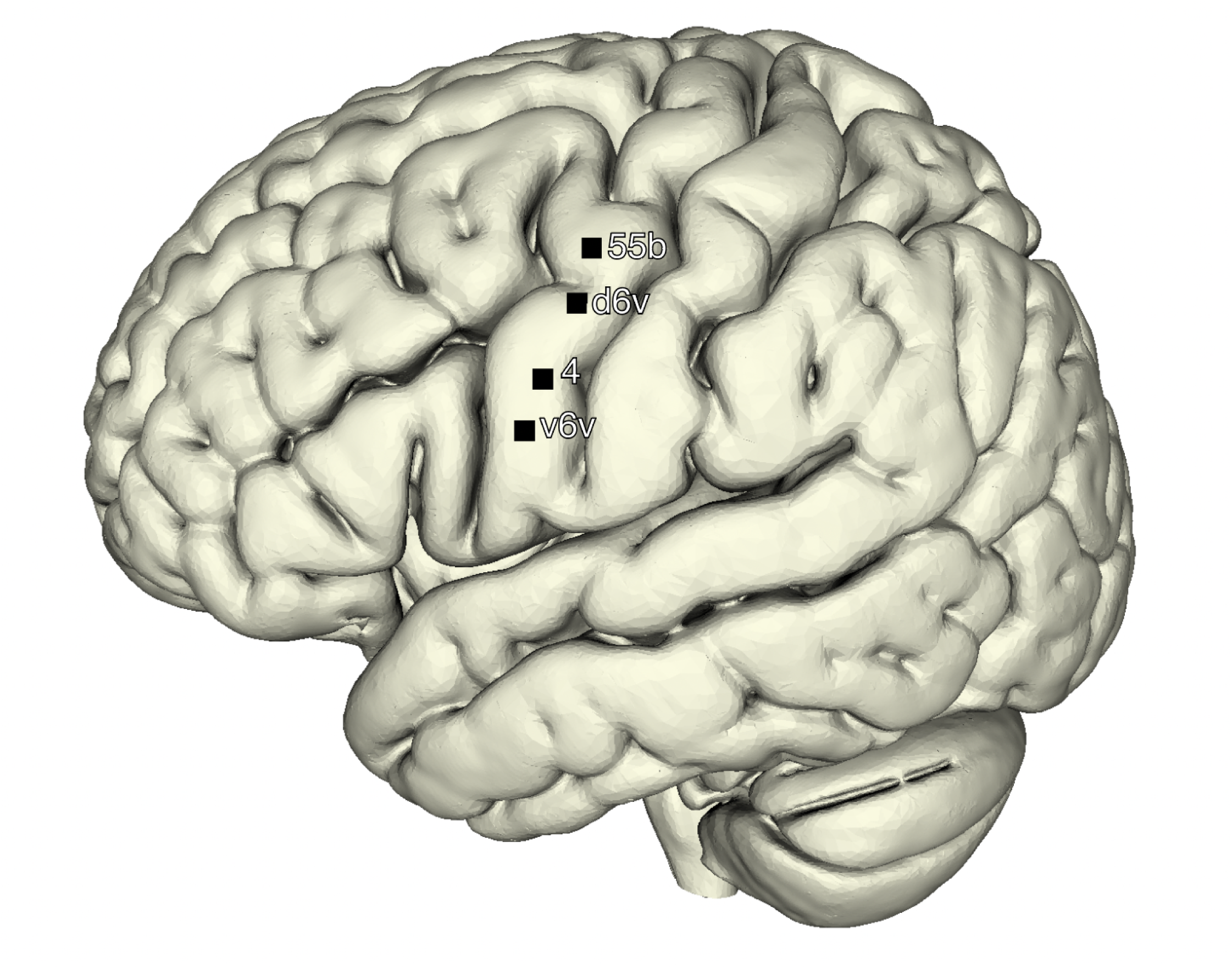
Estimated array locations on a standard brain. Here the locations of the four implanted microelectrode arrays are shown on the MNI2009b asymmetric brain. Post-operative estimates of the Utah array insertion locations were performed by corroborating intra-operative photographs with postoperative CT scan co-registered to the pre-operative anatomical T1 MRI. After identifying the array location points in ACPC space, a non-linear deformation was performed from SP2’s anatomical imaging to the MNI2009b template. Details of the registration procedure are provided in Section S1.12.

**Figure S12:**
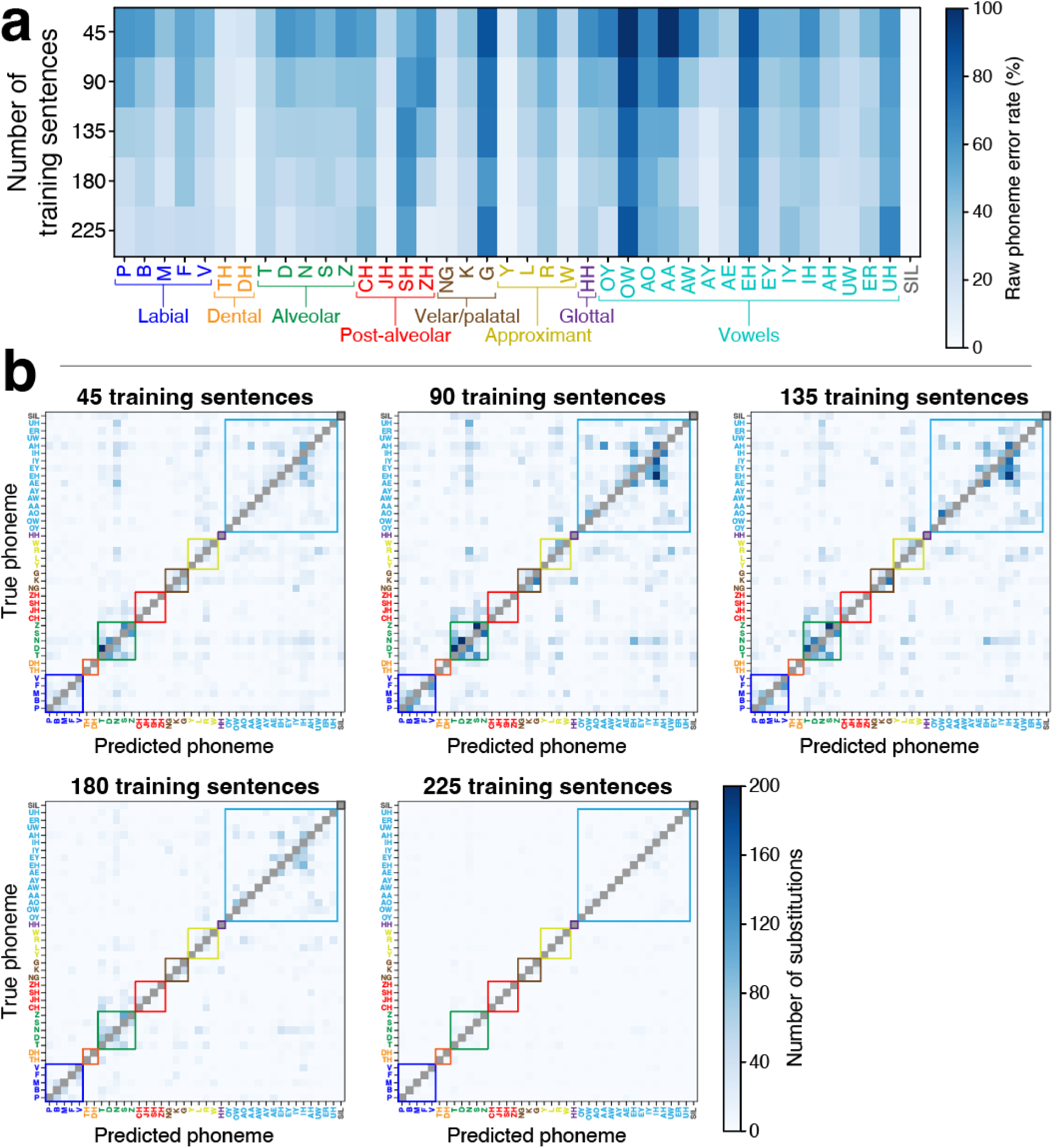
The decoder could be rapidly calibrated to predict phonemes accurately. To characterize the dynamics of training a speech neuroprosthesis with limited data, we trained decoders (offline) on all possible combinations of the first five blocks from session 2 (31 total combinations). Each block had 50 sentences, 45 of which were used for training, and the remaining 5 were used to validate decoding performance during training. For each combination of training data, five RNNs were trained with random seeds, for a total of 155 RNNs (31 combinations, 5 seeds each). All RNNs were evaluated on data from two held-out blocks, also from session 2. **a,** Phoneme error rate as a function of the number of sentences used to train a decoder. **b,** Phoneme substitution matrices for each amount of training data. See the caption of Fig. S17 for more details. The color bar (bottom right) applies to all plots in **b**.

**Figure S13:**
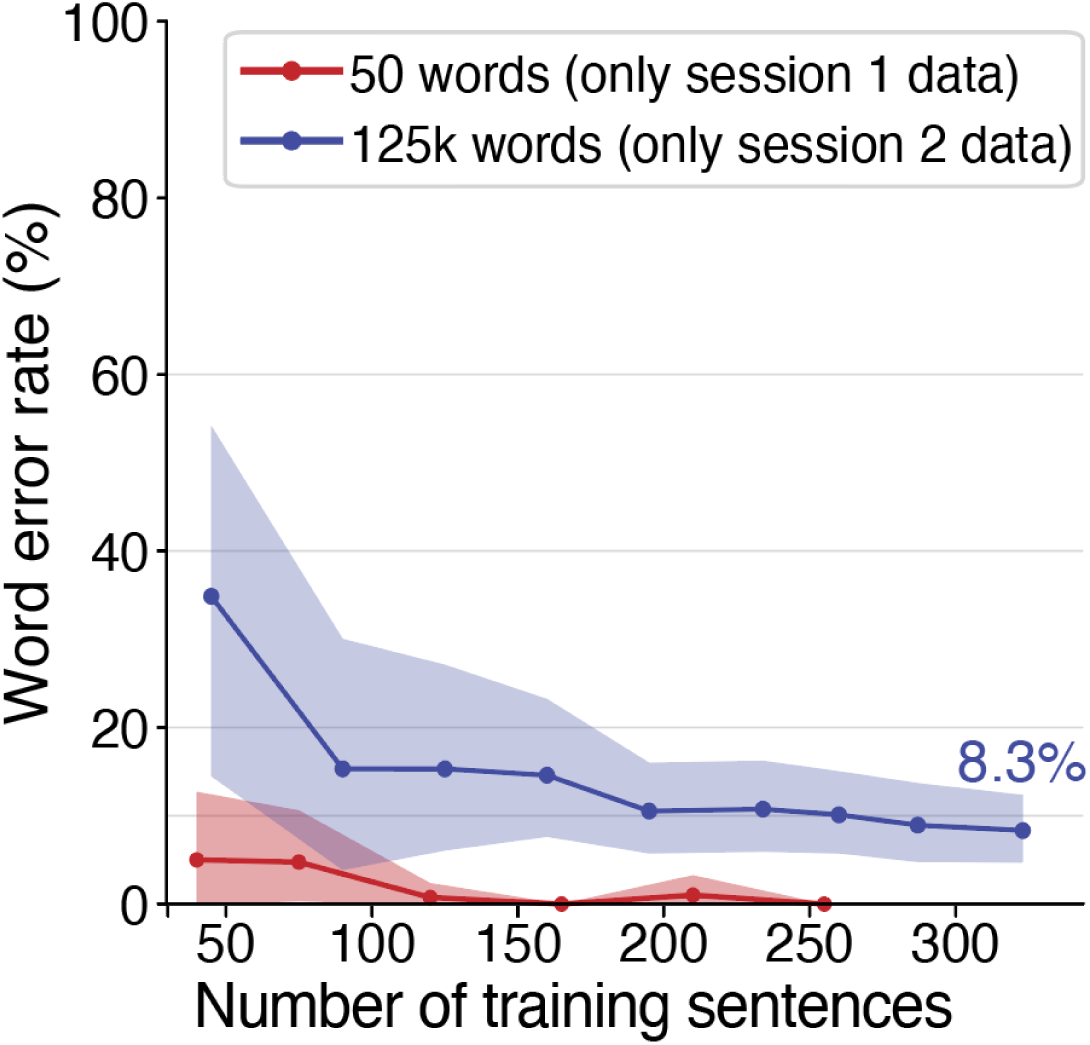
Offline decoding analyses indicate more rapid calibration was possible. Offline recreation of “day 1” performance for 50-word (red) and 125,000-word (blue) vocabularies with optimal decoding hyperparameters. Word error rate is plotted as a function of the number of training sentences. Decoders for 50-word and 125,000-word vocabulary sizes were trained on subsets of data from only sessions 1 or 2, respectively.

**Figure S14:**
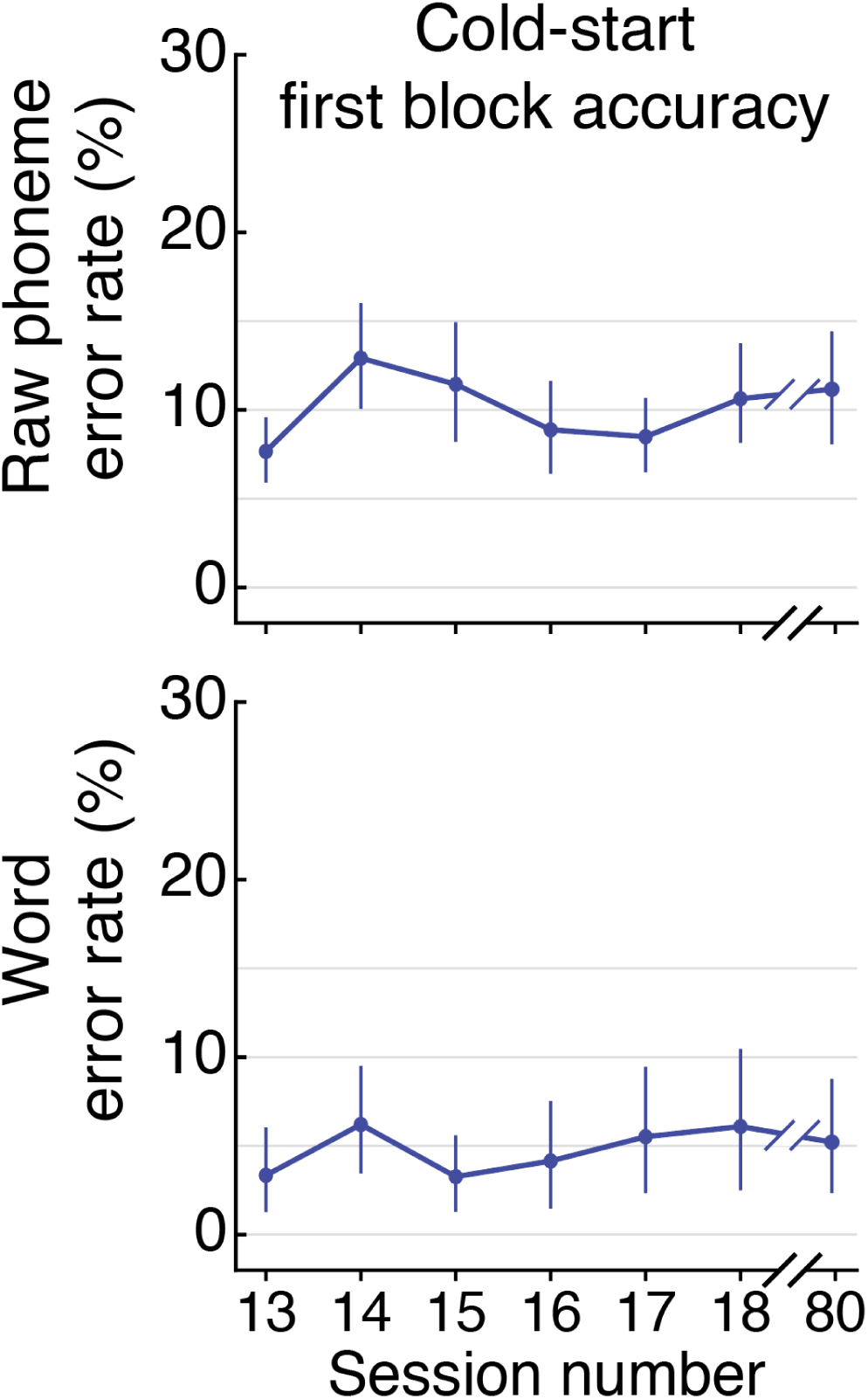
Speech decoding accuracy in the first block of each session. Raw phoneme error rates (top; average 10.3%, 95% CI: 9.2% to 11.3%) and word error rates (bottom; average 4.8%, 95% CI: 3.7% to 6.2%) for the first 50 sentences of evaluation sessions 13, 14, 15, 16, 17, 18, and 80. These 50 sentences were collected at the start of each session, right after the diagnostic block (Section S1.07). Data from the first block of session 69 is not included here due to a technical issue during the start of that session. For just the first twenty sentences of each block, the word error rate was 5.8% (95% CI: 3.6% to 8.3%). For just the first five sentences of each block, the word error rate was 12.0% (95% CI: 6.7% to 18.6%); this lower performance is because these sentences did not benefit from the full adaptive neural feature normalization and online decoder fine-tuning. In each of these first-of-the-session blocks, online decoder fine-tuning began after the first 10 sentences in sessions 13-18, and after the first 5 sentences in sessions 18 and 80.

**Figure S15:**
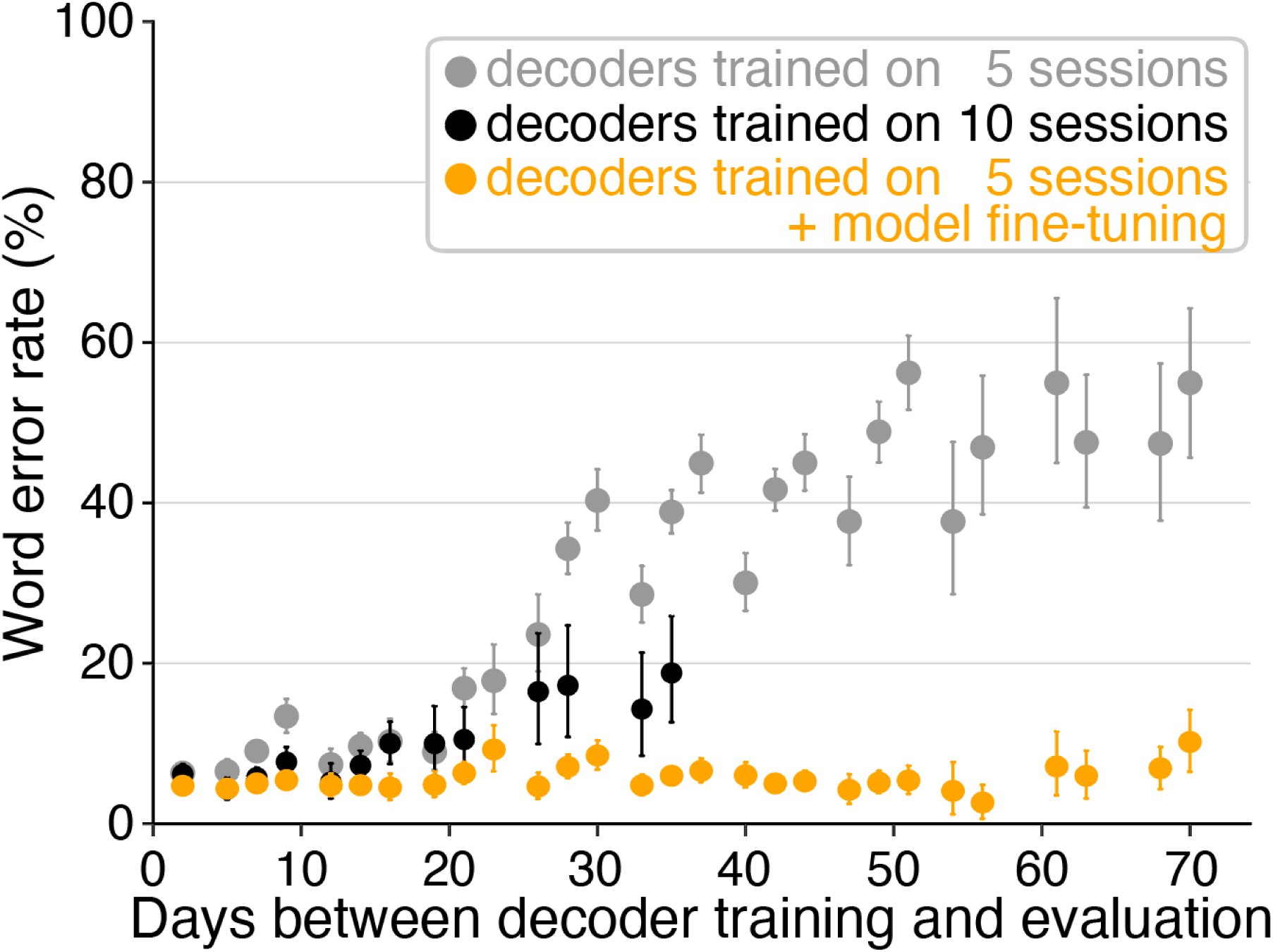
Offline decoding analyses indicate decoding can remain stable. Decoding stability over time with no model fine-tuning (gray and black points) or with model fine-tuning (orange points). Decoders were trained on data from 5 (gray, orange) or 10 (black) sequential sessions, and then evaluated on data from increasingly distant future sessions. Word error rate is plotted as a function of the number of days between the final day of data used to train each decoder and the date of the evaluation data. For the orange points, the decoder model was fine-tuned on previously seen data after inference on each consecutive sentence (Section S2.03). This analysis is limited to data from sessions 1-26. These results suggest that without online fine-tuning, a user could reasonably expect good performance for at least two weeks, but that eventually new training data will need to be collected to maintain performance. However, the online fine-tuning analysis suggests that with this capability, high decoding accuracy can continue for much longer.

**Figure S16:**
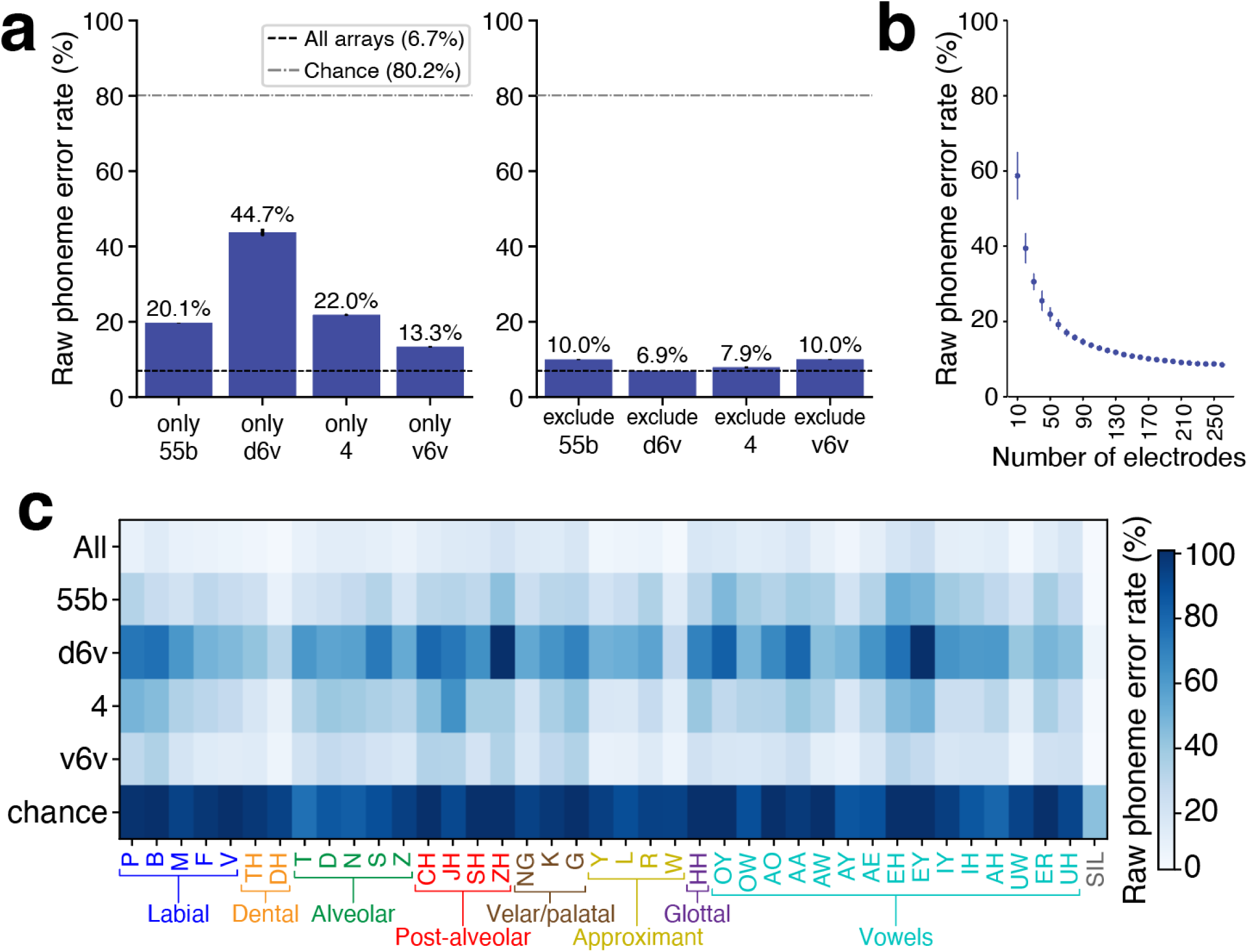
Phoneme decoding accuracy for each microelectrode array. For all data presented in this figure, RNNs were trained on all data from sessions 1-18 and evaluated on randomly-chosen held-out validation trials (10% of total trial count). **a,** Analysis of phoneme error rates derived from the RNN output. Left, decoding contribution of each individual array (mean ± standard deviation from 5 RNN seeds). Right, performance if any single array was removed. The black dashed line represents decoding performance using all four arrays. Omitting the dorsal 6v array did not have a detrimental effect on the phoneme error rate. The gray dashed line represents chance decoding performance (Section S4.01). **b,** An evaluation of decoding accuracy (phoneme error rate, mean ± standard deviation) when varying the number of electrodes used (randomly selected from all arrays; Section S4.01). **c,** Individual phoneme decoding accuracy for each array, compared to using all four arrays and chance.

**Figure S17:**
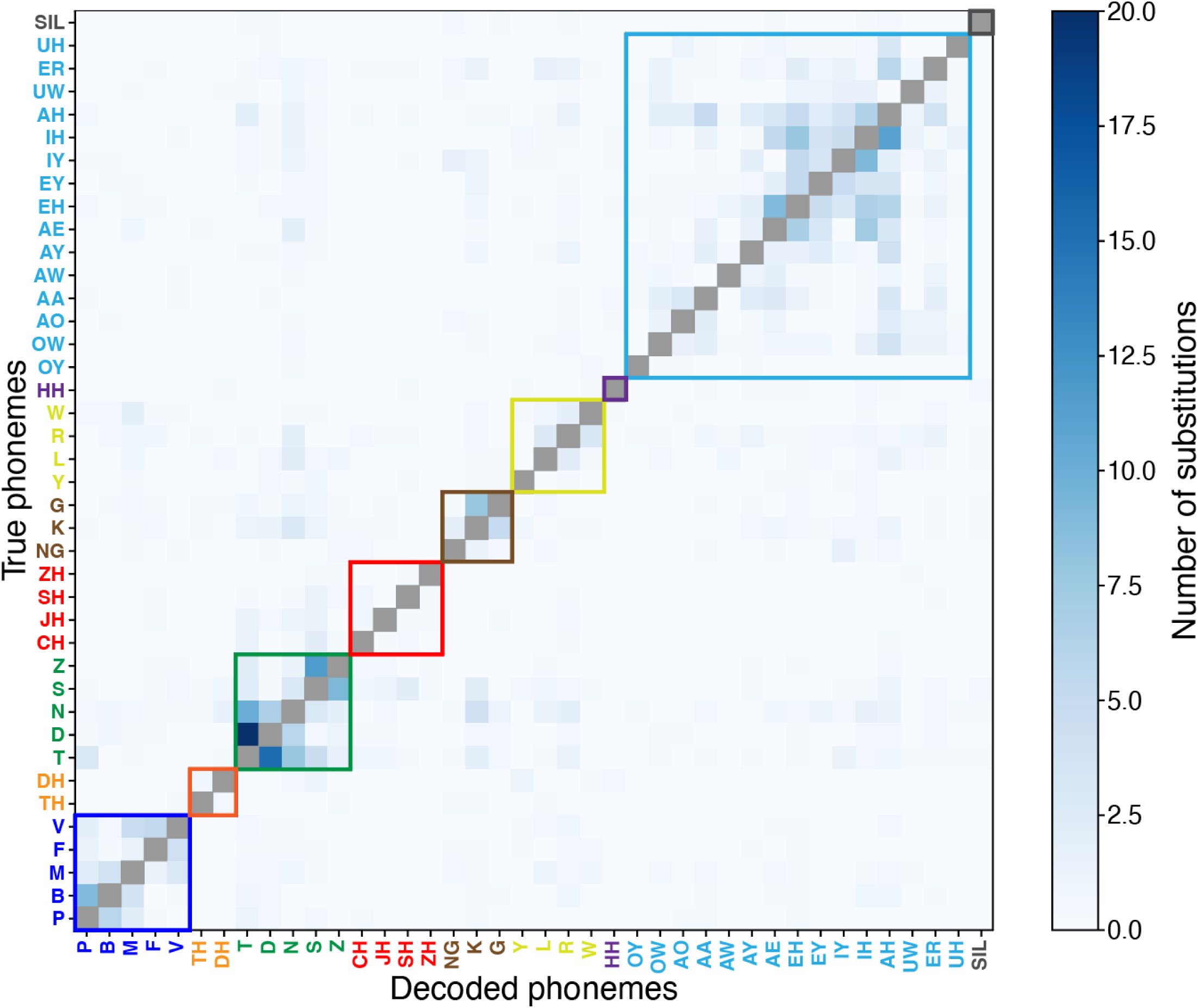
Phoneme substitution errors observed across all real-time decoded sentences. Substitution error rate between true and decoded phoneme sequences using all data from sessions 1-83. Entry *(i,j)* in the matrix indicates the number of substitutions between true phoneme *i* and decoded phoneme *j*. Substitutions were identified using an edit distance algorithm that determines the minimum number of insertions, deletions, and substitutions required to make the raw (pre-language model) decoded phoneme sequence match the true phoneme sequence. As phonemes could not be substituted for themselves, entries along the diagonal are colored gray to indicate that they are excluded from this analysis. The majority of substitutions appear to occur between phonemes that are articulated similarly (within place of articulation groupings indicated by the boxes colored the same as in Fig. 2e), including between voiced and unvoiced consonant pairs (e.g., /p/ vs /b/, and /t/ vs /d/).

**Figure S18:**
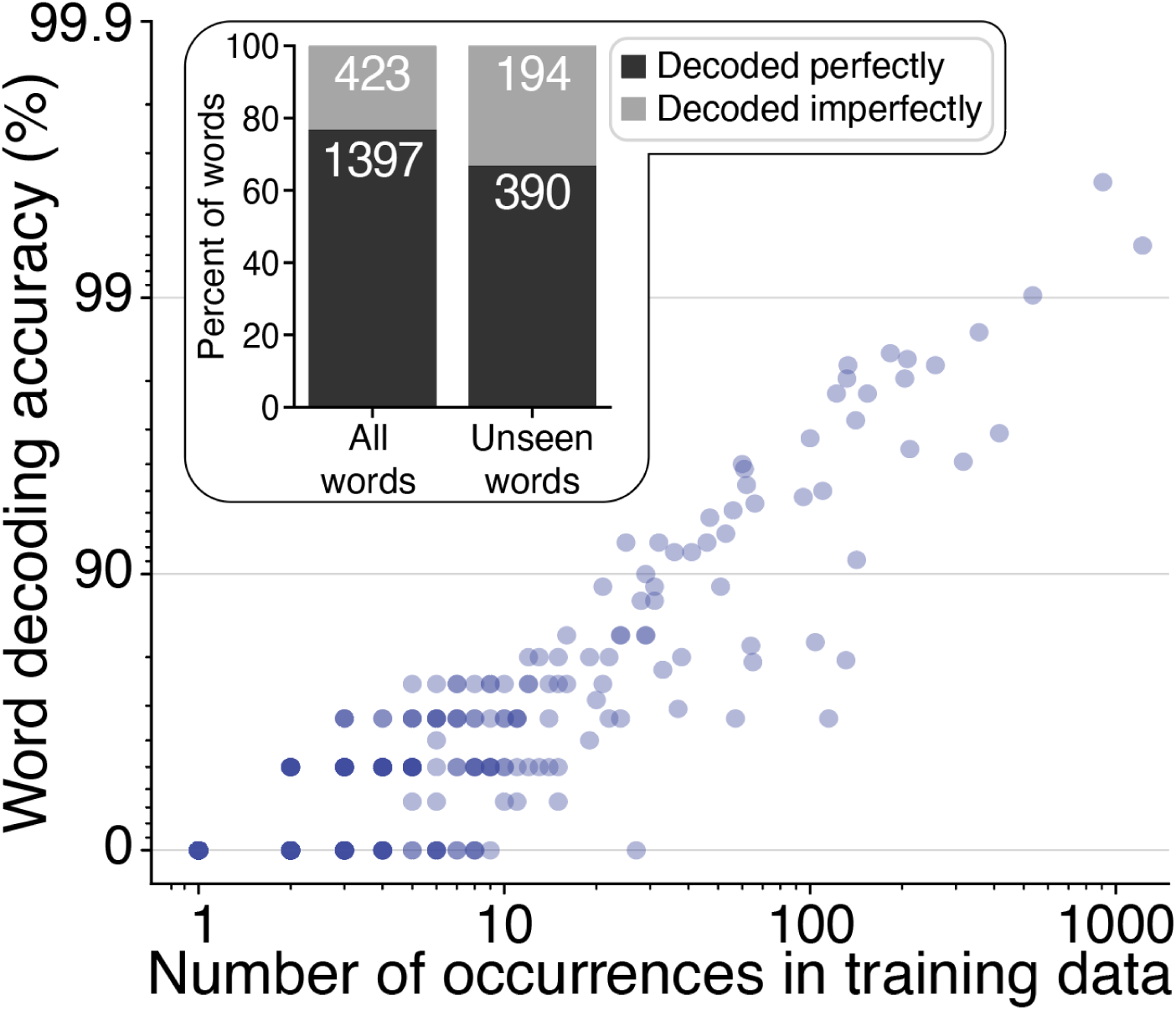
Offline decoding analyses indicate high generalizability to new words. This offline analysis characterizes how many training examples the system needs to learn to decode words that were initially decoded as incorrect. Word decoding accuracy (log scale) as a function of the number of occurrences of the word in the decoder’s training data (log scale), for all words that were initially decoded incorrectly. After about ten occurrences of a “difficult” word, it could be correctly decoded most of the time. Inset: decoding accuracy for all words (left, 76.8%) and for words that had never been seen in the prior training data (right, 66.8%). Here, decoding accuracy is presented in a binary fashion where words were determined to either have been “decoded perfectly” (i.e., correct over all occurrences of that word in the prompted sentence) or imperfectly. For example, a word that was attempted 1000 times but only decoded correctly 999 times would be classified as “decoded imperfectly” here. This analysis is limited to Copy Task and Conversation Mode data from sessions 1-32. These results indicate that some of the sentence-to-sentence variability in word error rates can be attributed to whether there were “new” (to the decoder) words encountered in that sentence.

**Figure S19:**
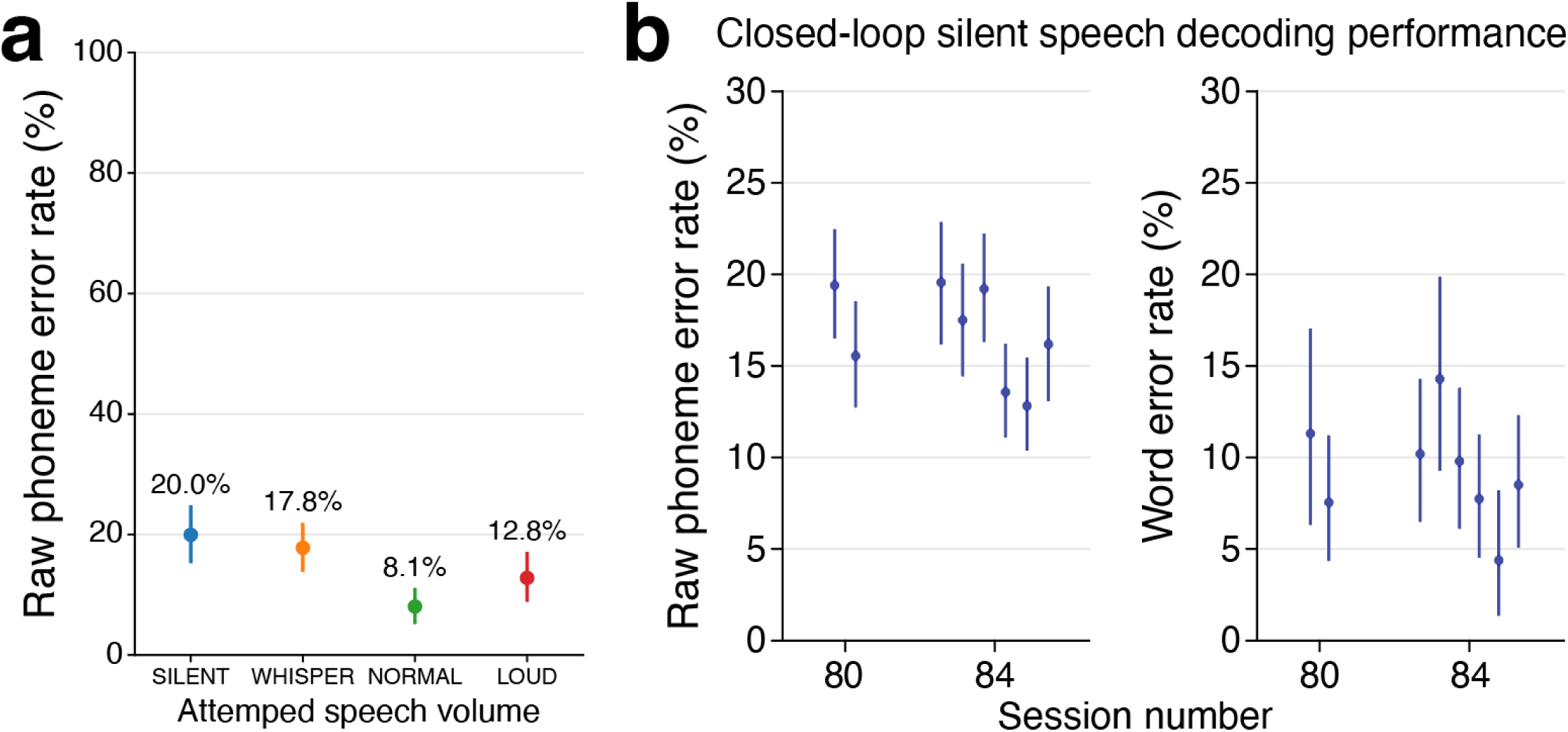
Decoding accuracy as a function of attempted speech volume. **a,** Phoneme error rate across all 623 trials of the Speech Amplitude task where the participant spoke one of six different words at each attempted speech amplitude using a decoder that was trained on the “NORMAL” speech amplitude condition (see Section S4.03 for task details). Points represent the average phoneme error rate and bars represent the 95% confidence interval. **b,** Closed-loop silent speech decoding from sessions 80 and 84 when performing the 125k word vocabulary Copy Task. For each session, the speech neuroprosthesis was trained on overt speech from all prior sessions, and then tested and fine-tuned on silent speech Copy Task data during the course of the session. Phoneme error rates (left) and word error rates (right) are shown for each block of silent speech decoding. Circles represent means and vertical lines represent 95% confidence intervals. Note that both training and evaluation blocks are included here. During these silent speech blocks, SP2 attempted to speak at 52.3 words per minute (95% confidence interval: 51.3 to 53.3).

**Figure S20:**
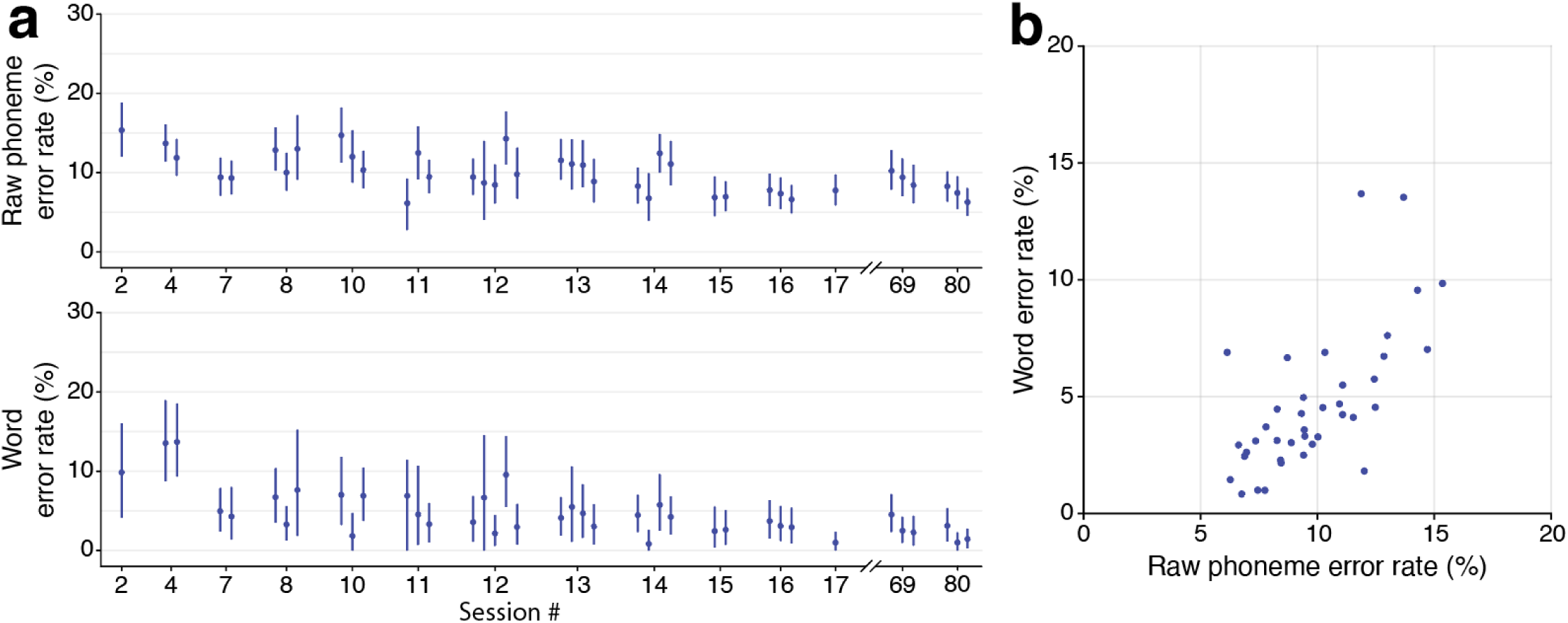
Online decoding accuracy for individual evaluation blocks. **a,** Raw phoneme error rates (top) and word error rates (bottom) are shown (mean and 95% confidence interval) for each individual evaluation block in each session. Additional details are provided in Table S3. **b,** This scatter plot shows the relationship between average word error rate and average raw phoneme error rate across all evaluation blocks (each block is one point in the plot).

**Figure S21:**
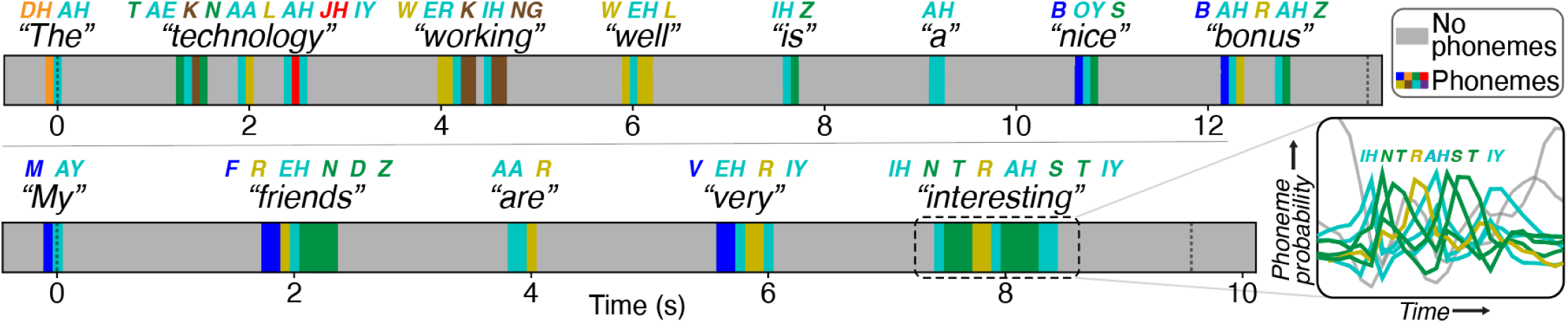
Speech detection and predicted phonemes during Conversation Mode. Timeline of two example sentences showing the most probable phoneme at each time step, as indicated by RNN outputs. Gray intervals indicate when the highest output probability is silence, while colored segments show the most probable phoneme. Phonemes are colored according to the phoneme category that they belong to (see Fig. S16c). Vertical dashed lines delineate the onset and termination of sentence construction. The decoded phonemes and words are annotated above each visualization. *Inset*, detailed view of selected phoneme probabilities as SP2 attempts to say the word “interesting”.

**Figure S22:**
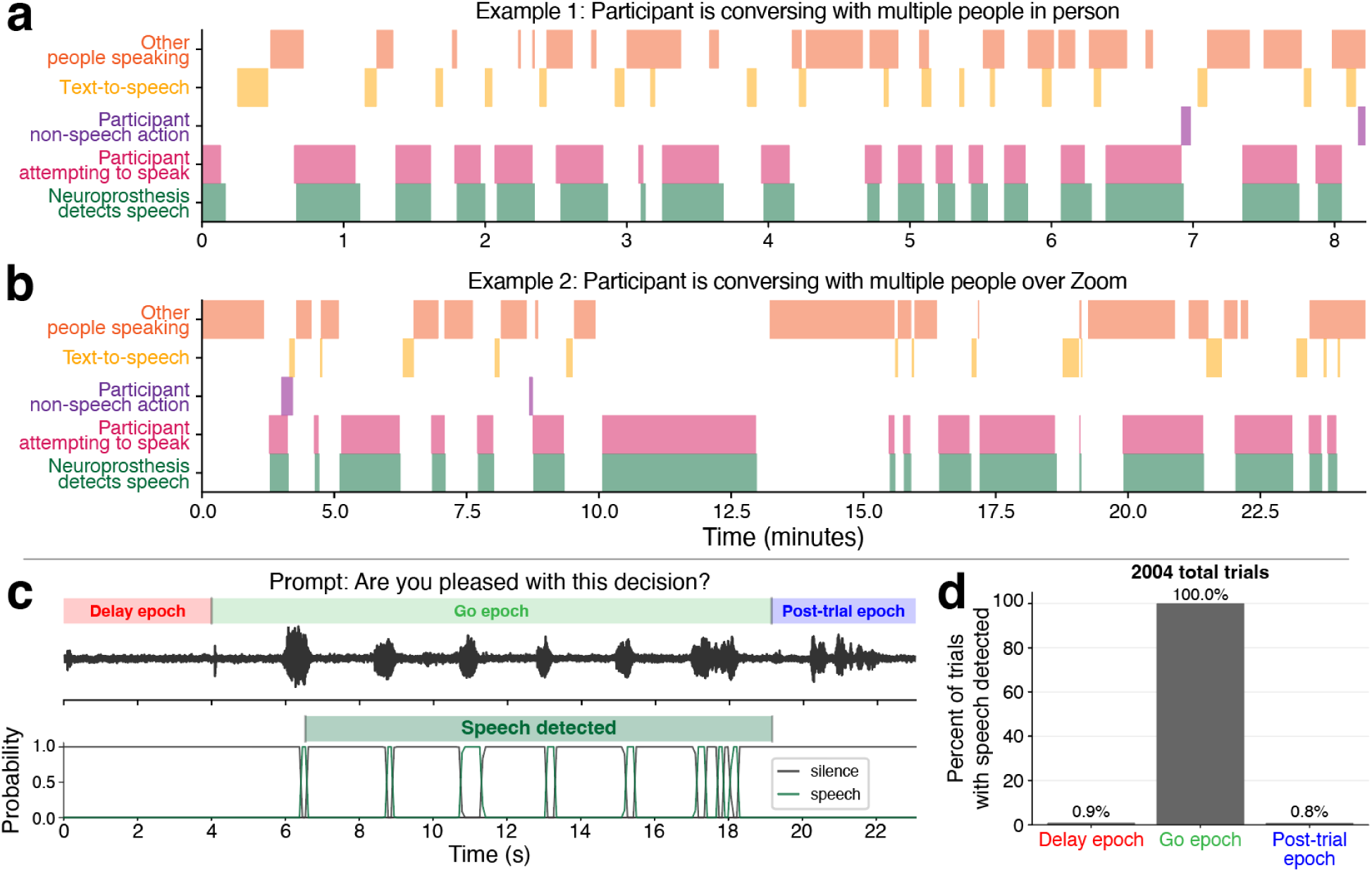
Reliable and specific speech detection underlies Conversation Mode. **a,** Manually labeled epochs from a Conversation Mode block in session 61 where the participant used the speech neuroprosthesis to have a conversation with researchers and family members who were in the room with him. Labeled epochs include (1) when other people were speaking, (2) when the participant was attempting to speak, (3) when the neuroprosthesis detected that the participant was attempting to speak, (4) when the participant was engaging in a non-speech action (e.g., yawning, coughing, laughing, etc.), and (5) when text-to-speech decoded sentences were played aloud. Timestamps for each event were manually labeled with reference to video recordings, audio recordings, and decoder data files. The neuroprosthesis reliably detected when the participant attempted to speak, and did not erroneously detect speech when he was not attempting to speak. **b,** Additional example from the same session where the participant used the neuroprosthesis to speak to people over a Zoom call. **c,** Example trial where we applied (offline) the same speech detection algorithm used in Conversation Mode to continuous neural data collected during the Copy Task. Here we could divide each trial into three epochs: (1) when the participant was not speaking and there was usually not background noise (delay epoch, red); (2) when the participant was speaking and there was usually not background noise (go epoch, green); and (3) when the participant was not speaking and he was listening to the text-to-speech audio for the previous decoded sentence (post-trial epoch, blue). The black trace (top) shows the microphone amplitude for this trial, where speech-related modulations can be seen during the go epoch, and text-to-speech audio occurs in the post-trial epoch. The bottom line plot shows the probability with which the decoder is predicting silence (gray) or speech (green) at each 80 ms time step. Speech is only detected during the go epoch. **d,** Frequency with which speech was detected in epochs from 2,004 Copy Task trials from closed-loop blocks during 10 speech evaluation sessions, using the same methodology as described in **c**. Speech was detected during 100% of go epochs, and in <1% of delay and post-trial epochs.

## Supplemental Tables

**Table S1:**
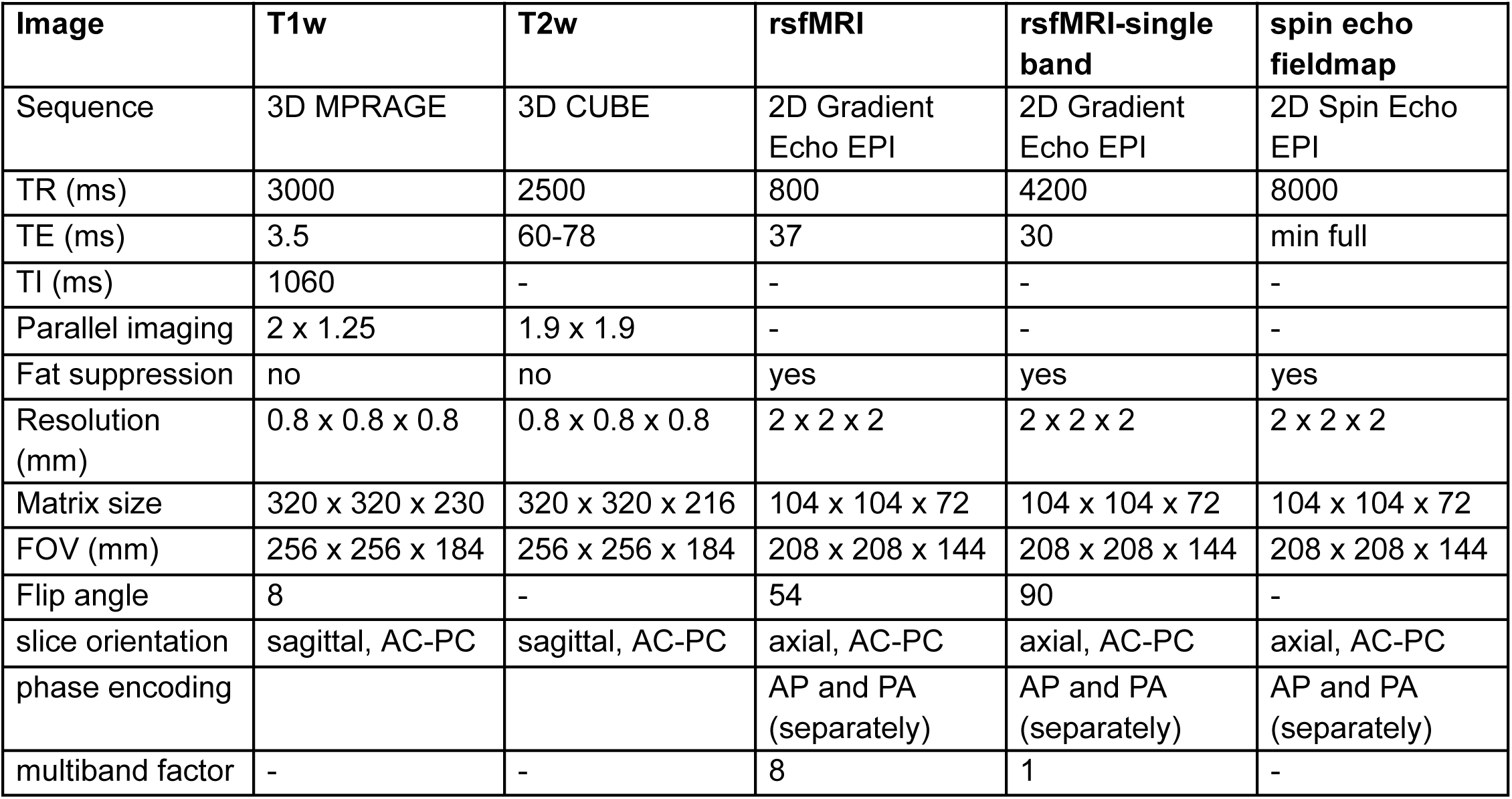
MRI Scan Parameters.

| Image | T1w | T2w | rsfMRI | rsfMRI-single band | spin echo fieldmap |
| --- | --- | --- | --- | --- | --- |
| Sequence | 3D MPRAGE | 3D CUBE | 2D Gradient Echo EPI | 2D Gradient Echo EPI | 2D Spin Echo EPI |
| TR (ms) | 3000 | 2500 | 800 | 4200 | 8000 |
| TE (ms) | 3.5 | 60-78 | 37 | 30 | min full |
| TI (ms) | 1060 | - | - | - | - |
| Parallel imaging | 2 x 1.25 | 1.9 x 1.9 | - | - | - |
| Fat suppression | no | no | yes | yes | yes |
| Resolution (mm) | 0.8 x 0.8 x 0.8 | 0.8 x 0.8 x 0.8 | 2 x 2 x 2 | 2 x 2 x 2 | 2 x 2 x 2 |
| Matrix size | 320 x 320 x 230 | 320 x 320 x 216 | 104 x 104 x 72 | 104 x 104 x 72 | 104 x 104 x 72 |
| FOV (mm) | 256 x 256 x 184 | 256 x 256 x 184 | 208 x 208 x 144 | 208 x 208 x 144 | 208 x 208 x 144 |
| Flip angle | 8 | - | 54 | 90 | - |
| slice orientation | sagittal, AC-PC | sagittal, AC-PC | axial, AC-PC | axial, AC-PC | axial, AC-PC |
| phase encoding |  |  | AP and PA (separately) | AP and PA (separately) | AP and PA (separately) |
| multiband factor | - | - | 8 | 1 | - |

**Table S2:** Data collection sessions.

| Session Number | Post-implant day | Description | Speech Data |
| --- | --- | --- | --- |
| 1 | 25 | 50-word training data collection and decoding | 290 Copy Task 50-word-vocab training sentences.<br>50 Copy Task 50-word-vocab evaluation sentences. |
| 2 | 27 | 125,000-word training data collection and evaluation | 330 Copy Task Switchboard training sentences.<br>40 Copy Task 50-word-vocab training sentences.<br>50 Copy Task 50-word-vocab evaluation sentences.<br>30 Copy Task Switchboard evaluation sentences.<br>10 Conversation Mode sentences. |
| 3 | 32 | 125,000-word training data collection | 300 Copy Task Switchboard training sentences. |
| 4 | 34 | 125,000-word training data collection, evaluation, and personal use | 280 Copy Task Switchboard training sentences.<br>100 Copy Task Switchboard evaluation sentences.<br>74 Conversation Mode sentences. |
| 5 | 39 | 125,000-word training data collection and other experiments | 140 Copy Task Switchboard training sentences. |
| 6 | 41 | 125,000-word training data collection | 200 Copy Task Switchboard training sentences. |
| 7 | 46 | 125,000-word training data collection and evaluation | 300 Copy Task Switchboard training sentences.<br>100 Copy Task Switchboard evaluation sentences. |
| 8 | 48 | 125,000-word training data collection and evaluation | 275 Copy Task Switchboard training sentences.<br>120 Copy Task Switchboard evaluation sentences. |
| 9 | 67 | 125,000-word training data collection and other experiments | 10 Copy Task Switchboard training sentences. |
| 10 | 69 | 125,000-word training data collection, evaluation, and personal use | 170 Copy Task Switchboard training sentences.<br>115 Copy Task Switchboard evaluation sentences.<br>180 Conversation Mode sentences. |
| 11 | 74 | 125,000-word training data collection, evaluation, and personal use | 120 Copy Task Switchboard training sentences.<br>50 Copy Task OpenWebText training sentences.<br>75 Copy Task Switchboard evaluation sentences.<br>140 Conversation Mode sentences. |
| 12 | 76 | 125,000-word training data collection, evaluation, and personal use | 50 Copy Task Switchboard training sentences.<br>40 Copy Task OpenWebText training sentences.<br>210 Copy Task Switchboard evaluation sentences.<br>85 Conversation Mode sentences. |
| 13 | 81 | 125,000-word training data collection and evaluation | 110 Copy Task Switchboard training sentences.<br>140 Copy Task Switchboard evaluation sentences. |
| 14 | 83 | 125,000-word training data collection, evaluation, and personal use | 115 Copy Task Switchboard training sentences.<br>45 Copy Task OpenWebText training sentences.<br>170 Copy Task Switchboard evaluation sentences.<br>280 Conversation Mode sentences. |
| 15 | 88 | 125,000-word training data collection, evaluation, and personal use | 140 Copy Task Switchboard training sentences.<br>90 Copy Task Switchboard evaluation sentences.<br>100 Conversation Mode sentences. |
| 16 | 90 | 125,000-word training data collection, evaluation, and personal use | 140 Copy Task Switchboard training sentences.<br>40 Copy Task OpenWebText training sentences.<br>150 Copy Task Switchboard evaluation sentences.<br>140 Conversation Mode sentences. |
| 17 | 95 | 125,000-word training data collection, evaluation, and personal use | 50 Copy Task Switchboard training sentences.<br>20 Copy Task Harvard training sentences.<br>50 Copy Task Switchboard evaluation sentences.<br>140 Conversation Mode sentences. |
| 18 | 97 | 125,000-word training data collection and personal use | 100 Copy Task Switchboard training sentences.<br>30 Copy Task Harvard training sentences.<br>180 Conversation Mode sentences. |
| 19 | 102 | Other data collection and personal use | 86 Copy Task Conversation Mode sentences. |
| 20 | 104 | Other data collection and personal use | 86 Conversation Mode sentences. |
| 21 | 109 | Other data collection and personal use | 50 Copy Task Switchboard training sentences.<br>50 Copy Task Harvard training sentences.<br>150 Copy Task frequent word training sentences.<br>51 Conversation Mode sentences. |
| 22 | 110 | Other data collection and personal use | 110 Copy Task frequent word training sentences.<br>82 Conversation Mode sentences. |
| 23 | 114 | Personal use | 291 Conversation Mode sentences. |
| 25 | 123 | Other data collection. | 150 Copy Task frequent word training sentences. |
| 26 | 125 | Other data collection and personal use | 100 Copy Task frequent word training sentences.<br>198 Conversation Mode sentences. |
| 27 | 127 | Personal use | 439 Conversation Mode sentences. |
| 29 | 132 | Other data collection and personal use | 240 Copy Task frequent word training sentences.<br>50 Copy Task random word training sequences.<br>115 Conversation Mode sentences. |
| 30 | 134 | Personal use | 287 Conversation Mode sentences |
| 31 | 137 | Other data collection and personal use | 43 Copy Task switchboard training sentences.<br>26 Copy Task frequent word training sentences.<br>94 Conversation Mode sentences. |
| 32 | 139 | Other data collection and personal use | 50 Copy Task switchboard training sentences.<br>250 Copy Task frequent word training sentences.<br>156 Conversation Mode sentences. |
| 33 | 141 | Personal use | 278 Conversation Mode sentences |
| 34 | 144 | Other data collection and personal use | 200 Copy Task frequent word training sentences.<br>18 Conversation Mode sentences. |
| 35 | 146 | Other data collection and personal use | 250 Copy Task frequent word training sentences.<br>369 Conversation Mode sentences. |
| 36 | 148 | Personal use | 627 Conversation Mode sentences. |
| 37 | 149 | Personal use | 261 Conversation Mode sentences. |
| 38 | 153 | Other data collection and personal use | 195 Copy Task frequent word training sentences.<br>208 Conversation Mode sentences. |
| 39 | 156 | Personal use | 555 Conversation Mode sentences. |
| 40 | 163 | Personal use | 652 Conversation Mode sentences. |
| 41 | 165 | Other data collection and personal use | 50 Copy Task Switchboard training sentences.<br>35 Copy Task spelling training sentences.<br>250 Copy Task frequent word training sentences.<br>255 Conversation Mode sentences. |
| 42 | 170 | Personal use | 513 Conversation Mode sentences. |
| 43 | 171 | Other data collection and personal use | 274 Conversation Mode sentences. |
| 44 | 172 | Other data collection and personal use | 320 Copy Task frequent word training sentences.<br>30 Copy Task random word training sentences.<br>86 Conversation Mode sentences. |
| 45 | 174 | Other data collection and personal use | 50 Copy Task Switchboard training sentences.<br>92 Copy Task frequent word training sentences.<br>216 Conversation Mode sentences. |
| 46 | 176 | Personal use | 20 Copy Task Switchboard training sentences.<br>296 Conversation Mode sentences. |
| 47 | 177 | Personal use | 284 Conversation Mode sentences. |
| 48 | 179 | Other data collection and personal use | 290 Copy Task frequent word training sentences.<br>10 Copy Task random word training sentences.<br>348 Conversation Mode sentences. |
| 49 | 181 | Other data collection and personal use | 190 Copy Task frequent word training sentences.<br>406 Conversation Mode sentences. |
| 50 | 183 | Personal use | 559 Conversation Mode sentences. |
| 51 | 184 | Personal use | 405 Conversation Mode sentences. |
| 52 | 186 | Other data collection and personal use | 301 Copy Task frequent word training sentences.<br>289 Conversation Mode sentences. |
| 53 | 188 | Other data collection and personal use | 50 Copy Task Switchboard training sentences.<br>240 frequent word training sentences. |
| 54 | 190 | Personal use | 20 Copy Task Switchboard training sentences.<br>384 Conversation Mode sentences. |
| 55 | 191 | Personal use | 554 Conversation Mode sentences. |
| 56 | 193 | Other data collection and personal use | 20 Copy Task random word training sentences.<br>100 Copy Task frequent word training sentences.<br>320 Copy Task 50-word vocabulary training sentences.<br>171 Conversation Mode sentences. |
| 57 | 195 | Other data collection and personal use | 200 Copy Task frequent word training sentences.<br>112 Copy Task 50-word vocabulary training sentences.<br>205 Conversation Mode sentences. |
| 58 | 202 | Other data collection and personal use | 30 Copy Task Switchboard training sentences.<br>223 Conversation Mode sentences. |
| 59 | 204 | Personal use | 553 Conversation Mode sentences. |
| 60 | 205 | Personal use | 756 Conversation Mode sentences. |
| 61 | 207 | Other data collection and personal use | 50 Copy Task Switchboard training sentences.<br>25 Copy Task spelling training sentences.<br>25 Copy Task proper noun training sentences.<br>475 Conversation Mode sentences. |
| 62 | 209 | Other data collection and personal use | 35 Copy Task Switchboard training sentences.<br>351 Conversation Mode sentences. |
| 63 | 211 | Personal use | 557 Conversation Mode sentences. |
| 64 | 212 | Personal use | 373 Conversation Mode sentences. |
| 65 | 214 | Other data collection and personal use | 264 Conversation Mode sentences. |
| 66 | 216 | Other data collection and personal use | 25 Copy Task Switchboard training sentences.<br>272 Conversation Mode sentences. |
| 67 | 218 | Personal use | 819 Conversation Mode sentences. |
| 68 | 219 | Personal use | 722 Conversation Mode sentences. |
| 69 | 223 | 125,000-word training data collection, evaluation, and personal use | 100 Copy Task Switchboard training sentences.<br>150 Copy Task Switchboard evaluation sentences.<br>306 Conversation Mode sentences. |
| 70 | 225 | Personal use | 417 Conversation Mode sentences. |
| 71 | 226 | Personal use | 288 Conversation Mode sentences. |
| 72 | 288 | Other data collection and personal use | 20 Copy Task Switchboard training sentences.<br>207 Conversation Mode sentences. |
| 73 | 230 | Other data collection and personal use | 20 Copy Task Switchboard training sentences.<br>200 Copy Task 50-word training sentences.<br>207 Conversation Mode sentences. |
| 74 | 232 | Personal use | 662 Conversation Mode sentences. |
| 75 | 233 | Personal use | 499 Conversation Mode sentences. |
| 76 | 235 | 125,000-word training data collection and personal use | 170 Copy Task Switchboard training sentences.<br>101 Conversation Mode sentences |
| 77 | 239 | Personal use | 20 Copy Task Switchboard training sentences.<br>873 Conversation Mode sentences. |
| 78 | 240 | Personal use | 486 Conversation Mode sentences. |
| 79 | 242 | Other data collection and personal use | 300 Copy Task Switchboard training sentences<br>324 Conversation Mode sentences |
| 80 | 244 | 125,000-word training data collection, evaluation, and personal use | 100 Copy Task Switchboard training sentences<br>150 Copy Task switchboard evaluation sentences<br>100 Copy Task Switchboard SILENT SPEECH training sentences<br>216 Conversation Mode sentences |
| 81 | 246 | Personal use | 676 Conversation Mode sentences |
| 82 | 247 | Personal use | 583 Conversation Mode sentences |
| 83 | 249 | Other data collection and personal use | 50 Copy Task Switchboard training sentences<br>222 Conversation Mode sentences |
| 84 | 251 | Silent speech training data collection, evaluation, and personal use | 300 Copy Task Switchboard SILENT SPEECH sentences<br>408 Conversation Mode sentences |

**Table S3:** Online decoding performance (125,000 word vocabulary).

| Session number | Post-implant day | Hours of training data | Number of training sentences | Number of evaluation sentences | Raw phoneme error rate (95% CI) | Word error rate (95% CI) | Words per minute (95% CI) |
| --- | --- | --- | --- | --- | --- | --- | --- |
| 2 | 27 | 1.9 | 608 | 30 | 15.36<br>(12.05, 18.87) | 9.84<br>(4.10, 16.04) | 36.73<br>(35.93, 37.62) |
| 4 | 34 | 4.0 | 1147 | 100 | 12.82<br>(11.15, 14.52) | 13.61<br>(10.28, 17.12) | 32.44<br>(32.09, 32.80) |
| 7 | 46 | 6.6 | 1749 | 99 | 9.36<br>(7.80, 10.98) | 4.61<br>(2.61, 6.95) | 33.08<br>(32.48, 33.63) |
| 8 | 48 | 8.0 | 2085 | 115 | 11.74<br>(10.09, 13.39) | 5.40<br>(3.52, 7.57) | 31.95<br>(31.46, 32.42) |
| 10 | 69 | 8.9 | 2296 | 115 | 12.45<br>(10.57, 14.40) | 6.15<br>(3.94, 8.66) | 28.23<br>(27.79, 28.66) |
| 11 | 74 | 10.1 | 2535 | 74 | 10.17<br>(8.51, 11.91) | 3.89<br>(1.92, 6.22) | 31.52<br>(30.77, 32.37) |
| 12 | 76 | 11.5 | 2878 | 208 | 10.36<br>(8.99, 11.77) | 4.96<br>(3.31, 6.87) | 37.72<br>(36.57, 38.93) |
| 13 | 81 | 12.2 | 3047 | 138 | 10.58<br>(9.16, 12.06) | 4.06<br>(2.67, 5.66) | 31.65<br>(31.09, 32.26) |
| 14 | 83 | 13.4 | 3321 | 169 | 10.18<br>(8.85, 11.57) | 4.37<br>(2.99, 5.86) | 34.41<br>(33.65, 35.21) |
| 15 | 88 | 15.9 | 3826 | 90 | 6.93<br>(5.44, 8.45) | 2.54<br>(0.99, 4.45) | 32.26<br>(31.63, 32.87) |
| 16 | 90 | 17.6 | 4209 | 148 | 7.27<br>(6.19, 8.41) | 3.25<br>(2.00, 4.66) | 33.78<br>(33.00, 34.61) |
| 17 | 95 | 18.8 | 4444 | 50 | 7.76<br>(5.85, 9.76) | 0.99<br>(0.00, 2.30) | 31.52<br>(30.75, 32.32) |
| 69 | 223 | 80.9 | 17645 | 146 | 9.33<br>(7.94, 10.77) | 3.10<br>(2.00, 4.32) | 31.52<br>(30.75, 32.32) |
| 80 | 244 | 94.5 | 20893 | 146 | 7.33<br>(6.20, 8.46) | 1.82<br>(1.01, 2.76) | 27.75<br>(27.43, 28.06) |

**Table S4:** Additional selected personal use transcripts.^1^.

| Context | Selected transcripts | WER |
| --- | --- | --- |
| Session 11: SP2 thanks a member of the research team for complimenting his plants | <u>SP2:</u> testing testing one two<br><u>SP2:</u> hello how is everyone<br><u>SP2:</u> thank you for complimenting my rubber and stick [snake] plan [plants]<br>...<br><u>SP2:</u> when i first got them they were in a one inch pot<br><u>SP2:</u> and they resided on my window sill<br><u>SP2:</u> hell yeah<br><u>SP2:</u> your degree is not in botany<br><u>SP2:</u> you know the saying that before you have kids you need to successfully take care of place [plants] in [and] the [then] past [pets]<br><u>SP2:</u> the progression is place [plants] and then past [pets] | 11.3% |
| Session 15: SP2 gives feedback to SDS (author) about how he's enjoying using the speech neuroprosthesis for conversational speech. | <u>SP2:</u> he should know better than to ask me what i want to say<br><u>SP2:</u> i'm a smart ass<br><u>SP2:</u> does he want me to talk in english or russian<br><u>SP2:</u> yes it should<br><u>SP2:</u> we can ease into it by talking in spanish first<br>...<br><u>SP2:</u> have you noticed anything different with the program<br><u>SP2:</u> it is way more accurate than before<br><u>SP2:</u> it is about ninety eight percent accurate by my informal estimation<br>...<br><u>SP2:</u> thank you again for all the improvements and the time it took to make them | 0% |
| Session 17: SP2 gives feedback about using the neural-click decoder (right hand squeeze) to signal the end of sentences. Then he suggests adding a third confirmation button "mostly correct" (in addition to "100% correct" and "incorrect") to the self-initiated conversational task. | <u>SP2:</u> does it make a sound when i press the button<br><u>SP2:</u> that makes sense<br><u>SP2:</u> do you want some feedback about my right hand squeezing<br><u>SP2:</u> so i can actually still squeeze my hand<br><u>SP2:</u> but it was not a problem because i really had to intend to raise [squeeze] my hand or it would not work<br><u>SP2:</u> the only problem was if i was running [yawning]<br><u>SP2:</u> so when i was doing that i would involuntarily raise [squeeze] my hand and that would trigger the complete button<br><u>SP2:</u> but initially my fears were mainly unfounded<br><u>SP2:</u> yes i do but not because i didn't like using the eye tracker method but because i like having multiple options<br><u>SP2:</u> thank you<br>...<br><u>SP2:</u> have you thought about adding a third party [button] that is almost correct<br><u>SP2:</u> yes that would be good<br><u>SP2:</u> we can try the eye tracker<br><u>SP2:</u> how many rolls [trials] do you need in total<br><u>SP2:</u> let's do one country [hundred] at a time | 4.7% |
| Session 18: SP2 is telling a friend about the speech decoder. | <u>SP2:</u> testing testing<br><u>SP2:</u> thank you<br><u>SP2:</u> what i was trying to say is that i have noticed that the computer has the same problem understanding what i am saying that people have meaning the same exact words that people have a problem with the computer also has a problem understanding<br><u>SP2:</u> totally | 4.9% |
|  | <u>SP2:</u> yes i can in the last session the computer had problems with understanding when i said next and it made the mistake of typing this instead of next and i thought that people who can understand me often make the same mistake<br><u>SP2:</u> totally<br>...<br><u>SP2:</u> when it is thinking of what to write it is because it is confused about what i said and it is running through different models of possible sentences<br><u>SP2:</u> know [no] what i am doing is all the same type of degree [decoding]<br><u>SP2:</u> the last word should have be be [been] guarding [decoding]<br><u>SP2:</u> so we have found that there is a slight increase in accuracy when i realize [vocalize] what i am saying but we think that is possibly true because the model trained on this approach rather than me doing what i am doing now<br><u>SP2:</u> the word really [realize] should have been vocally [vocalize]<br>...<br><u>SP2:</u> we are in the future<br><u>SP2:</u> on this post [wednesday] i will have my first day of only personal use<br><u>SP2:</u> on this coming west [wednesday]<br><u>SP2:</u> why can you not see [say] mr [wednesday]<br><u>SP2:</u> and it is having a problem with hearing me when i am saying that particular day of the week and now i am talking a little shit to it |  |
| Session 31: An interviewer is asking SP2 about his experience with using the speech decoder. | <u>Interviewer:</u> Hello [SP2], thank you so much for letting us into your home and taking the time to talk to us, it is nice to meet you.<br><u>SP2:</u> well it is nice to meet you and also your excellent camera made [man]<br><u>SP2:</u> your name is [camera operator's name] right<br><u>Camera operator:</u> Yup! Nice to meet you. Thank you for letting us into your home.<br><u>SP2:</u> that is okay i will be very happy to have both of you here to witness what i can do with my super powers<br><u>Interviewer:</u> So tell me [SP2] why are you doing this, and what it means for you to be a part of this.<br><u>SP2:</u> well i have this terrible disease and it is slowly taking away my ability to move and to talk<br><u>Interviewer:</u> I am so sorry, [SP2].<br><u>SP2:</u> it is fine but i am not a fan [ashamed] of you seeing me cry<br><u>SP2:</u> it should have said that i am not at camp [ashamed] to have you see me cry<br><u>SP2:</u> so because i have this terrible disease i have had the pleasure of meeting some amazing people like the nurse [ones] here<br><u>SP2:</u> the one [ones] where [here]<br><u>SP2:</u> i did my homework when i was thinking about having brain surgery because it was not an easy decision for me but i trusted the team that was behind me and i asked david if he would do this if he was in my position and while i will not tell you what he said it helped me make my decision with confidence<br><u>Interviewer:</u> Okay, how did it help you?<br><u>SP2:</u> i think that i understand that there are human beings that are behind all of this science and technology where they are thinking about what would be better for me and my family rather than simply thinking about me as a test subject<br><u>SP2:</u> does that make sense to you<br><u>Interviewer:</u> Yes, very much it does, and I can assure you that protecting you and your family is at the top of their list. I would like to thank you again, and I want to ask you how this has helped you communicate with your loved ones?<br><u>SP2:</u> why are you making me cry again<br><u>Interviewer:</u> It's okay if you'd rather move on to a different subject.<br><u>SP2:</u> i can definitely answer that question i was just giving you a hard time to lighten the mood | 2.7% |
|  | SP2: i really do not have much time and opportunity to use my **home** [humor] when i am relying on other people to translate what i say so please indulge my attempts at **home** [humor] because i really miss making jokes<br>Interviewer: So tell me about communicating, how is this helping you?<br>SP2: i have absolutely loved talking to my friends and family again without help from other people who can still understand to me<br>SP2: so when my symptoms started my daughter was only [redacted]<sup>2</sup> and now she is [redacted]<sup>2</sup> and she doesn't remember what i sounded like before this disease took away my ability to talk normally and she was a little shy at first but now is super proud that her **mother** [father] is a robot<br>SP2: that has been the **lead** [highlight] but i would also say that it is it **possible** [pretty cool] that i have been able to talk to other adults who do remember what i sounded like and they have been brought to tears to hear me again<br>Interviewer: That's profound!<br>SP2: so what people have told me is that they can totally understand what this is as a concept but when they see it in action it is [a] totally different type of experience<br>Interviewer: Definitely.<br>SP2: i have been able to talk to my parents over it and keep up with the conversation because they are from the south and talk really really really slowly<br>SP2: so i have been able to use this device to help me communicate with my colleagues who are working far away from here and i am on **mobile** [meetings] with them and this will work fine to help me communicate on calls<br>Interviewer: That's great.<br>SP2: it really is because i have an awesome job and i feel like people have really invested in me to help make me who i am and i feel like i have a lot of really important work left to do and this will help me do it<br>SP2: one of the things that people with my disease suffer from is isolation and depression because they do not feel like they matter anymore and something like this technology will help bring people back into life and into society<br>SP2: that really cannot be understated how important that is<br>SP2: because we will probably not find a cure for this disease but we will find medicines that help people live longer but if they are completely miserable than what is the point and something like this could really add value to people's life<br>SP2: so i think about this from my personal perspective and also the perspective of people like me who might not be as lucky but deserve the same treatment<br>SP2: does that make sense<br>Interviewer: Yes it does, absolutely.<br>SP2: what else would you like to know<br>Interviewer: I think you covered pretty much most of the things I had in mind. I loved hearing what you had to say and engaging in this conversation with you which was amazing with the help of this technology. And your sense of humor! If there's anything else that you would like to share, I am here to hear it.<br>SP2: i hope that we are very close to the time when everyone who is in a position like me has the same option to have this device as i do<br>Interviewer: I hope so too. I want to thank you so much, [SP2].<br>SP2: let's make it happen okay<br>Interviewer: Yes, and thank you so much.<br>SP2: my pleasure really<br>Camera operator: That was wonderful, thanks for letting us into your home.<br>SP2: of course my pleasure |  |

<sup>1</sup>Transcripts included here are non-exhaustive and exclude sensitive personal conversations where SP2 used the speech neuroprosthesis to converse with friends, family members, or medical professionals. Selected transcripts show snippets of conversations SP2 had with the research team or others. Gaps in time between transcribed sentences are represented by ellipses (...). There are no gaps in the Session 31 transcription with the interviewer. Reported word error rates (WER) in this table apply only to the provided transcripts and not to the sessions beyond these provided transcripts. Incorrectly decoded words are colored **red**, followed by the word that SP2 meant to say (confirmed with him) in [green brackets].
<sup>2</sup>Potentially identifiable information has been removed from these transcripts as per medRxiv policy.

**Table S5:** Decoding parameters.

| Parameter Category | Parameter Name | Parameter Description | Parameter Value |
| --- | --- | --- | --- |
| Recurrent neural network offline training | # Units | Number of units in each GRU layer | 512 |
|  | # Layers | Number of GRU layers | 5 |
|  | Kernel Size | Number of input feature time bins stacked together as a single input for the RNN | 14 |
|  | Kernel Stride | Number of time steps the RNN skips forward every step | 4 |
|  | L2 | L2 regularization cost | 1e-0.5 |
|  | Dropout | Probability of dropout during training for GRU layers | 0.4 |
|  | Input # units | Number of units in each input layer | 512 |
|  | Input activation | Activation function of input layers | softsign |
|  | Input dropout | Probability of dropout during training for input layers | 0.2 |
|  | White noise SD | Standard deviation of white noise added to input data for regularization | 1.0 |
|  | Smooth kernel SD | Amount of temporal gaussian smoothing of neural data during training | 2 |
|  | Constant offset SD | Standard deviation of constant offset noise added to input data to improve robustness against non-stationarity feature means | 0.2 |
|  | Batch Size | Number of sentences included in each batch | 64 |
|  | Training Batches | Number of training batches, varied depending on amount of training data | 2,000 to 50,000 |
|  | Learning Rate | Linearly decaying learning rate | 0.02 to 0.00 |
| | $\beta_1$ | ADAM stochastic gradient descent parameter | 0.9 |
| | $\beta_2$ | ADAM stochastic gradient descent parameter | 0.999 |
| | $\epsilon$ | ADAM stochastic gradient descent parameter | 0.1 |
| Recurrent neural network online training | Learning Rate | Learning rate used during online fine-tuning | 0.004 |
|  | Batch size | Number of sentences included in each batch | 64 |
|  | New data percent | Percent of data per batch from new data from the current session | 0.6 |
|  | Min training steps | Minimum number of training epochs to execute | 32 |
|  | Max training steps | Maximum number of training epochs to execute | 200 |
|  | Loss threshold | Loss threshold to stop training once under | 0.5 to 0.6 |
|  | White noise SD | Standard deviation of white noise added to input data for regularization | 1.0 |
|  | Smooth kernel SD | Amount of temporal gaussian smoothing of neural data during training | 2 |
|  | Constant offset SD | Standard deviation of constant offset noise added to input data to improve robustness against non-stationarity feature means | 0.2 |
| N-gram Language Model | blank penalty | Penalty applied on blank labels | log(9) |
|  | acoustic scale | Scaling factor on RNN's log probabilities | 0.3 |
|  | Min active | Beam search decoder's minimum active states | 200 |
|  | Max active | Beam search decoder's maximum active states | 7000 |
|  | beam | Beam size | 17 |
|  | n-best | Number of decoding hypotheses | 100 |
| Large Language Model | alpha | Interpolation weight between LMs | 0.5 |
|  | acoustic scale | Scaling factor on RNN's log probabilities | 0.3 |

## Notes

Funding: - NS Card: ○ A.P. Giannini Postdoctoral Fellowship - JM Henderson: ○ Stanford Wu Tsai Neurosciences Institute ○ HHMI ○ NIH-NIDCD (1U01DC019430) - SD Stavisky: ○ DP2 from the NIH Office of the Director and managed by NIDCD (1DP2DC021055) ○ DOD CDMRP ALS Pilot Clinical Trial Award (AL220043) ○ Pilot Award from the Simons Collaboration for the Global Brain (872146SPI) ○ Searle Scholars Program ○ Career Award at the Scientific Interface from the Burroughs Wellcome Fund ○ UC Davis Early Career Faculty Award for Creativity and Innovation - LR Hochberg: ○ NIH-NIDCD (U01DC017844) ○ VA RR&D (A2295-R)

### Competing Interest Statement

Stavisky, Henderson, and Willett, are inventors on intellectual property owned by Stanford University that has been licensed to Blackrock Neurotech and Neuralink Corp. Wairagkar, Stavisky, and Brandman have patent applications related to speech BCI owned by the Regents of the University of California. Stavisky was an advisor to wispr.ai and received equity. Brandman is a surgical consultant to Paradromics Inc. Henderson is a consultant for Neuralink Corp, serves on the Medical Advisory Board of Enspire DBS and is a shareholder in Maplight Therapeutics. The MGH Translational Research Center has a clinical research support agreement with Neuralink, Synchron, Axoft, Precision Neuro, and Reach Neuro, for which LRH provides consultative input. Mass General Brigham (MGB) is convening the Implantable Brain-Computer Interface Collaborative Community (iBCI-CC); charitable gift agreements to MGB, including those received to date from Paradromics, Synchron, Precision Neuro, Neuralink, and Blackrock Neurotech, support the iBCI-CC, for which LRH provides effort. Glasser is a consultant to Sora Neuroscience, Manifest Technologies, and Turing Medical.

### Clinical Trial

NCT00912041

### Funding Statement

This study was funded by the following funding sources:
NS Card: A.P. Giannini Postdoctoral Fellowship
JM Henderson: Stanford Wu Tsai Neurosciences Institute, HHMI, NIH-NIDCD (1U01DC019430)
SD Stavisky: DP2 from the NIH Office of the Director and managed by NIDCD (1DP2DC021055), DOD CDMRP ALS Pilot Clinical Trial Award (AL220043), Pilot Award from the Simons Collaboration for the Global Brain (872146SPI), Searle Scholars Program, Career Award at the Scientific Interface from the Burroughs Wellcome Fund, UC Davis Early Career Faculty Award for Creativity and Innovation
LR Hochberg: NIH-NIDCD (U01DC017844), VA RR&D (A2295-R)

### Author Declarations

The IRB of University of California, Davis gave ethical approval for this work (protocol #1843264).

### Summary of Updates

The manuscript and supplemental appendix were revised in response to peer review. Existing figures were rearranged. New data, analyses, and figures were added. Aparna Srinivasan was added as an author because she contributed to data collection and analyses during the revision process.

